# Ebola Virus Disease mathematical models and epidemiological parameters: a systematic review and meta-analysis

**DOI:** 10.1101/2024.03.20.24304571

**Authors:** Rebecca K. Nash, Sangeeta Bhatia, Christian Morgenstern, Patrick Doohan, David Jorgensen, Kelly McCain, Ruth McCabe, Dariya Nikitin, Alpha Forna, Gina Cuomo-Dannenburg, Joseph T. Hicks, Richard J. Sheppard, Tristan Naidoo, Sabine van Elsland, Cyril Geismar, Thomas Rawson, Sequoia Iris Leuba, Jack Wardle, Isobel Routledge, Keith Fraser, Pathogen Epidemiology Review Group, Natsuko Imai-Eaton, Anne Cori, H. Juliette T. Unwin

**Affiliations:** MRC Centre for Global Infectious Disease Analysis & WHO Collaborating Centre for Infectious Disease Modelling, Jameel Institute, School of Public Health, Imperial College London, UK; Health Protection Research Unit in Modelling and Health Economics; Modelling and Economics Unit, UK Health Security Agency, London, UK; Department of Statistics, University of Oxford, Oxford, UK; Health Protection Research Unit in Emerging and Zoonotic Infections. University of Liverpool, Liverpool, UK; Center for the Ecology of Infectious Diseases, Odum School of Ecology, University of Georgia, Athens, GA, USA; Institute of Global Health Sciences, University of California, San Francisco, USA; School of Mathematics, University of Bristol, Bristol, UK

**Keywords:** Ebola virus disease, systematic review, epidemiological parameters, mathematical models

## Abstract

**Background:** Ebola Virus Disease (EVD) poses a recurring risk to human health. Modelling can provide key insights informing epidemic response, hence synthesising current evidence about EVD epidemiology and models is critical to prepare for future outbreaks.

**Methods:** We conducted a systematic review (PROSPERO CRD42023393345) and meta-analysis of EVD transmission models and parameters characterising EVD transmission, evolution, natural history, severity, risk factors and seroprevalence published prior to 7th July 2023 from PubMed and Web of Science. Two people screened each abstract and full text. Papers were extracted using a bespoke Access database, 10% were double extracted. Meta**-** analyses were conducted to synthesise information where possible.

**Findings:** We extracted 1,280 parameters and 295 models from 522 papers. Basic reproduction number estimates were highly variable (central estimates between 0.1 and 12.0 for high quality assessment scores), as were effective reproduction numbers, likely reflecting spatiotemporal variability in interventions. Pooled random effect estimates were 15.4 days (95% Confidence Interval (CI) 13.2-17.5) for the serial interval, 8.5 (95% CI 7.7-9.2) for the incubation period, 9.3 (95% CI 8.5-10.1) for the symptom-onset-to-death delay and 13.0 (95% CI 10.4-15.7) for symptom-onset-to-recovery. Common effect estimates were similar albeit with narrower CIs. Case fatality ratio estimates were generally high but highly variable (from 0 to 100%), which could reflect heterogeneity in underlying risk factors such as age and caring responsibilities.

**Interpretation:** While a significant body of literature exists on EVD models and epidemiological parameter estimates, many of these studies focus on the West African Ebola epidemic and are primarily associated with Zaire Ebola virus. This leaves a critical gap in our knowledge regarding other Ebola virus species and outbreak contexts.

**Funding:** UKRI, NIHR, Academy of Medical Sciences, Wellcome, UK Department for Business, Energy, and Industrial Strategy, BHF, Diabetes UK, Schmidt Foundation, Community Jameel, Royal Society, and Imperial College London.

**Research in Context:** *Evidence before this study:* We searched Web of Science and PubMed up to 7th July 2023 using the search terms: Ebola, epidemiology, outbreaks, models, transmissibility, severity, delays, risk factors, mutation rates and seroprevalence. We identified 179 reviews or overviews of different aspects of Ebola virus disease (EVD) transmission, of which we explored 11 that had “systematic” or “meta” in the title plus one included by expert recommendation. Five reviews focused on case fatality ratios, with estimates ranging between 34-42% for the Bundibugyo Ebola virus species, 53-69% for the Sudan species, 31.6-100% for the Zaire species, and pooled estimates ranging between 28-65% from reviews not specifying the species. Three reviews estimated seroprevalence to be between 3.3-8% depending on the setting and time. Three reviews investigated risk factors and found that caring for a case in the community and participation in traditional funeral rites are strongly associated with acquiring disease. Two reviews reported the incubation period to be 6.3 days for the Bundibugyo species, a range of 3.35-14 days for the Sudan species, and a range of 9-11.4 days across studies on the Zaire species. We found one review considering each of the following: basic reproduction number (1.34– 2.7 for Sudan species and 1.8 for Zaire species), serial interval (15-15.3 days for Zaire species), latent period (11.75 days for a combination of Zaire and unspecified species), and secondary attack rates (12.5%, species unspecified). Two reviews consider transmission models, identifying that it is difficult to accurately model the impact of time-dependent changing factors without high quality data, and data are often missing, complicating proper parameterisation of the underlying transmission mechanisms. One specific review looked at the Sudan EVD in response to the outbreak in Uganda in 2023, which highlighted the lack of vaccines and treatment available for this species.

*Added value of this study:* We provide a comprehensive summary of all available peer reviewed literature of transmission models and the variables needed to parameterise them across all EVD species and outbreaks. Our study synthesises all available analyses until 2023 and additionally considers attack rates, overdispersion and mutation rates. We give updated pooled random effects meta-analyses of incubation periods, serial intervals, symptom onset to death and symptom onset to recovery and, where possible, provide species-specific estimates in the Supplementary Material. We also provide ranges for the basic reproduction number and case fatality ratios without running meta-analyses because these are very setting dependent. We identify that most evidence (92%) is for the Zaire species and highlight that there are knowledge gaps for other species, which should be explored in the future. All our data is held within a bespoke open-source R package to enable others to use this information easily during their model building and updates.

*Implications of all the available evidence:* Previous outbreaks of infectious pathogens, including the 2013-2016 West African EVD epidemic, emphasise the usefulness of computational modelling in assessing epidemic dynamics and the impact of mitigation strategies. Our study provides an updated and broader overview of all the necessary information for designing and parameterising mathematical models for use in future outbreaks of EVD, including a centralised database for other researchers to use and contribute data to.

## Introduction

The COVID-19 pandemic, and multiple recent outbreaks of re-emerging pathogens, have highlighted the tremendous threat of infectious pathogens to the human population. The Democratic Republic of Congo (DRC) experienced an outbreak of Ebola virus disease (EVD) from 2018-2020. Seven other EVD outbreaks in DRC, Guinea and Uganda have been declared since. In Spring 2022, an outbreak of mpox affected several countries beyond the known endemic region (1). In February-March 2023, Equatorial Guinea and Tanzania faced a Marburg Virus Disease outbreak, the first since 2014 (2). These recurring events reinforce the need for pandemic preparedness. The World Health Organisation (WHO) has listed Ebola virus (EV) as one of nine pathogens posing the greatest threat to public health due to its high epidemic potential and lack of sufficient countermeasures (3).

EV is a deadly filovirus (4), transmitted through close contact and bodily fluids especially during traditional burials and caregiving, which has caused 38 known outbreaks since its discovery in 1976 (Table S1)(5). Most of these outbreaks have occurred in Central and Western Africa, and the largest epidemic, the West African (WA) Ebola epidemic, caused over 11,000 reported deaths between 2013 and 2016 mainly across Guinea, Liberia and Sierra Leone (6,7). Four species of EV are known to affect humans: Zaire, Bundibugyo, Sudan and Taï Forest. One species, Reston, is only known to cause disease in non-human primates (4) and a sixth species, Bombali, was identified in samples taken from bats in Sierra Leone (8). The symptoms of Ebola infection can be sudden and include fever, fatigue, muscle pain, headache and sore throat followed by vomiting, diarrhoea, rash, and internal and external bleeding.

Treatment of EVD involves supportive care, such as rehydration (intravenous fluids or oral rehydration) and the stabilisation of oxygen levels and blood pressure. There are two monoclonal antibody treatments recommended for confirmed cases of infection with the Zaire species, REGN-EB3 and mAb114, but access is limited due to uncertainties surrounding pricing and future supply (9). There are no licenced treatments for other EVD species. A vaccine (Ervebo), trialled against the Zaire species during the WA epidemic (10), is now used as part of outbreak response activities to prevent disease using a “ring vaccination” strategy. An alternative vaccine (Zabdeno/Mvabea) consists of two doses given eight weeks apart and is therefore not suitable for use in an emergency context where immediate protection is necessary (11). Three vaccine candidates for the Sudan species are in various trial phases (12). In the absence of widely available vaccines and therapeutics, mitigation of EVD outbreaks relies on a suite of public health and social measures such as case identification and isolation, contact tracing and quarantine, personal protective equipment for health care workers, safe and dignified burials, and community engagement.

Mathematical modelling and outbreak analytics are a component of monitoring and responding to epidemics and were used effectively to characterise transmission dynamics and severity during the WA epidemic and recent DRC outbreaks (13,14). Mathematical epidemic models typically use various parameters characterising the pathogen as inputs, for example the incubation and infectious periods, with the robustness of modelling outputs directly depending on the reliability of parameter values. Further, these parameters have direct implications for outbreak control; for example, the upper bound of the incubation period determines the necessary duration of follow-up for contacts of cases (13). Therefore, it is important to centralise evidence around these parameters, to inform the rapid design of mathematical models that could support the response to future EVD outbreaks. Here we undertake a systematic review and meta-analysis to build a database of EVD models and related parameters.

## Methods

PRISMA guidelines were used for this systematic review and checklists have been included in Tables S4 & S5.

### Search strategy and selection criteria

We searched PubMed and Web of Science databases for English peer-reviewed articles including EVD transmission models, parameters characterising EVD transmission, evolution, natural history, severity and seroprevalence, and risk factors published prior to 7th July 2023 (see Supplementary Material (SM) Section 2.1 for search terms). Each title and abstract and then full text were screened by two reviewers from a group of 15 (RKN, SB, CM, PD, DJ, KM, RM, AF, GC-D, JH, TN, IR, SvE, AC and HJTU) using Covidence (15) and inclusion / exclusion criteria from SM Section 2.2. Disagreements were resolved by consensus between reviewers. We used backward citajon chaining from 12 of the 179 review papers (16–27) identified during screen to add in missing papers (see SM Section 2.3).

19 reviewers (RKN, SB, CM, PD, KM, RM, DN, GC-D, JH, RS, TN, SvE, CG, TR, SIL, JW, KF, AC and HJTU) extracted data about the articles, models, and parameters from our included papers using Microsoft Access (see SM Section 2.4 for full list of parameters and extraction information). To ensure consistency of the data extraction process, data from 55 randomly selected papers (10%) were double extracted, with disagreements resolved by consensus. We used a customised questionnaire to assess the quality and risk of bias of each paper (SM Section 2.5).

### Data analysis

All analysis was done in R using the orderly2 R package (28) for workflow management (see SM Section 2.6 for full details). Curated and annotated data are made available in our R package epireview (29). We conducted meta-analyses for incubation periods, serial intervals and time from symptom onset to death or recovery, where the number of estimates exceeded at least two, using the metamean function from the meta R package (30). The metamean function was also used to perform sub-group meta-analyses to explore whether these delays varied by EV species. We did not do meta-analyses for other parameters due to too few data in the required format (see SM section 3.6), or high variability between the study settings.

Main text figures and tables only include parameters from articles with a high quality assessment (QA) score of at least 50% (see SM Section 2.5), and all parameters are included in the SM. Our full analysis can be reproduced using https://github.com/mrc-ide/priority-pathogens.

## Results

Our search returned 24,338 articles, which reduced to 14,690 after deduplication and the addition of two papers identified through other systematic reviews (Figure 1). Following title and abstract screening, 1,674 articles were retained for full text screening, with 522 meeting our inclusion criteria. We extracted 1,280 parameters (from 354 articles) (Table S8) and 295 models (from 280 articles). We could link 1,213 of the parameters to a specific EVD outbreak; the vast majority of these (71%, n=858) reported on the WA Ebola epidemic. 1,229 parameters could be linked to a specific EV species and 92% (n=1,136) were associated with the Zaire species.

**Figure 1:**
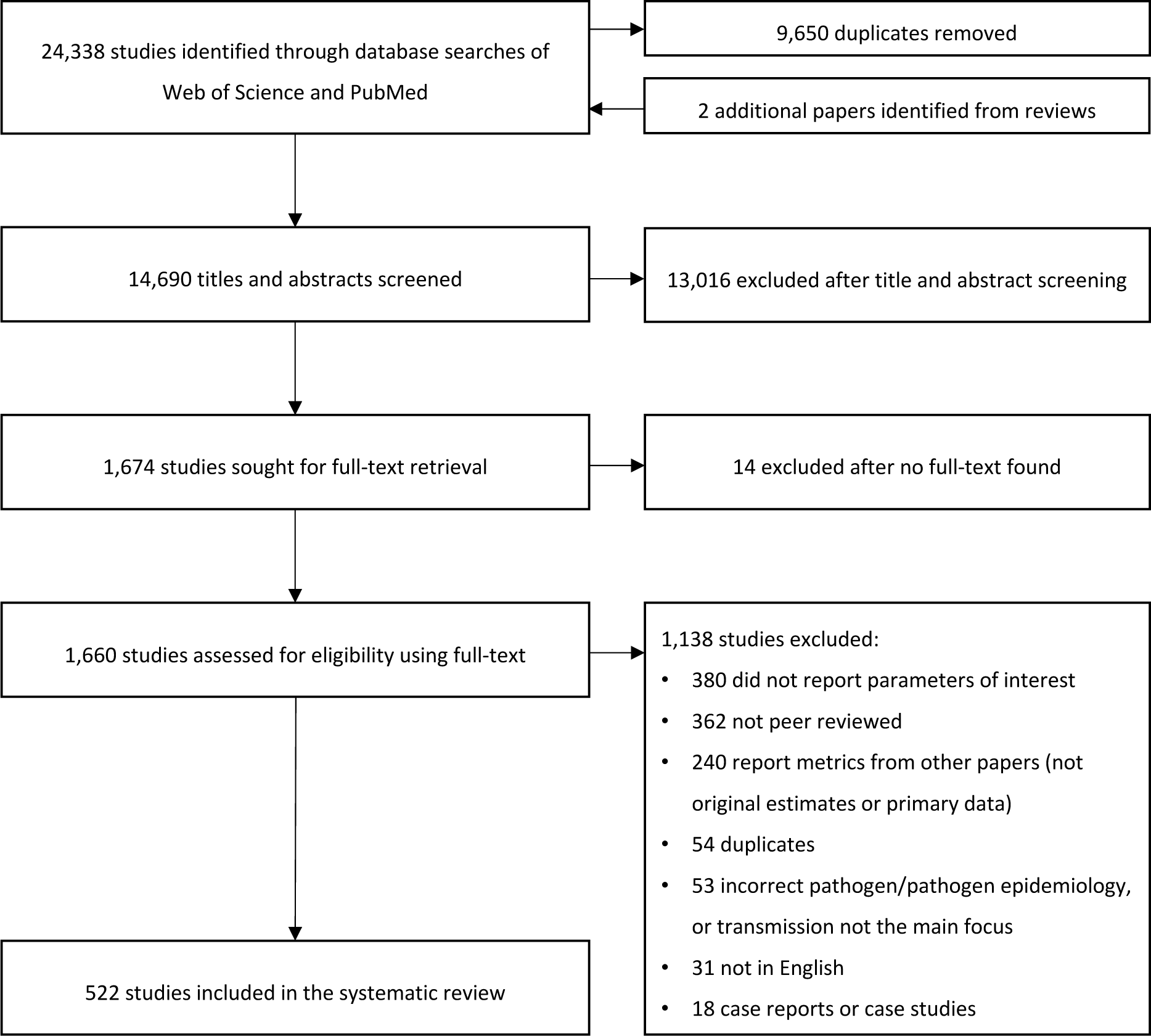
PRISMA flowchart illustrating the systematic review process.

Published EVD seroprevalence estimates were highest in countries with reported outbreaks (Table S1, Table S9), but varied depending on the population sampled. In the DRC, community-based seropositivity was between 0 and 18.7% across the different assays, whereas in hospitals it was between 0 and 37.0%. Similarly, in Guinea, a single population-level estimate was 0.07% but ranged from 59.4%-99.8% in hospitals. Despite no official outbreaks, seropositivity has been reported among population groups in Cameroon, Central African Republic, Kenya and Madagascar, Mali and Tanzania (Table S10).

Across articles reporting attack rates (Table S12), central estimates were all below 10%, except for one paper (31) which suggested that up to 31% of physicians had been infected in the 1995 outbreak in Kikwit, DRC. Most estimates (7/10) focused on the general population with generally low uncertainty.

Published estimates of the basic reproduction number (R_0_) were highly variable (Table S13). After removing papers with low QA scores (<50%), 71 R_0_ central estimates across 52 studies ranged from 0.05-12 (Figure 2 and Table 1A). 82% (n=58) of R_0_ estimates, including the most extreme central estimates, were for the 2013-16 WA epidemic; across all other outbreaks, R_0_ central estimates ranged from 1.1-7.7 (both for the 2018-20 DRC outbreak). Uncertainty around WA central estimates of R_0_ was also highly variable, with lower bounds of 0 and upper bounds of 18.5. 97% of R_0_ estimates (n=69) were for the Zaire species and two were for the Sudan species (from outbreaks in Uganda from 2000-01 and 2022-23). Sudan R_0_ central estimates were less variable than Zaire estimates (range 2.0 to 2.7).

**Figure 2:**
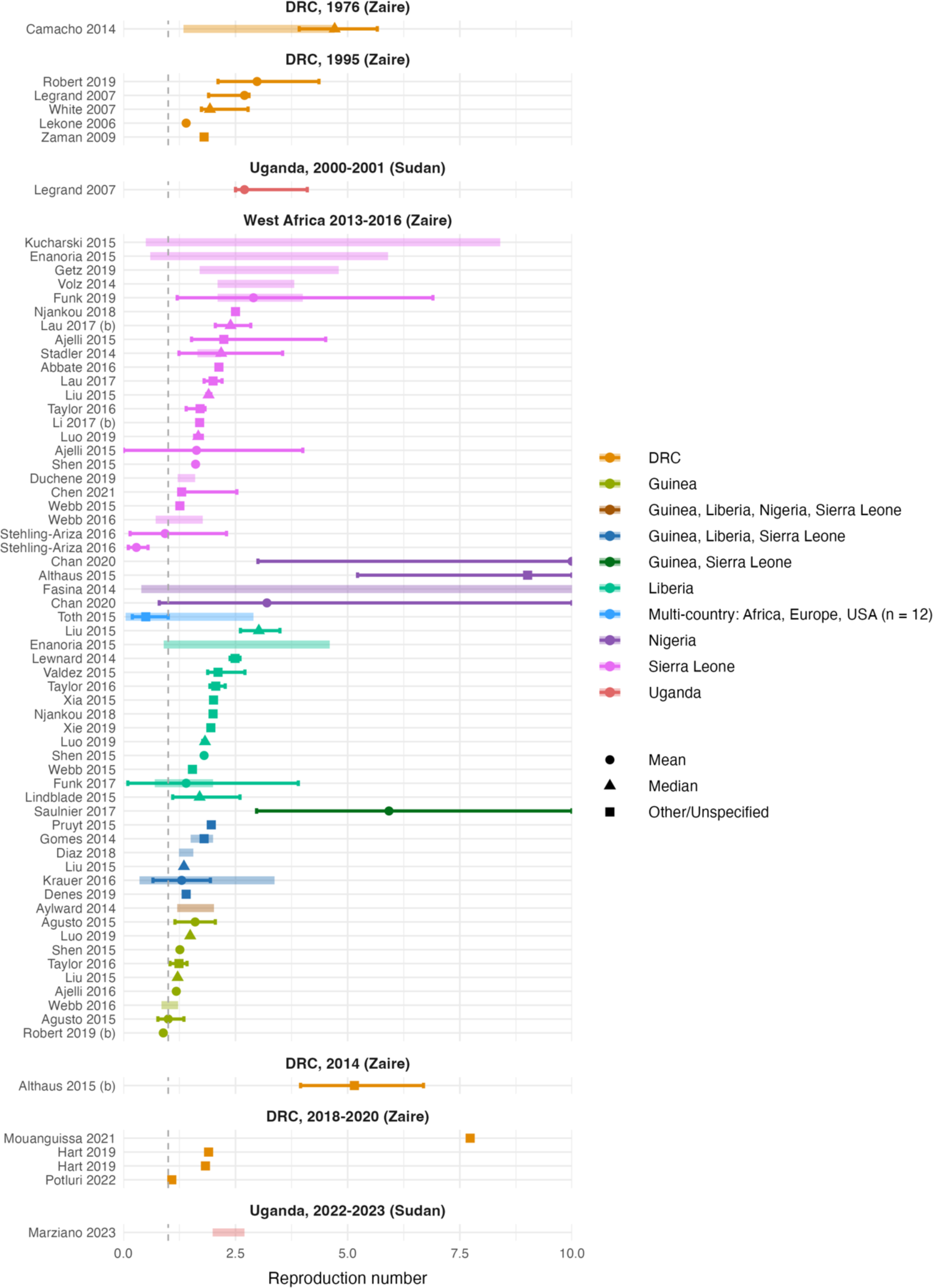
Basic reproduction numbers (R_0_) by outbreak. Each panel corresponds to a different outbreak of EVD with the associated EV species in brackets. Points represent central estimates, with symbol shapes corresponding to central value type. Thick coloured shaded lines represent the range of central estimates when R_0_ was estimated, for example, across regions or over time. Solid coloured bars represent the uncertainty around the central estimate; this was reported in different formats including standard deviation (in which case the bar represents +/− the standard deviation), 95% highest posterior density interval, range, interquartile range, 95% CrI (credible interval) and 95% CI. The x-axis has been restricted to a maximum of 10 for clarity. All parameters are from articles with a QA score of >= 50% (see Table S12 for all R_0_ estimates).

**Table 1:**
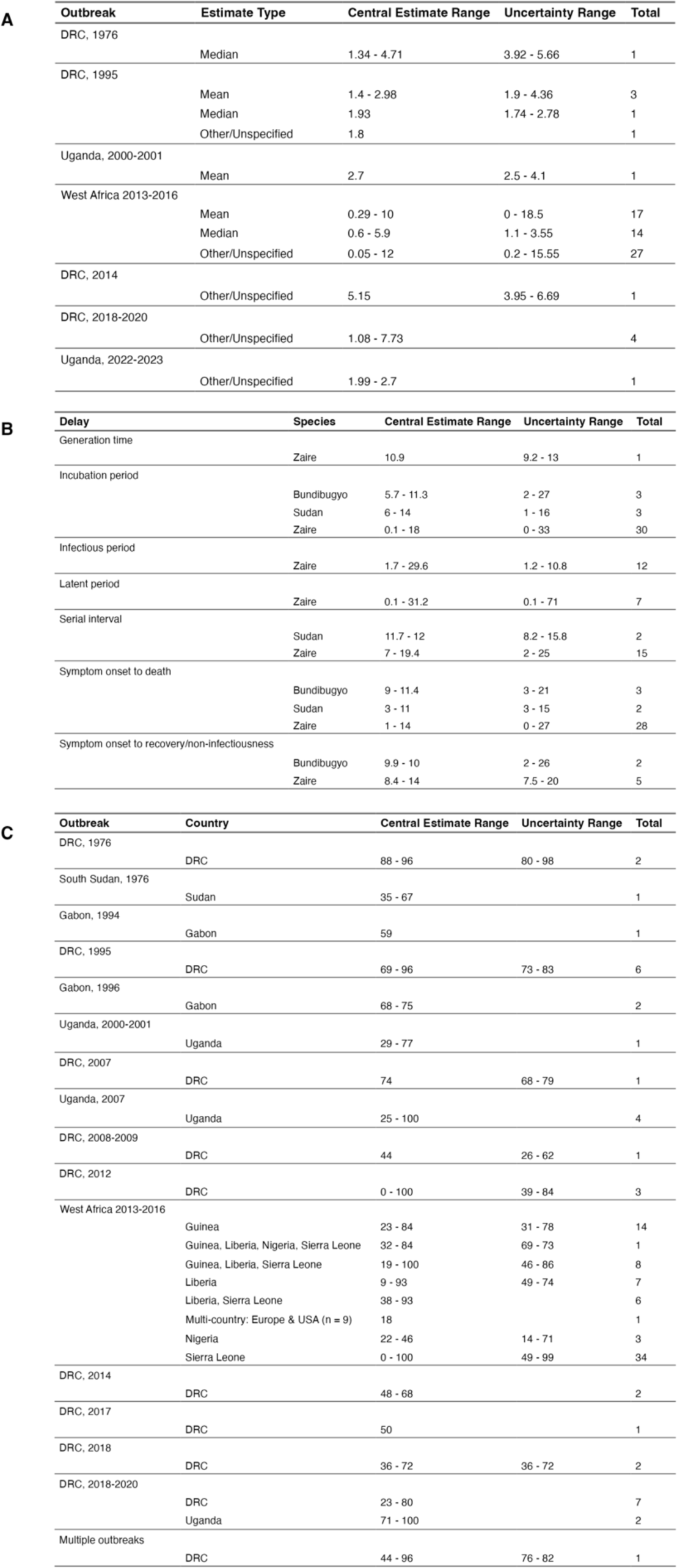
Ranges of estimates for A) basic reproduction numbers by outbreak and estimate type, B) epidemiological delays by Ebola virus species, C) Case Fatality Ratios (CFRs) by outbreak and country. The total column specifies the number of parameters (QA filtered >=50%) included in the summary range. Some parameter entries provide aggregated ranges of central estimates e.g. across time and countries (see SM Section 3.4) when more than three values were provided. Additionally, not all central estimates were reported with an associated uncertainty interval.

Estimates of the effective reproduction number, R_eff_, which measures transmissibility in the presence of potential interventions and population immunity, were also highly variable (Table S14). In 23 high QA studies (>=50%), central estimates ranged from 0-9 across 32 parameter estimates, of which all were for EV Zaire. The majority (72%, n=23) were for the WA epidemic, and the rest were all from the DRC, with only one estimate prior to the WA epidemic (0.73 in the 1995 DRC outbreak).

Estimates of secondary attack rates (SAR) (Table S15), growth rates (Table S16) and doubling times (Table S17) were similarly heterogeneous across studies, with central estimates of SAR in the range 0.1-89%, daily growth rates between 0.0 and 1.4 and doubling times up to decades.

15 estimates of overdispersion in the offspring distribution were extracted from 12 studies, with central estimates ranging from 0.02-2.2, with lower values indicating more overdispersion (Table S18). Other than one entry with an unspecified EV species, all were for Zaire (n=14), and most (87%, n=13) were for the WA Ebola epidemic.

31 studies examined risk factors for EV infection (Table S19). Conflicting findings were found across studies, with most risk factors found to be both non-significant and significant. Risk factors such as contact with an infected individual (close contact with the individual or their bodily fluids, household contact and non-household contact), participation in funerals (either attendance or involvement in unsafe burial practices), age, occupation, and hospitalisation were most frequently found to be significant. However, sex was more commonly found to be non-significant (n=12 analyses) than significant (n=3).

Risk factors for seropositivity were similar to those investigated for infection and showed comparable conflicting findings across studies (Table S20). Contact with an animal was most commonly found to be significant. Age, sex, occupation, close contact, household and non-household contact, were most frequently non-significant. Participation in funerals was found equally significant and non-significant. Two studies analysed risk factors for onwards transmission; significant risk factors included age, sex, socioeconomic status, survival, funeral, and being part of the first generation of a transmission chain (Table S19).

21 estimates of the serial interval (SI) were reported from 17 studies (n=19 for Zaire species, n=2 for Sudan), and a single generation time estimate was reported (for Zaire, Table 1B and Figure S2). The pooled mean SI estimate for high QA studies (n=6, all Zaire species) was 16.5 days (95% Confidence Interval (CI) 16.1-16.9, I^2^=94%) for common effect and 15.4 days (95% CI 13.2-17.5, I^2^=94%) for random effect (Figure 3A). Estimates including three additional low QA studies were similar (Figure S6).

**Figure 3:**
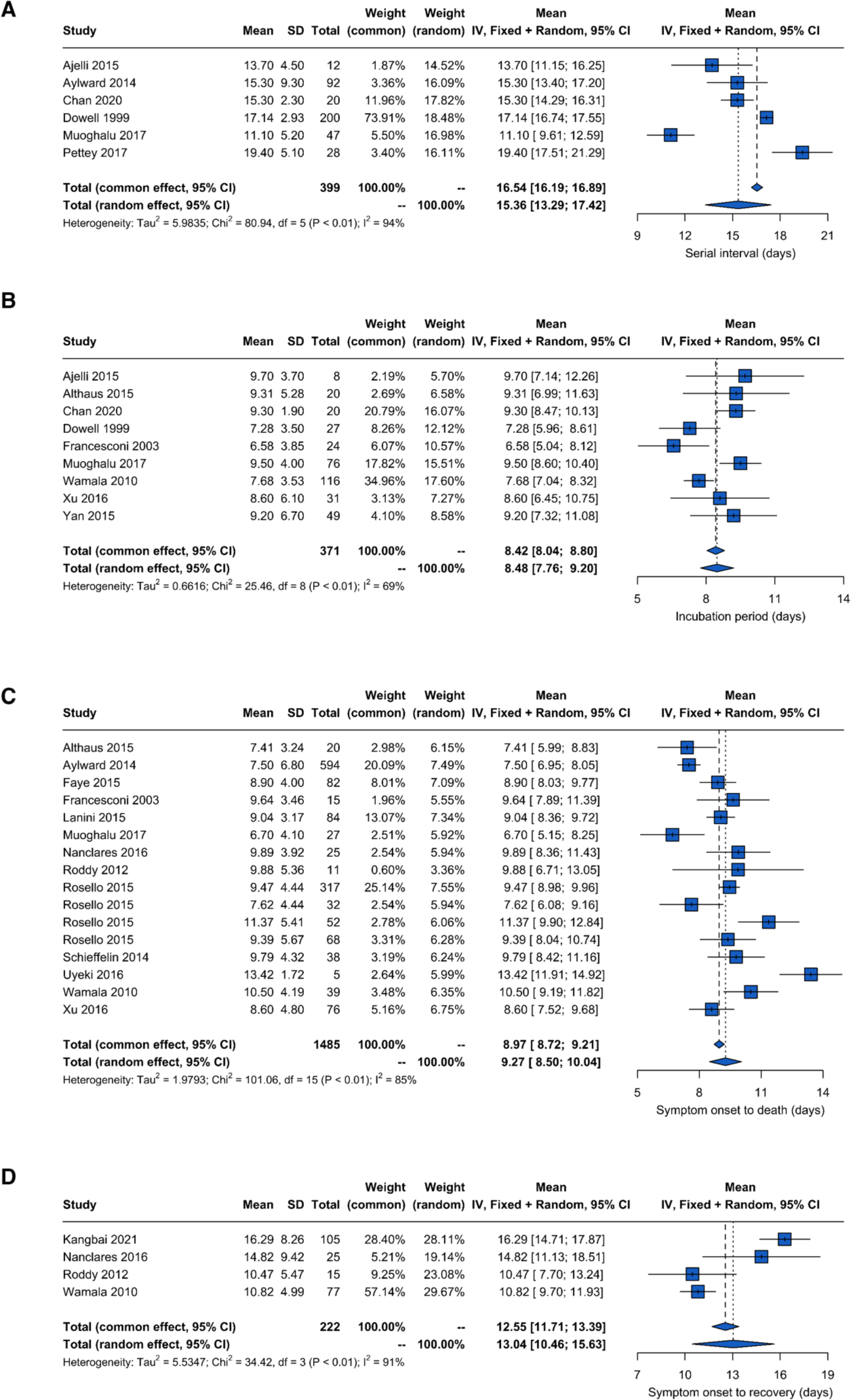
Meta-analysis for the mean A) serial interval, B) incubation period, C) time from symptom onset to death, and D) time from symptom onset to recovery. All included studies have a QA score of >=50%. Parameters used in the meta-analyses are paired mean and standard deviation of the sample or were converted into mean and SD of the sample from the following combinations: mean and standard error, median and interquartile range, or median and range (see SM Section 3.6). Blue squares are mean estimates from each study with 95% confidence intervals. Diamonds represent the overall mean across studies for the common and random effect models. The random effect model accounts for within-study and between-study variance.

We identified 52 estimates for the incubation period from 43 studies, 11 estimates of the latent period from 10 studies, and 27 estimates of the infectious period from 23 studies (Figure S2). Excluding papers with low QA scores (<50%), all estimates were for Ebola Zaire except for 6 estimates of the incubation period (n=3 Bundibugyo and n=3 Sudan). Central estimates ranged from 0.1-31.2 days for the latent period and from 1.7-29.6 days for the infectious period (Table 1B). There were too few high QA latent and infectious period estimates (n=1 for each) in the required format to perform meta-analyses (see Methods and SM Section 3.6). After including all infectious period estimates (regardless of QA score; n=3), the pooled mean was 5.4 days (95% CI 5.3-5.5, I^2^=100%) for common effect and 5.0 days (95% CI 3.7-6.3, I^2^=100%) for random effect (Figure S6). The pooled mean incubation period estimate, based on 9 studies, was 8.4 days (95% CI 8.0-8.8, I^2^ = 69%) for total common effect and 8.5 days (95% CI 7.7-9.2, I^2^ = 69%) for total random effect (Figure 3B. Despite sparse estimates for species other than Zaire in the analysis (n=3), there are statistically significant differences (p=0.02) in mean incubation periods between EV species (Figure S5A).

Clinical progression (e.g. symptom onset to death or recovery) and case management (e.g. hospitalisation) are also key modelling inputs. We extracted estimates for delays from symptom onset to test, test result, diagnosis, reporting (or World Health Organisation (WHO) notification), seeking care, admission to care, quarantine, recovery, negative test or undetectable viral load, discharge from care, and death (Figure S3). We also extracted estimates for delays from admission to care, to discharge, recovery, and/or death (Figure S4). We found delays in the clinical timeline to be highly variable across contexts. Across all studies (including those with low QA), the central estimate of symptom onset to reporting delay varied between 0-25.7 days, although most estimates were around 1-2 weeks (Figure S2). Similarly, the central symptom onset to discharge from care delay varied between 6.3 and 28 days (Figure S3). Time in care was also highly variable irrespective of the outcome (Figure S4) but tended to be shorter for those dying: the central delay from admission to care to death was in the range 0-11 days versus 2.6-17 days for admission to recovery (Figure S4).

In contrast, reported delays from symptom onset to admission to care were remarkably consistent. Central estimates varied between 0 and 23 days for high QA studies (Figure S3), but most (44/47) central estimates fell between 3-6 days. 14 estimates from 12 studies were available for the delay from symptom onset to recovery, with central estimates of those with high QA ranging from 8.4-14.0 days (Table 1B and Figure S3). Based on four estimates from four high QA studies, the pooled mean estimate for the time from symptom onset to recovery was 12.6 days (95% CI 11.7-13.4, I^2^ = 91%) for total common effect and 13.0 days (95% CI 10.4-15.7, I^2^ = 91%) for total random effect (Figure 5). We extracted 48 estimates from 39 studies for the delay from symptom onset to death, with a pooled mean estimate across 16 suitable estimates from 13 high QA studies of 9.0 days (95% CI 8.7-9.2, I^2^ = 85%) for total common effect and 9.3 days (95% CI 8.5-10.1, I^2^ = 85%) for total random effect (Figure 5). Species sub-group meta-analyses using random effects models indicate that the time from symptom onset to death may be longer (p=0.01), and symptom onset to recovery may be shorter (p<0.01), for those infected with the Bundibugyo species compared to Zaire (Figure S5 B&C).

166 estimates of case fatality ratios (CFR) were extracted from 130 papers. Early estimates from the DRC in 1976 and 1995 suggest a CFR of greater than 69% (Table 1C, Figure 4, Table S21). However, more recent estimates from the WA epidemic (2013–16) and DRC (2018–2020) have lower central values but range from 0 to 100% in some settings. Age, sex and occupation were frequently investigated as potential risk factors for death in both multivariate and univariate analyses (Table S22). Age was found to be significantly associated with death in 41/68 parameter entries, whereas most analyses did not find a significant association between death and sex or occupation.

**Figure 4:**
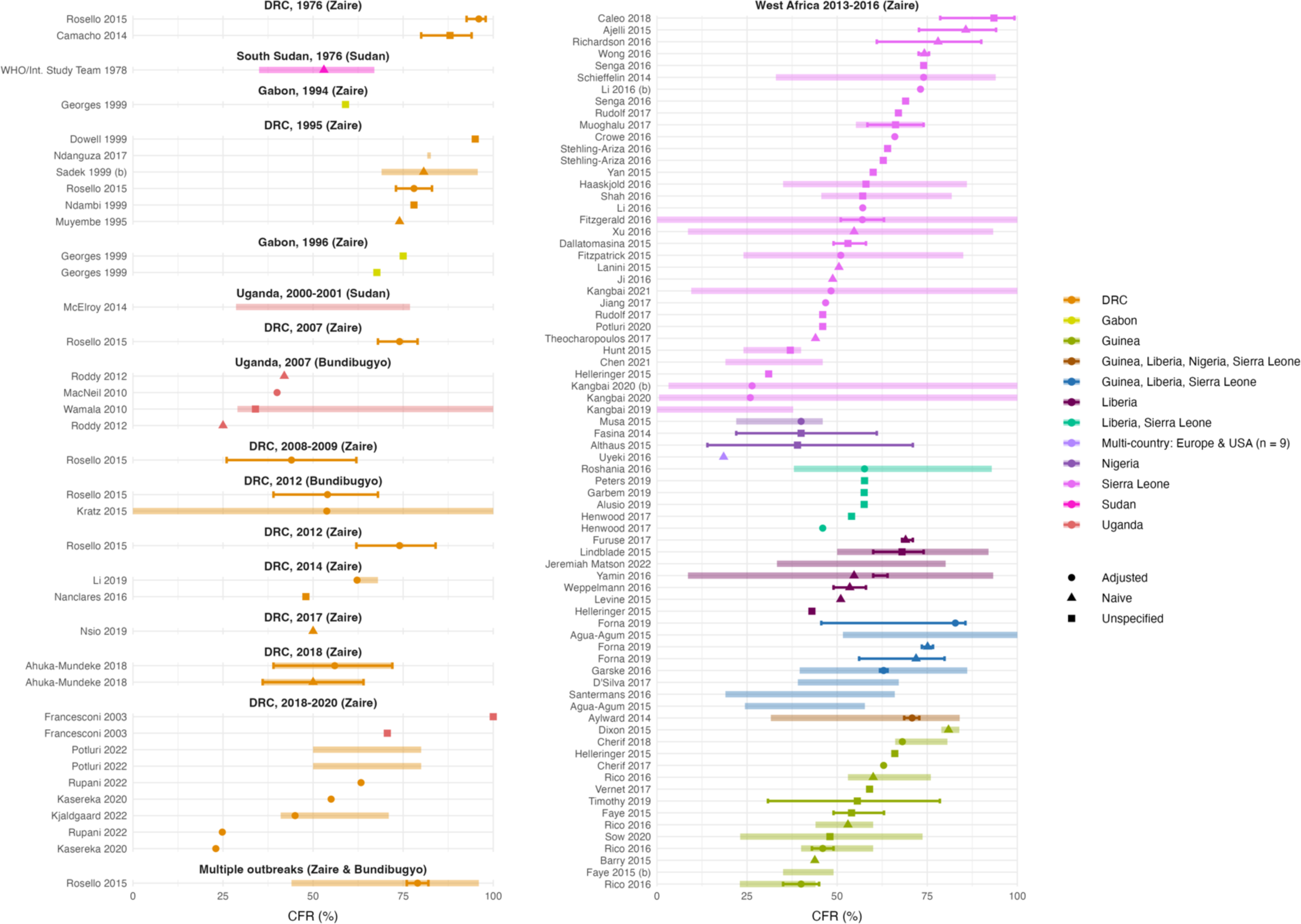
Case Fatality Ratio (CFR) estimates (%) across outbreaks. Each panel corresponds to a different EVD outbreak, with the associated EV species in brackets. Points represent central estimates, with symbol shapes corresponding to analysis type: adjusted, naïve or unspecified. Thick coloured shaded lines represent the range of central estimates when the CFR was estimated, for example, across regions or over time. Solid coloured bars represent the uncertainty around the central estimates, reported in different formats including 95% CrIs and 95% CIs. All parameters are from articles with QA scores of >= 50% (see Table S21 for all CFR estimates).

We extracted 24 estimates of mutation, substitution and evolutionary rates from 20 studies (Table S23). Central estimates ranged from 0.36 × 10^−4^-36 × 10^−4^ substitutions per site per year. The upper bound was an estimate from a single outbreak in DRC (32). In contrast, the same study estimated a much lower substitution rate of 6.9 × 10^−4^ across multiple countries and outbreaks from 1976-2018. Apart from three estimates for all species (Bundibugyo, Sudan, Zaire and Taï forest) (33,34) and a single estimate for Bundibugyo alone (35), all estimates were for Zaire (one study did not specify the species).

Finally, we extracted existing models. Most of the published EVD models were compartmental (210/295), followed by branching process (19/295) and agent-based models (17/295) (Table S24). There were also 49 other model types or combinations of models. Various assumptions were made in the models including homogenous mixing, heterogeneity in transmission rates between groups or over time and the latent period being the same as the incubation period (Table S25). Despite this wealth of knowledge, only 13% of models (n=37) have any publicly available code associated with them, limiting re-usability.

## Discussion

This systematic review presents a comprehensive set of epidemiological parameters and mathematical models for EVD. Most estimates of epidemiological delays were highly variable across studies, which agrees with previous reviews (18,19). This is likely driven by differences in epidemic context, with time to hospitalisation often being used as a marker of response performance (13,14). We found significant differences between species for incubation periods and time from symptom onset to recovery and death, although there was much more evidence for Zaire than for other species. We note the frequent inconsistency in the definition of endpoints, for example reporting versus WHO notification, or recovery versus testing negative, made comparison of epidemiological delays across studies challenging.

Despite the plethora of published evidence, we chose not to summarise all parameters through meta-analysis because there were too few estimates or the definitions and contexts across which parameters have been estimated varied. For example, seroprevalence estimates varied dramatically depending on the population groups being sampled. Seropositivity was very high when looking at serology in people who had past infections e.g. (36) compared to the general population e.g. (37).

Differences in R_0_ values may be driven by more than just the epidemic context e.g. some of the early epidemics with high R_0_ estimates were driven mostly by nosocomial transmission (38,39) or the methodological approach taken differed (e.g. branching processes e.g. (13) or using phylogenies e.g (40)). Uncertainty in R_0_ is especially high in the Nigeria WA epidemic context, likely because it was a small outbreak (41). Estimates of seroprevalence (22,24,27)secondary attack rates (17), and R_0_ (18)are in line with previous reviews, although we did not focus on specific settings so there is greater variability.

CFR estimates also varied greatly between and within outbreaks. This could be due to differences in both the resilience of healthcare systems, patient demographics and conflict (13,42,43). The range in our estimates are consistent with these previous reviews (18,24–27). However, there is insufficient evidence to distinguish whether one or more species have an inherently higher severity, or whether the observed differences are driven by contextual factors such as improved case management over time.

We also extracted parameters not considered in previous systematic reviews. We found substantial but uncertain levels of overdispersion ranging from 0.02 to 2.2. Evidence from Lloyd-Smith et al. (44) would suggest that this corresponds to between ∼30-90% of transmission being attributed to the most infectious 20% of individuals, which is less over dispersive than Severe Acute Respiratory Syndrome (SARS) but highly uncertain. The relatively high mutation rate of the Ebola virus confirms that genomic data may be an important asset to characterise the transmission dynamics in future outbreaks e.g. by reconstructing who infected whom (45). However, like most parameters there is high variability and uncertainty, and most evolutionary estimates are for the Zaire species and the WA epidemic context. Some of the variability in evolutionary rates are likely due to differences in substitution rates between outbreaks compared to within outbreaks(46).

Our study has several limitations. First, due to the extensive literature published on EVD, we restricted our review to published peer-reviewed studies in English. Second, although we attempted to ensure consistency in data extraction by double extracting 55 of the 522 papers, inconsistencies across extractors or incompleteness of data extraction from studies with multiple parameters are possible. Additionally, due to the subjectivity of quality assessment, and the scoring of papers as a whole rather than by individual parameter, consensus among reviewers was difficult to achieve. Third, substantial heterogeneity in the way estimates were reported sometimes made direct comparisons between studies challenging. Often, studies did not distinguish whether uncertainty pertained to the sample or the sample mean, which impacted our ability to include them in meta-analyses. Fourth, specificity of the different seroprevalence tests was often not mentioned and historic papers did not always specify the assay used, making comparisons challenging. Fifth, the large range of reported evolutionary rates may reflect our data extraction method; in particular we did not differentiate between studies using samples solely from humans and those including some samples from animals, nor the method used for estimation or sampling. Finally, we did not extract odds ratios characterising risk factors, nor the direction of protection or risk, and acknowledge that significance classification is somewhat arbitrary and dependent on study design. We encourage readers specifically interested in risk factors to investigate these papers further, for example by accessing the data from this review through the epireview R package (29).

Being prepared at the onset of an infectious disease outbreak is imperative to mount a rapid and effective response to combat the spread of disease. Here we present, synthesise, and analyse the breadth of evidence on the transmissibility, severity, delays, risk factors, mutation rates and seroprevalence of Ebola virus, as well as identify transmission models for EVD, expanding on previous modelling reviews (20,23). We curated 1,280 parameter estimates and 295 model descriptions from 522 studies and make our data available through an easy-to-use R package, epireview (29). We expect that this comprehensive repository will serve as an important resource for modellers and public health community, who can also add to this dynamic database as and when more evidence becomes available, ensuring that this database provides a live picture of the latest evidence on EVD. Much is already known about the Ebola Zaire species; however, our review highlights a critical lack of evidence for other species such as Sudan, Bundibugyo and Taï forest, which is important given that the most recent EVD outbreak was of the Sudan species. Initial analyses suggest statistically significant differences in key parameters between species such as the incubation period and delays from onset to death or recovery. Finally, we note the paucity of publicly available source code for EVD models; publicly releasing the source code for future models will increase usability of existing models in real-time settings.

## Data Availability

Availability of data and materials https://github.com/mrc-ide/epireview/tree/main/data
Code availability https://github.com/mrc-ide/epireview, https://github.com/mrc-ide/priority-pathogens

## Declarations

### Availability of data and materials

https://github.com/mrc-ide/epireview/tree/main/data

### Code availability

https://github.com/mrc-ide/epireview, https://github.com/mrc-ide/priority-pathogens

### PROSPERO

CRD42023393345

(https://www.crd.york.ac.uk/prospero/display_record.php?RecordID&RecordID=393345)

### Authors’ contributions

SB, SvE, NI-E and AC, conceptualised this systematic review. RKN, SB, CM, DJ, AF, SvE, AC and HJTU searched the literature and screened the titles and abstracts. RKN, SB, CM, PD, DJ, KM, RM, AF, GC-D, JTH, TN, IR, AC and HJTU reviewed all full-text articles. RKN, SB, CM, PD, KM, RM, DN, GC-D, JTH, RS, TN, SvE, CG, TR, SIL, JW, KF, AC and HJTU extracted the data. RKN, SB and HJTU did formal analysis of, visualised, and validated the data. RKN and SB were responsible for software infrastructure. AC acquired funding. RKN, SB, GC-D, SvE, NI-E, AC and HJTU were responsible for project administration. RKN, PD, GC-D, DN, SvE and HJTU were responsible for training individuals on and accessing Covidence and designing the Access system. HJTU supervised the systematic review. RKN, SB, AC, and HJTU wrote the original draft of the manuscript. All authors were responsible for the methodology and review and editing of the manuscript. All authors debated, discussed, edited, and approved the final version of the manuscript. All authors had final responsibility for the decision to submit the manuscript for publication.

