## Supplementary material for "Ebola Virus Disease mathematical models and epidemiological parameters: a systematic review and meta-analysis"

**5 Pathogen Epidemiology Review Group (PERG) membership** **51**

### List of Figures

S1 QA scores (%) across parameters. 9
S2 Time periods for the infection process 30
S3 All delays from symptom onset to various endpoints 31
S4 Delays beyond admission to care 32
S5 Meta-analysis for A) incubation period B) onset-to-death and C) onset-to-recovery regardless of QA score, with a species sub-group analysis 34
S6 Meta-analysis for A) infectious period and B) serial interval regardless of QA score 35

### List of Tables

S1 Outbreaks of Ebola Virus Disease 4
S2 Inclusion and exclusion criteria 6
S3 Review papers used in backwards citation screening 6
S4 Quality assessment questionnaire 8
S5 PRISMA 2020 Abstracts Checklist 10
S6 PRISMA 2020 Checklist. 11
S7 Overview of the total number of parameters extracted within each parameter group 13
S8 Overview of the total number of parameters extracted for each parameter type 14
S9 Seroprevalence in countries with history of officially reported EVD cases 15
S10 Seroprevalence in countries with no history of officially reported EVD cases and multi-country analyses 16
S11 Risk factors for seropositivity 17
S12 Attack rates 18
S13 Basic R estimates 20
S14 Effective R estimates 23
S15 Secondary attack rates 25
S16 Growth rates 26
S17 Doubling times 27
S18 Overdispersion table 28
S19 Risk factors for infection table 29
S20 Risk factors for onward transmission by significance and adjustment. 29
S21 Case fatality ratio results 37
S22 Risk factors associated with severity 41
S23 Mutation rates 42
S24 Summary of EVD model types. 43
S25 Summary of EVD model assumptions 43
S26 Models overview 44
S27 Pathogen Epidemiology Review Group (PERG) membership 51

### 1 Overview

In this document, we present additional details and results from our systematic review of mathematical models and epidemiological parameters of Ebola Virus Disease. We first list the 38 known outbreaks of Ebola Virus in Section 2. We then present details of the search terms, inclusion and exclusion criteria, data extraction process, and quality assessment strategy (Section 3). Section 4.1 gives an overview of the epidemiological parameters extracted. Subsequent sections (Sections 4.2 to 4.6) detail results on specific parameters, including summaries across all studies. Transmission models identified in our review are described in Section 4.7.

### 2 Ebola outbreaks

Table S1: Outbreaks of Ebola Virus Disease up to January 2023. This table has been adapted from [1] * = laboratory-acquired infection, ** = imported infection, ± = now South Sudan, ⋄ = cases reported in Guinea, Liberia, Sierra Leone, Mali, Nigeria, Senegal, Italy, Spain, UK & USA.

| Year | Country | Location | Species |
|---|---|---|---|
| 1976 | DRC | Mongala Province | Zaire |
| 1976 | South Sudan | Western and Central Equatoria State | Sudan |
| 1976 | United Kingdom* | Wiltshire, England | Sudan |
| 1977 | DRC | Sud-Ubangi Province | Zaire |
| 1979 | South Sudan | Western Equatoria State | Sudan |
| 1994 | Gabon | Ogooué-Ivindo Province | Zaire |
| 1994 | Côte d'Ivoire | Taï National Park | Tai Forest |
| 1995 | DRC | Kwilu Province | Zaire |
| 1996 | Russia* | Unknown | Zaire |
| 1996(a) | Gabon | Ogooué-Ivindo Province | Zaire |
| 1996(b) | Gabon | Ogooué-Ivindo Province | Zaire |
| 1996 | South Africa** | Johannesburg | Zaire |
| 2000-2001 | Uganda | Northern and Western regions | Sudan |
| 2001-2002 | Gabon & Republic of the Congo | La Zadié, Ivindo and Mpassa Districts (Gabon) and Cuvette-Ouest Department (Republic of the Congo) | Zaire |
| 2003(a) | Republic of the Congo | Cuvette-Ouest Department | Zaire |
| 2003(b) | Republic of the Congo | Cuvette-Ouest Department | Zaire |
| 2004 | Sudan± | Western Equatoria State | Sudan |
| 2004 | Russia* | Unknown | Zaire |
| 2005 | DRC | Cuvette-Ouest Department | Zaire |
| 2007 | DRC | Kasai (formerly Kasaï-Occidental) | Zaire |
| 2007 | Uganda | Western Region | Bundibugyo |
| 2008-2009 | DRC | Kasai (formerly Kasaï-Occidental) | Zaire |
| 2011 | Uganda | Central Region | Sudan |
| 2012 | Uganda | Western Region | Sudan |
| 2012 | DRC | Haut-Uélé Province (formerly Orientale) | Bundibugyo |
| 2012-2013 | Uganda | Central Region | Sudan |

| | | | |
|---|---|---|---|
| 2013-2016 | West-Africa$^{\diamond}$ | | Zaire |
| 2014(b) | DRC | Équateur Province | Zaire |
| 2017 | DRC | Bas-Uélé Province | Zaire |
| 2018 | DRC | Équateur Province | Zaire |
| 2018-2020 | DRC (& Uganda) | North Kivu, Ituri and South Kivu Provinces | Zaire |
| 2020 | DRC | Équateur Province | Zaire |
| 2021(a) | DRC | North Kivu Province | Zaire |
| 2021 | Guinea | Nzérékoré Region | Zaire |
| 2021(b) | DRC | North Kivu Province | Zaire |
| 2022(a) | DRC | Équateur Province | Zaire |
| 2022(b) | DRC | North Kivu Province | Zaire |
| 2022-2023 | Uganda | Central, Eastern and Western regions | Sudan |

### 3 Additional Methods

#### 3.1 Study Selection

We searched PubMed and Web of Science initially on 8$^{\text{th}}$ March 2019 and then updated our search on 7$^{\text{th}}$ July 2023 using the following search terms: Ebola AND ((transmissi* OR epidemiolog*) OR (model* NOT imag*) OR (severity OR "case fatality ratio*" OR CFR OR "case fatality rate*" OR "mortality rate*" OR "attack rate*") OR ("infectious period*" OR "serial interval*" OR "incubation period*" OR "generation time*" OR "generation interval*" OR "latent period*" OR latency) OR (heterogeneit* OR superspread* OR "super spread*" OR super-spread* OR overdispersion OR overdispersed OR over-dispersion OR over-dispersed OR "over dispersion" OR "over dispersed") OR (infectivity OR infectiousness OR "growth rate*" OR "reproduction number*" OR "reproductive number*" OR R0 OR "reproduction ratio*" OR "reproductive rate*") OR ("pre-existing immunity" OR serological OR serology OR serosurvey*) OR (evolution* OR mutation* OR substitution*) OR (outbreak* OR cluster* OR epidemic*) OR ("risk factor*")).

We used the online platform Covidence to perform title screening, abstract screening, and full text review. Our inclusion and exclusion criteria are listed in full in Table S2. Each paper was screened by two reviewers, with disagreements resolved by consensus. No reason was recorded for exclusion at the title and abstract stage. At the full text screening stage, a reason for exclusion was selected from the exclusion criteria.

#### 3.2 Inclusion and exclusion criteria

Following our PROSPERO registration CRD42023393345, our inclusion / exclusion criteria are given in SM Table S2. We did not extract outbreak size information since the Ebola outbreaks are well documented (see Table S1 for a full list [1]). We excluded correspondence, letters to editors with the exception of Nature Letters, which published short articles reporting primary research and conference proceedings, unless they were known to be peer-reviewed.

We excluded reviews at the full-text extraction stage but checked the cited literature to ensure that we had included all relevant papers.

#### 3.3 Backwards citation screening

We identified 179 reviews during our screening process and chose to check we had included all the references from the 11 papers with systematic or meta-analysis in their title that were on relevant topics. We also included one additional review [2] which we knew was highly cited for modelling parameters.

Table S2: Inclusion and exclusion criteria

| **Inclusion** | **Exclusion** |
|---|---|
| Measures/estimates of human: Reproduction numbers (R, $R_0$, $R_t$, r, $R_e$), growth rate (r), doubling times, generation time, serial interval, incubation/latent period, case fatality ratio (CFR), attack rate, mutation rate (e.g. from phylogenetic study), overdispersion, risk factors (risk and the measure). | Non-English language publication |
| Mathematical or statistical model of transmission. | Studies of co-infections. (local, regional, national, international). |
| Measures of seroprevalence and negative seroprevalence in humans. | Animal studies. |
| Nature letters. | Qualitative studies, e.g., KAP studies. |
| | Pathogen not the primary focus of study. |
| | Duplicates. |
| | Does not match any of the inclusion criteria. |
| | In-vitro studies. |
| | Non-peer reviewed publications, conference proceedings, abstracts, posters, letters to the editor |

Table S3: Review papers used in backwards citation screening

| Parameter | Reviews |
|---|---|
| Case fatality ratios | VanKerkhove2015 [2], Rojek2019 [3], Kawuki2021 [4], Nyakarahuka2016 [5], Belhadi2022 [6] |
| Risk factors | VanKerkhove2015 [2], Brainard2016 [7], Selveraj2018 [8] |
| Incubation period | VanKerkhove2015 [2], Velasquez 2015 [9] |
| Latent period | Velasquez2015 [9] |
| Serial interval | VanKerkhove2015 [2] |
| Other delays | VanKerkhove2015 [2] |
| Reproduction number | VanKerkhove2015 [2] |
| Secondary attack rates | Dean2016 [10] |
| Seroprevalence | Bower2017 [11], Nyakarahuka2016 [5] and Rojek2019 [3] |
| Modelling reviews | Wong2017 [12], Abdalla2022 [13] |

#### 3.4 Data extraction

We extracted the data using a bespoke Microsoft Access (Version 2305) database. The extractors extracted data using three forms that then populated the data tables. The exact questions asked in the form and full list of parameters extracted can be found in the Wiki of out GitHub repository for our package (https://github.com/mrc-ide/epireview).

##### 3.4.1 Article

We extracted meta information about the paper including the first author, title, journal, digital object identifier (DOI) and page numbers where available to identify the papers. Each article was scored using seven questions about the quality or risk of bias of the paper (see SM Section 3.5).

###### 3.4.2 Parameters

Our extraction focused on parameters related to seroprevalence, transmission, natural history, severity and evolution. We extracted the following information for each category:

- Seroprevalence
  - seroprevalence in countries with and without reported outbreaks,
  - risk factors for seropositivity.
- Transmission
  - the basic and effective reproduction numbers,
  - growth rate,
  - doubling time,
  - attack rate,
  - secondary attack rate (defined as the probability that an infection occurs among susceptible people within a specific group for example household),
  - overdispersion and
  - risk factors associated for transmission.
- Natural history
  - time periods for the infection process including the serial interval (defined as time between symptoms onset in the infector to symptoms onset in the infectee), generation time (defined as time between infection in the infector and infectee) and incubation period (defined as the time between infection and symptoms onset in an individual),
  - delays beyond symptom onset and
  - delays beyond admission to care.
- Severity
  - case fatality ratios (CFRs) and
  - risk factors associated with severity.
- Evolution
  - evolutionary rates,
  - mutation rates and
  - substitution rates.

We extracted the parameter value type (e.g., mean, standard deviation, median) and uncertainty measure (capturing the precision of estimates) for each parameter when available. If more than three estimates for a single parameter type were presented in a paper, e.g. from different populations, using different methods, or across different time periods, these estimates were extracted as a range of central estimates. Contextual information about the study location and dates, sample size, basic demographic information, and timing of the survey in relation to reported outbreaks were also extracted.

We recorded the methods used for reproduction number estimation (e.g. renewal equations, empirical methods or next generation matrices). For CFRs, we extracted whether the estimation approach was naive or adjusted and recorded numerators and denominators for the CFR and seroprevalence estimates where available. For genomic data, we noted the gene studied if specified and whether the sequence data were available.

We did not extract odds ratio estimates for risk factors because it would be difficult to compare them due to differences in stratifications or reference groups. Instead, we chose to extract an overview of the risk factors explored across studies. This included the outcome of interest e.g. death, the risk factor for that outcome e.g. sex, whether the estimates were statistically significant and whether they were from univariate or multivariate analyses.

###### 3.4.3 Model

We extracted details of Ebola transmission models that were either fit to data or theoretical only. This included the type of model, transmission routes, whether the model was deterministic or stochastic, assumptions, interventions, the availability of model code, and in the case of a compartmental model, model subclassification (e.g., SIR, SEIR).

##### 3.5 Quality Assurance (QA)

Seven questions were used to assess the quality of each paper and the risk of bias, as detailed in SM Table S4. These questions were designed specifically for our review as we found that existing quality assessment criteria did not align with our research given that we were extracting data for both models and various epidemiological parameters. Each question was scored yes (1), no (0) or NA (-) with a total score for the paper assigned as the number of questions answered 'yes' divided by the number of questions answered 'yes' or 'no'. Further information about how these questions were interpreted for each parameter can be found in the Wiki of our GitHub repository for our package (https://github.com/mrc-ide/epireview).

Table S4: Quality assessment questionnaire

| Theme | Question |
|---|---|
| Is the methodological/statistical approach suitable? (how the data are used) | 1. Clear and reproducible |
| | 2. Robust and appropriate for the aim [subjective criteria] |
| Are the assumptions appropriate? (input parameters/assumptions - what goes into the methodology) | 3. Clear and reproducible |
| | 4. Justified (published study or analysis of data)[objective criteria] |
| Are the data appropriate for the selected methodological approach? | 5. Clearly described and reproducible |
| | 6. Are issues in the data clearly discussed and acknowledged? |
| | 7. Are issues in the data accounted for in the chosen methodological approach? |

We observed considerable variability in the QA scores across the extracted parameters (Figure S1). Among 11 included articles published before 1991, only one had a QA score surpassing 50%. Among the 43 included articles published between 1991 and 2014, no discernible trend in QA scores was observed. However, a surge in articles related to EVD emerged after 2014, coinciding with the West African Ebola Epidemic. Of the 522 articles included in our review, 468 were published after 2014. 36% of these articles had a QA score below 50%.

Given the volume of and variability in the data, we aimed to focus on estimates from high quality articles. Therefore, in the figures and tables presented in the main text, we only included parameters from articles with a QA score of at least 50% (which encompasses just below three-quarters (73%) of all articles). The threshold value was selected to balance the inclusion of as much data as possible with the robustness of our analysis, aiming to minimise the influence of unreliable estimates on the results.

A sensitivity analysis for the meta-analyses showed that the inclusion of parameters from low QA scoring papers produced similar results. All parameters are included in the tables and figures in the Supplementary Material.

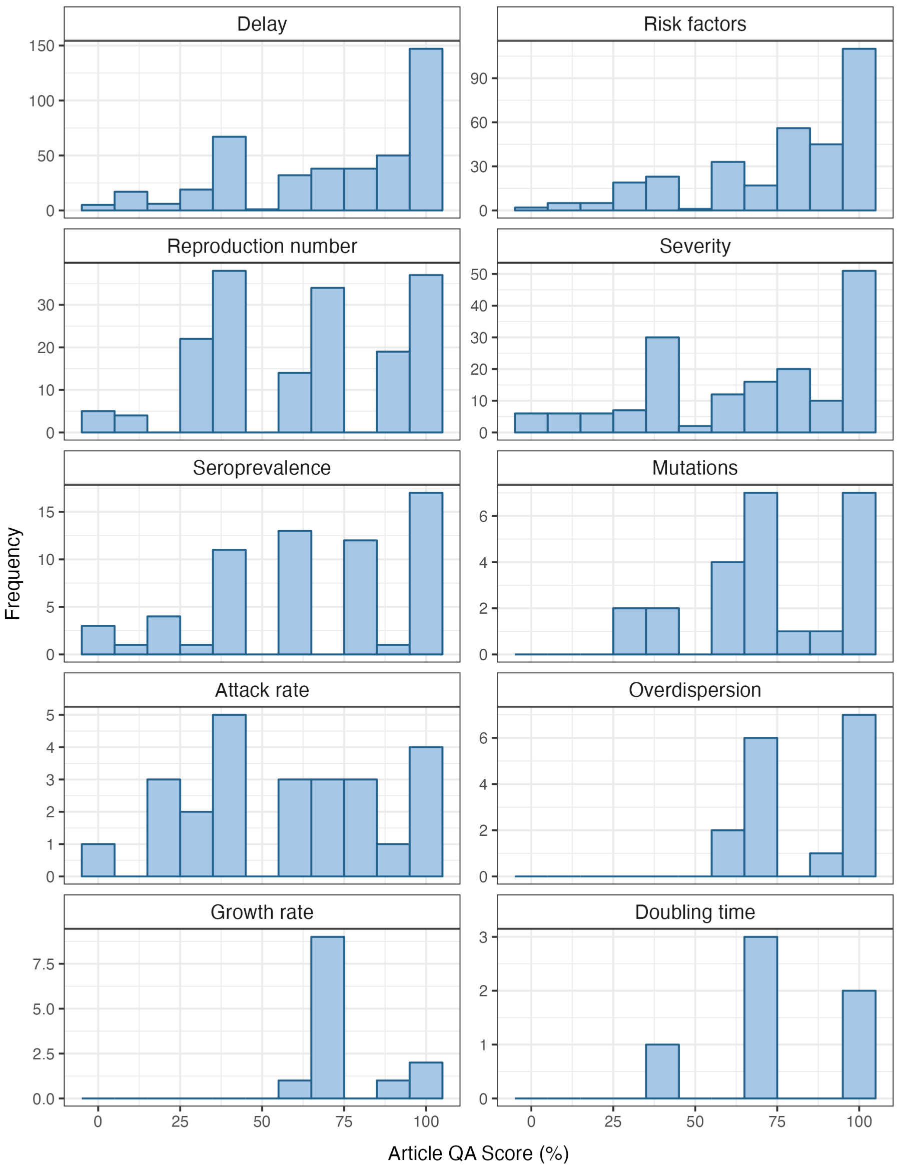

Figure S1: QA scores (%) across parameters.

##### 3.6 Data analysis

All analysis was performed in R using orderly2 for workflow management (https://github.com/mrc-ide/priority-pathogens) and epireview for review specific functions and data storage.

We did not extract information on EVD outbreaks as they are well documented; instead we assigned an outbreak to each extracted parameter based on the country and time for which the parameter had been estimated.

For meta-analyses, we used the metamean function from the meta R package [14]. Parameters were only included in meta-analyses if studies reported a sample size alongside paired (i) parameter means with a standard deviation (SD) of the sample, (ii) means with standard errors, or (iii) medians with a range or interquartile range. For (ii) we converted the standard error to SD using the sample size [15]. For (iii) we used the functionality of metamean to derive the mean and SD using the method outlined by Cai et al. [16], which allows for unknown non-normal distributions. Where data for multiple EV species were available in this format, we used metamean to perform meta-analyses with species sub-groups with p-values being calculated using a $\chi^2$ test.

##### 3.7 PRISMA checklist

Table S5: PRISMA 2020 Abstracts Checklist

| Section & Topic | Item # | Checklist item | Reported (Yes/No) |
|---|---|---|---|
| **Title** | | | |
| Title | 1 | Identify the report as a systematic review. | Yes |
| **Background** | | | |
| Objectives | 2 | Provide an explicit statement of the main objective(s) or question(s) the review addresses. | Yes |
| **Methods** | | | |
| Eligibility criteria | 3 | Specify the inclusion and exclusion criteria for the review. | Yes |
| Information sources | 4 | Specify the information sources (e.g. databases, registers) used to identify studies and the date when each was last searched. | Yes |
| Risk of bias | 5 | Specify the methods used to assess risk of bias in the included studies. | Yes |
| Synthesis of results | 6 | Specify the methods used to present and synthesise results. | Yes |
| **Results** | | | |
| Included studies | 7 | Give the total number of included studies and participants and summarise relevant characteristics of studies. | Yes |
| Synthesis of results | 8 | Present results for main outcomes, preferably indicating the number of included studies and participants for each. If meta-analysis was done, report the summary estimate and confidence/credible interval. If comparing groups, indicate the direction of the effect (i.e. which group is favoured). | Yes |
| **Discussion** | | | |
| Limitations of evidence | 9 | Provide a brief summary of the limitations of the evidence included in the review (e.g. study risk of bias, inconsistency and imprecision). | Yes |
| Interpretation | 10 | Provide a general interpretation of the results and important implications. | Yes |
| **Other** | | | |
| Funding | 11 | Specify the primary source of funding for the review. | Yes |
| Registration | 12 | Provide the register name and registration number. | Yes |

Table S6: PRISMA 2020 Checklist.

| Section & Topic | Item # | Checklist item | Location reported |
|---|---|---|---|
| **Title** | | | |
| Title | 1 | Identify the report as a systematic review. | page 1 |
| **Abstract** | | | |
| Abstract | 2 | See the PRISMA 2020 for Abstracts checklist. | Table S5 |
| **Introduction** | | | |
| Rationale | 3 | Describe the rationale for the review in the context of existing knowledge. | page 2 |
| Objectives | 4 | Provide an explicit statement of the objective(s) or question(s) the review addresses. | page 5 |
| **Methods** | | | |
| Eligibility criteria | 5 | Specify the inclusion and exclusion criteria for the review and how studies were grouped for the syntheses. | page 5 |
| Information sources | 6 | Specify all databases, registers, websites, organisations, reference lists and other sources searched or consulted to identify studies. Specify the date when each source was last searched or consulted. | page 5 |
| Search strategy | 7 | Present the full search strategies for all databases, registers and websites, including any filters and limits used. | page 5 + Figure 1 |
| Selection process | 8 | Specify the methods used to decide whether a study met the inclusion criteria of the review, including how many reviewers screened each record and each report retrieved, whether they worked independently, and if applicable, details of automation tools used in the process. | page 5 |
| Data collection process | 9 | Specify the methods used to collect data from reports, including how many reviewers collected data from each report, whether they worked independently, any processes for obtaining or confirming data from study investigators, and if applicable, details of automation tools used in the process. | page 5 |
| Data items | 10a | List and define all outcomes for which data were sought. Specify whether all results that were compatible with each outcome domain in each study were sought (e.g. for all measures, time points, analyses), and if not, the methods used to decide which results to collect. | page 6, SM Section 3.4.2 |
| | 10b | List and define all other variables for which data were sought (e.g. participant and intervention characteristics, funding sources). Describe any assumptions made about any missing or unclear information. | page 6, SM Section 3.4.2 |
| Study risk of bias assessment | 11 | Specify the methods used to assess risk of bias in the included studies, including details of the tool(s) used, how many reviewers assessed each study and whether they worked independently, and if applicable, details of automation tools used in the process. | page 5 |
| Effect measures | 12 | Specify for each outcome the effect measure(s) (e.g. risk ratio, mean difference) used in the synthesis or presentation of results. | page 6 |
| Synthesis methods | 13a | Describe the processes used to decide which studies were eligible for each synthesis (e.g. tabulating the study intervention characteristics and comparing against the planned groups for each synthesis (item #5)). | page 6 |
| | 13b | Describe any methods required to prepare the data for presentation or synthesis, such as handling of missing summary statistics, or data conversions. | page 6 |
| | 13c | Describe any methods used to tabulate or visually display results of individual studies and syntheses. | page 6 |
| | 13d | Describe any methods used to synthesize results and provide a rationale for the choice(s). If meta-analysis was performed, describe the model(s), method(s) to identify the presence and extent of statistical heterogeneity, and software package(s) used. | page 6 |
| | 13e | Describe any methods used to explore possible causes of heterogeneity among study results (e.g. subgroup analysis, meta-regression). | page 6 |
| | 13f | Describe any sensitivity analyses conducted to assess robustness of the synthesized results. | - |
| Reporting bias assessment | 14 | Describe any methods used to assess risk of bias due to missing results in a synthesis (arising from reporting biases). | page 5 |
| Certainty assessment | 15 | Describe any methods used to assess certainty (or confidence) in the body of evidence for an outcome. | page 5 |
| **Results** | | | |

| | | | |
|---|---|---|---|
| Study selection | 16a | Describe the results of the search and selection process, from the number of records identified in the search to the number of studies included in the review, ideally using a flow diagram. | Figure 1 |
| | 16b | Cite studies that might appear to meet the inclusion criteria, but which were excluded, and explain why they were excluded. | Available on request |
| Study characteristics | 17 | Cite each included study and present its characteristics. | SM Two |
| Risk of bias in studies | 18 | Present assessments of risk of bias for each included study. | SM Section 3.5 |
| Results of individual studies | 19 | For all outcomes, present, for each study: (a) summary statistics for each group (where appropriate) and (b) an effect estimate and its precision (e.g. confidence/credible interval), ideally using structured tables or plots. | pages 6-10 |
| Results of syntheses | 20a | For each synthesis, briefly summarise the characteristics and risk of bias among contributing studies. | SM |
| | 20b | Present results of all statistical syntheses conducted. If meta-analysis was done, present for each the summary estimate and its precision (e.g. confidence/credible interval) and measures of statistical heterogeneity. If comparing groups, describe the direction of the effect. | page 8/9 |
| | 20c | Present results of all investigations of possible causes of heterogeneity among study results. | page 6-10 |
| | 20d | Present results of all sensitivity analyses conducted to assess the robustness of the synthesized results. | Figure S5 |
| Reporting biases | 21 | Present assessments of risk of bias due to missing results (arising from reporting biases) for each synthesis assessed. | Figure S5 |
| Certainty of evidence | 22 | Present assessments of certainty (or confidence) in the body of evidence for each outcome assessed. | pages 6-10 |
| **Discussion** | | | |
| Discussion | 23a | Provide a general interpretation of the results in the context of other evidence. | page 20 |
| | 23b | Discuss any limitations of the evidence included in the review. | page 21 |
| | 23c | Discuss any limitations of the review processes used. | page 21 |
| | 23d | Discuss implications of the results for practice, policy, and future research. | page 21-22 |
| **Other Information** | | | |
| Registration and protocol | 24a | Provide registration information for the review, including register name and registration number, or state that the review was not registered. | page 1 |
| | 24b | Indicate where the review protocol can be accessed, or state that a protocol was not prepared. | page 1 |
| | 24c | Describe and explain any amendments to information provided at registration or in the protocol. | n/a |
| Support | 25 | Describe sources of financial or non-financial support for the review, and the role of the funders or sponsors in the review. | page 2 |
| Competing interests | 26 | Declare any competing interests of review authors. | page 22 |
| Availability of data, code and other materials | 27 | Report which of the following are publicly available and where they can be found: template data collection forms; data extracted from included studies; data used for all analyses; analytic code; any other materials used in the review. | page 22 |

### 4 Results

#### 4.1 Overview of all EVD parameters

Table S7: Overview of the total number of parameters extracted within each parameter group (we do not include parameters in this table where data is available only in the figures).

| Parameter Group | Total Parameters |
|---|---|
| Delay | 420 |
| Risk factors | 316 |
| Reproduction number | 173 |
| Severity | 166 |
| Seroprevalence | 63 |
| Attack rate | 25 |
| Mutations | 24 |
| Overdispersion | 16 |
| Growth rate | 13 |
| Doubling time | 6 |

Table S8: Overview of the total number of parameters extracted for each parameter type. * = "Other delay" encompasses the 32 other delay types that were only extracted from a single study.

| Parameter Type | Total Parameters |
|---|---|
| **Delay** | |
| Symptom onset to admission to care | 60 |
| Incubation period | 52 |
| Symptom onset to death | 48 |
| Admission to care to death | 33 |
| Other delay* | 33 |
| Infectious period | 27 |
| Admission to care to death/discharge | 22 |
| Serial interval | 22 |
| Admission to care to discharge from care | 20 |
| Symptom onset to recovery/non-infectiousness | 15 |
| Symptom onset to reporting | 15 |
| Symptom onset to discharge from care | 14 |
| Admission to care to recovery/non-infectiousness | 12 |
| Latent period | 11 |
| Death to burial | 7 |
| Symptom onset to quarantine | 5 |
| Symptom onset to seeking care | 4 |
| Symptom onset to test | 4 |
| Reporting to death | 3 |
| Reporting to discharge from care | 3 |
| Exposure/infection to admission to care | 2 |
| Exposure/infection to death | 2 |
| Funeral start to funeral end | 2 |
| Symptom onset to antibody detection (IgM/IgG) | 2 |
| Symptom onset to other | 2 |
| **Risk factors** | |
| Risk factors | 316 |
| **Reproduction number** | |
| Basic reproduction number (R0) | 118 |
| Effective reproduction number (Re) | 55 |
| **Severity** | |
| Case Fatality Ratio (CFR) | 166 |
| **Seroprevalence** | |
| IgG | 36 |
| IFA | 13 |
| Unspecified | 8 |
| IgM | 6 |
| **Mutations** | |
| Evolutionary rate | 12 |
| Substitution rate | 10 |
| Mutation rate | 2 |
| **Attack rate** | |
| Secondary attack rate | 15 |
| Attack rate | 10 |
| **Overdispersion** | |
| Overdispersion | 16 |
| **Growth rate** | |
| Growth rate (r) | 13 |
| **Doubling time** | |
| Doubling time | 6 |

#### 4.2 Seroprevalence

##### 4.2.1 Seroprevalence in countries with history of officially reported EVD cases

Table S9: Seroprevalence in countries with history of officially reported EVD cases. * = These are not duplicate entries. One entry corresponds to Zaire antigens, and the other corresponds to Sudan antigens.

| Country | Location | Survey date | Central estimate | Central range | Central type | Uncertainty (95% CI) | Population Sample | Sample size | Disaggregated by | Article | QA score (%) |
|---|---|---|---|---|---|---|---|---|---|---|---|
| **Seroprevalence - IFA** | | | | | | | | | | | |
| DRC | Bumba Zone | 1 Sep 1977 - 24 Oct 1977 | | | | | Community based | | Age, Other, Sex, Symptoms | International Commission 1978 | 40.0 |
| DRC | Sud-Ubangi | 1981 - 1985 | 15.00 | | | | Contact based | 188 | Age | Jezek 1999 | 20.0 |
| DRC | Mabalako, Beni, Butembo, Katwa | Unspecified | 2.30 | | | 1.1 - 4 | Hospital based | 488 | Other | Nkuba-Ndaye 2022 | 80.0 |
| Gabon | Makokou | 1994 - Unspecified | 0.00 | | | | Hospital based | | | Georges 1999 | 80.0 |
| Guinea | | 12 May 2016 - 8 Sep 2017 | 4.10 | 3.32 - 20 | | 3.12 - 5.28 | Contact based | 1,390 | Level of exposure, Other, Sex, Symptoms | Diallo 2019 | 100.0 |
| Sudan | Nzara Maridi | Jun 1976 - Nov 1976 | | 19 - 83 | | | Other | | Occupation, Other, Region | WHO/Int. Study Team 1978 | 60.0 |
| Uganda | Karamoja | May 1984 | 3.00 | | | | Community based | 132 | Region | Rodhain 1989* | 20.0 |
| Uganda | Karamoja | May 1984 | 3.00 | | | | Community based | 132 | Region | Rodhain 1989* | 20.0 |
| **Seroprevalence - IgG** | | | | | | | | | | | |
| DRC | Kikwit | May 1995 - Jul 1995 | 9.00 | 4 - 31 | | | Hospital based | 429 | Occupation | Tomori 1999 | 60.0 |
| DRC | Kikwit | 28 Jul 1995 - 16 Aug 1995 | 2.20 | 0 - 11.1 | | | Trade / business based | 414 | Occupation | Busico 1999 | 60.0 |
| DRC | Kikwit | 18 Aug 1995 - 6 Sep 1995 | 9.30 | 0 - 18.1 | Other | | Community based | 161 | Age, Occupation, Other, Region, Sex | Busico 1999 | 60.0 |
| DRC | Watsa region | Aug 2002 | 18.70 | | Mean | 14.4 - 23.5 | Community based | 300 | Sex | Mulangu 2016 | 80.0 |
| DRC | Sankuru district | Aug 2007 - Sep 2007 | 11.00 | | | | Community based | 3,415 | | Mulangu 2018 | 80.0 |
| DRC | Boende, Kabondo-Dianda, Yambuku, North and South Kikwit | Sep 2015 - Aug 2017 | 8.30 | | Other | | Community based | 1,366 | | Bratcher 2021 | 100.0 |
| DRC | Boende health zone | Sep 2015 - Nov 2015 | 28.10 | | | | Hospital based | 565 | | Hoff 2019 | 60.0 |
| DRC | Boende health zone | Sep 2015 - Nov 2015 | 15.80 | | | | Hospital based | 565 | | Hoff 2019 | 60.0 |
| DRC | Boende | Nov 2015 | 22.50 | 0 - 37 | | | Hospital based | 582 | Age, Level of exposure, Other, Sex | Doshi 2022 | 80.0 |
| DRC | North Kivu Province, Rubare, Kiwanja Umoja, Biruma, Ntamugemga, Kalengera | May 2017 - Apr 2018 | 11.03 | | | | Household based | 272 | Region | Goldstein 2020 | 57.1 |
| DRC | Mbandaka | Jun 2018 - Jul 2018 | 4.60 | | | | Hospital based | 539 | Other | Shaffer 2022 | 80.0 |
| DRC | Mbandaka, Equateur Province | 12 Nov 2018 - 16 Nov 2018 | 5.00 | | Other | | Trade / business based | 19 | | Lucas 2020 | 40.0 |
| Gabon | Lastoursville, Makokou, Matadi-Ngouassa, Moukoro, Doussleoussou, Doussala | 1981 - 1997 | 1.20 | 0 - 2.2 | Other | | Community based | 1,147 | Region, Sex | Lahm 2007 | 100.0 |
| Gabon | | 24 Jan 1996 - 4 Feb 1996 | 10.20 | | Mean | | Community based | 235 | Age, Occupation, Sex | Bertherat 1999 | 40.0 |
| Gabon | Mayibout, Mvadi villages | Feb 1996 - Unspecified | | 14.9 - 30 | | | Hospital based | 205 | Region | Georges 1999 | 80.0 |
| Gabon | Ogooue-Ivindo region | Jul 1997 - Oct 1997 | 1.00 | 1 - 1.4 | Mean | 0.5 - 1.9 | Community based | 2,533 | | Heffernan 2005 | 100.0 |
| Gabon | Rural Gabonese populations | Jun 2005 - Sep 2008 | 15.30 | 1.5 - 32.4 | | 14.3 - 16.5 | Community based | 4,349 | Region | Nkoghe 2011 | 60.0 |
| Gabon | | 2005 - 2008 | 15.30 | | | | Population based | 4,349 | Age, Other, Region, Sex | Becquart 2010 | 100.0 |
| Guinea | | Jun 2012 - Oct 2012 | 0.07 | | | | Population based | 1,483 | | Keita 2018 | 100.0 |
| Guinea | | 23 Mar 2015 - 11 Jul 2016 | | 59.41 - 99.8 | | | Hospital based | | Other, Time | Diallo 2021 | 100.0 |
| Guinea, Liberia, Sierra Leone | Lassa Diagnostic Laboratory | 2007 - 2014 | 5.20 | 0 - 10 | | | Other | 672 | Time | O'Hearn 2016 | 40.0 |
| Republic of the Congo | Brazzaville, Pointe-Noire, Cuvette-Ouest | Mar 2011 - Jul 2011 | 2.50 | 1.6 - 4 | | | Community based | 809 | Region | Moyen 2015 | 100.0 |
| Sierra Leone | Bombali District | Feb 2015 - Mar 2015 | | 3.8 - 100 | | | Other | | Other, Region | Mafopa 2017 | 60.0 |
| Sierra Leone | | Mar 2015 - Dec 2015 | | 21.2 - 27.9 | | | Hospital based | 694 | | Liu 2018 | 100.0 |
| Sierra Leone | | 3 Jul 2015 - 10 Sep 2015 | | 0 - 100 | | 0 - 100 | Household based | 486 | Age, Occupation, Other, Sex, Symptoms | Glynn 2017 | 100.0 |
| Sierra Leone | Sukudu | Oct 2015 - Jan 2016 | 22.00 | | | | Housing estate based | 221 | | Kelly 2018 | 85.7 |
| Sierra Leone | Kambia District | 16 Mar 2016 - 29 Jun 2018 | 8.40 | 0 - 27.8 | Other | 7 - 10 | Community based | 1,282 | Level of exposure, Occupation, Other | Manno 2022 | 100.0 |
| Sierra Leone | Kono District | Sep 2016 - Jul 2017 | 9.50 | | | | Household based | 421 | | Kelly 2022 | 40.0 |
| Sierra Leone | Makeni | Mar 2017 | | 40.1 - 97.7 | Other | | Other | 481 | Other | Halfmann 2019 | 40.0 |
| Uganda | Ibanda, Kamwenge, Luweero | Jan 2015 - Feb 2015 | 2.30 | | | | Community based | 724 | Level of exposure | Nyakarahuka 2020 | 100.0 |
| **Seroprevalence - IgM** | | | | | | | | | | | |
| DRC | Kikwit | 28 Jul 1995 - 16 Aug 1995 | 0.00 | | Other | | Trade / business based | 414 | | Busico 1999 | 60.0 |
| Gabon | Minkebe | 27 Dec 1994 | 27.00 | | | | Unspecified | 33 | | Amblard 1997 | 20.0 |
| Gabon | Andock, Minkebe, Mekouka, Mvadi, Mayibout villages | Dec 1994 - Unspecified | | 1.8 - 10 | | | Hospital based | | Region, Time | Georges 1999 | 80.0 |
| Sierra Leone | | Mar 2015 - Dec 2015 | | 2.4 - 11 | | | Hospital based | 694 | | Liu 2018 | 100.0 |
| **Seroprevalence - Unspecified** | | | | | | | | | | | |
| DRC | Tandala | 1978 | 7.00 | 1 - 21 | | | Population based | 1,096 | Age, Region | Heymann 1980 | 28.6 |
| DRC | Boende health zone | Sep 2015 - Nov 2015 | 9.50 | | | | Hospital based | 565 | | Hoff 2019 | 60.0 |
| DRC | Boende health zone | Sep 2015 - Nov 2015 | 2.80 | | | | Hospital based | 565 | | Hoff 2019 | 60.0 |
| Liberia | Bong County | 15 Sep 2014 - 4 Jan 2015 | 42.00 | | | | Hospital based | 382 | Age, Occupation, Other, Region, Sex | Levine 2015 | 100.0 |
| Sierra Leone | Sukundu | Dec 2014 - Jan 2015 | 7.50 | | | | Contact based | 187 | | Richardson 2016 | 80.0 |
| South Sudan | Nzara, Yambio | 22 Sep 1979 - 9 Oct 1979 | 18.00 | 13 - 38 | | | Household based | 106 | Level of exposure | Baron 1983 | 14.3 |

##### 4.2.2 Seroprevalence in countries with no history of officially reported EVD cases

Table S10: Seroprevalence in countries with no history of officially reported EVD cases and multi-country analyses

| Country | Location | Survey date | Central estimate | Central range | Central type | Uncertainty (95% CI) | Population Sample | Sample size | Disaggregated by | Article | QA score (%) |
|---|---|---|---|---|---|---|---|---|---|---|---|
| **Seroprevalence - IFA** | | | | | | | | | | | |
| Cameroon | Moloundou, Lomie, Lolodorf-Bipindi, Pete, Yaounde | 1979 - 1980 | 9.70 | 3.2 - 23.5 | Mean | | Community based | 1,517 | Age, Other, Region | Bouree 1983 | 40.0 |
| Central African Republic | Nola, Ikaumba, Bozo, Bangassou, Bouar, Obo, Mbre, Birao | Unspecified | 21.30 | | | | Population based | 4,295 | Region | Johnson 1993 | 60.0 |
| Djibouti | Randa, Djibouti City | 1987 | 0.00 | | | | Other | 160 | | Salah 1988 | 0.0 |
| Kenya | Nzoia, Laisamis, Masinga, Lodwar, Malinda, Kilifi | Unspecified | 1.42 | | | | Population based | 1,899 | Region | Johnson 1983 | 40.0 |
| Multi-country: Africa (n = 6) | Cameroon, Central African Republic, Chad, Republic of the Congo, Equatorial Guinea, Gabon | Jan 1985 - Jun 1987 | 12.40 | 1.85 - 32.72 | Other | | Hospital based | 5,070 | Region | Gonzalez 1989 | 42.9 |
| **Seroprevalence - IgG** | | | | | | | | | | | |
| Central African Republic | Lobaye district, Belemboke, Nola and Bangassou | Dec 1992 - Nov 1995 | 5.30 | 1.9 - 16.6 | | | Community based | 1,331 | Region, Sex, Time | Paul Gonzalez 2000 | 40.0 |
| Mali | Bamba, Banzana, Soromba | 2015 | | 1.5 - 6.1 | Other | | Other | 600 | Other | Bane 2021 | 40.0 |
| Multi-country: Africa (n = 5) | Cameroon, DRC, Republic of the Congo, Ghana, Uganda | 1997 - 2012 | 2.20 | 0 - 4.4 | | 0.3 - 4.8 | Other | 2,430 | Region | Steffan 2019 | 100.0 |
| Tanzania | | Jun 2018 - Nov 2018 | 1.80 | | | | Other | 500 | Age, Other, Region, Sex, Symptoms | Rugarabamu 2021 | 80.0 |
| United Kingdom | | 16 Dec 2015 - 16 Jun 2016 | 0.75 | | | | Hospital based | 268 | | Houlihan 2017 | 100.0 |
| **Seroprevalence - IgM** | | | | | | | | | | | |
| Tanzania | | Jun 2018 - Nov 2018 | 1.60 | | | | Other | 500 | Age, Other, Region, Sex, Symptoms | Rugarabamu 2021 | 80.0 |
| Tanzania | | Jun 2018 - Nov 2018 | 0.90 | | | | Hospital based | 308 | Age, Region, Sex | Rugarabamu 2022 | 80.0 |
| **Seroprevalence - Unspecified** | | | | | | | | | | | |
| Madagascar | Antananarivo, Mandoto, Andasibe, Tsiroanomandidy, Ampijoroa | Unspecified | 4.50 | | | | Population based | 381 | Region | Mathiot 1989 | 0.0 |
| Madagascar | Antananarivo, Mandoto, Andasibe, Tsiroanomandidy, Ampijoroa | Unspecified | 0.00 | | | | Population based | 381 | Region | Mathiot 1989 | 0.0 |

##### 4.2.3 Risk factors associated with seropositivity

Table S11: Risk factors for seropositivity. Total is the number of parameters extracted for each risk factor for seropositivity.

| Risk Factor for Seropositivity | Significant | | | Not significant | | | Total |
|---|---|---|---|---|---|---|---|
| | Adjusted | Not adjusted | Unspecified | Adjusted | Not adjusted | Unspecified | |
| Age | 2 | 2 | 4 | 3 | 7 | 2 | 20 |
| Sex | 2 | 4 | 2 | 3 | 5 | 3 | 19 |
| Occupation | 1 | 2 | 2 | 1 | 7 | 0 | 13 |
| Contact with animal | 3 | 2 | 1 | 3 | 2 | 0 | 11 |
| Close contact | 1 | 2 | 0 | 1 | 4 | 0 | 8 |
| Household contact | 1 | 1 | 0 | 1 | 1 | 1 | 5 |
| Funeral | 1 | 1 | 0 | 1 | 1 | 0 | 4 |
| Non-household contact | 0 | 0 | 0 | 0 | 1 | 0 | 1 |
| Other | 6 | 3 | 4 | 5 | 8 | 1 | 27 |

#### 4.3 Transmission parameters

##### 4.3.1 Attack rate

Table S12: Attack rates

| Country | Survey date | Central estimate (%) | Central range | Uncertainty (95% CI) | Population Sample | Sample size | Population Group | Disaggregated by | Article | QA score (%) |
|---|---|---|---|---|---|---|---|---|---|---|
| **South Sudan, 1976** | | | | | | | | | | |
| Sudan | Jun 1976 - Nov 1976 | | 0.34 - 1.42 | | Community based | 280 | Persons under investigation | Region | WHO/Int. Study Team 1978 | 60.0 |
| **DRC, 1995** | | | | | | | | | | |
| DRC | May 1995 - Jul 1995 | 9 | 4 - 31 | | Hospital based | 429 | Healthcare workers | Occupation | Tomori 1991 | 100.0 |
| **Uganda, 2000-2001** | | | | | | | | | | |
| Uganda | 2000 | | 0.01 - 0.238 | | Community based | | Persons under investigation | Region | Okware 2015 | 20.0 |
| **Republic of the Congo, 2005** | | | | | | | | | | |
| Republic of the Congo | 18 Apr 2005 - 8 Jul 2005 | 7.1 | | | Community based | | General population | Region | Nkoghe 2011 (b) | 42.9 |
| Republic of the Congo | 18 Apr 2005 - 8 Jul 2005 | 8.6 | | | Community based | | General population | Region | Nkoghe 2011 (b) | 42.9 |
| Republic of the Congo | 18 Apr 2005 - 8 Jul 2005 | 8.7 | | | Community based | | General population | Region | Nkoghe 2011 (b) | 42.9 |
| **Uganda, 2007** | | | | | | | | | | |
| Uganda | 29 Nov 2007 - 20 Feb 2008 | 0.043 | 0.003 - 0.185 | | Community based | 116 | General population | Age, Region, Sex | Wamala 2010 | 80.0 |
| **West Africa 2013-2016** | | | | | | | | | | |
| Guinea | 22 Jun 2017 - 9 Jul 2017 | 7.29 | | 4.38 - 11.28 | Household based | 247 | General population | | Timothy 2019 | 100.0 |
| Liberia | 14 May 2014 - 7 Mar 2015 | 0.25 | | 0.15 - 0.4 | Community based | 17 | General population | | Kuehne 2016 | 28.6 |
| Sierra Leone | May 2014 - Apr 2015 | 2.7 | | | Household based | 1161 | General population | | Caleo 2018 | 100.0 |

###### 4.3.2 Basic reproduction number

Table S13: Basic R estimates

| Country | Survey date | Central estimate | Central range | Central type | Uncertainty | Uncertainty type | Method | Disaggregated by | Article | QA score (%) |
|---|---|---|---|---|---|---|---|---|---|---|
| **DRC, 1976** | | | | | | | | | | |
| DRC | Aug 1976 - Nov 1976 | 4.71 | 1.34 - 4.71 | Median | 3.92 - 5.66 | 95% CI | Compartmental model | Other | Camacho 2014 | 71.4 |
| **DRC, 1995** | | | | | | | | | | |
| DRC | 1 Mar 1995 - 16 Jul 1995 | 1.40 | | Mean | | | Compartmental model | | Lekone 2006 | 100.0 |
| DRC | 1 Mar 1995 - 12 Jul 1995 | 1.80 | | Unspecified | | | Compartmental model | | Zaman 2009 | 100.0 |
| DRC | 1 Mar 1995 - 21 Jul 1995 | 2.11 | | Unspecified | | | Compartmental model | | Ndanguza 2013 | 42.9 |
| DRC | 1 Mar 1995 - 21 Jul 1995 | 2.22 | | Mean | 2.22 - 2.22 | 95% CrI | | | Ndanguza 2013 | 42.9 |
| DRC | 1 Mar 1995 - 21 Jul 1995 | 2.22 | | Mean | 0.13 | Standard Deviation | Compartmental model | | Ndanguza 2013 | 42.9 |
| DRC | 6 Mar 1995 - 12 Jul 1995 | 1.83 | | Median | 0.06 | Standard Deviation | Compartmental model | | Chowell 2004 | 28.6 |
| DRC | Mar 1995 - Jul 1995 | | 1.5 - 2.08 | Mean | | | Compartmental model | Other | Ward 2023 | 42.9 |
| DRC | 1995 | 1.93 | | Median | 1.74 - 2.78 | IQR | Other | | White 2007 | 85.7 |
| DRC | 1995 | 2.70 | | Mean | 1.9 - 2.8 | 95% CI | Compartmental model | | Legrand 2007 | 100.0 |
| DRC | 1995 | 2.98 | | Mean | 2.11 - 4.36 | 95% CrI | Next generation matrix | Occupation | Robert 2019 | 100.0 |
| **Uganda, 2000-2001** | | | | | | | | | | |
| Uganda | 20 Aug 2000 - 7 Jan 2001 | 1.34 | | Median | 0.03 | Standard Deviation | Compartmental model | | Chowell 2004 | 28.6 |
| Uganda | 2000 | 2.70 | | Mean | 2.5 - 4.1 | 95% CI | | | Legrand 2007 | 100.0 |
| **DRC, 2012** | | | | | | | | | | |
| DRC | 2012 | 0.86 | | Mean | 0.37 - 1.6 | 95% CrI | Branching process | | Choi 2019 | 42.9 |
| DRC | 2012 | 1.11 | | Mean | 1.06 - 1.17 | 95% CrI | Other | | Choi 2019 | 42.9 |
| DRC | 2012 | 1.37 | | Mean | 0.85 - 2.02 | 95% CrI | Branching process | | Choi 2019 | 42.9 |
| **West Africa 2013-2016** | | | | | | | | | | |
| Guinea | Dec 2013 - Mar 2016 | 0.89 | | Mean | | | Empirical (contact tracing) | Disease generation | Robert 2019 (b) | 100.0 |
| Guinea | 1 Jan 2014 - Dec 2014 | 1.30 | | Unspecified | | | Compartmental model | | Uekermann 2019 | 28.6 |
| Guinea | 1 Mar 2014 - 18 Mar 2015 | 1.12 | | Mean | 1.11 - 1.12 | 95% CI | Compartmental model | | Wang 2015 | 42.9 |
| Guinea | 22 Mar 2014 - 29 Aug 2019 | 1.00 | | Mean | 0.77 - 1.35 | Range | Compartmental model | | Agusto 2015 | 57.1 |
| Guinea | 22 Mar 2014 - 22 Mar 2015 | 1.26 | | Mean | 1.22 - 1.29 | 95% CI | Next generation matrix | | Shen 2015 | 71.4 |
| Guinea | 22 Mar 2014 - 29 Aug 2019 | 1.60 | | Mean | 1.15 - 2.05 | Range | Compartmental model | | Agusto 2015 | 57.1 |
| Guinea | 22 Mar 2014 - 22 Aug 2014 | 2.46 | | Unspecified | | | Compartmental model | | Fisman 2014 (b) | 28.6 |
| Guinea | 22 Mar 2014 - 25 Jan 2015 | 4.16 | | Unspecified | | | Compartmental model | | Li 2015 | 42.9 |
| Guinea | 25 Mar 2014 - 3 May 2015 | 1.21 | | Median | 1.21 - 1.21 | 95% CI | Growth rate | | Liu 2015 | 71.4 |
| Guinea | 25 Mar 2014 - 25 Oct 2014 | 1.49 | | Median | 1.48 - 1.5 | 95% CI | Next generation matrix | | Luo 2019 | 85.7 |
| Guinea | 27 May 2014 - 1 Jun 2015 | | 0.85 - 1.22 | Unspecified | | | Compartmental model | Time | Webb 2016 | 57.1 |
| Guinea | Aug 2014 - May 2015 | 1.18 | | Mean | 1.17 - 1.19 | 95% CI | Next generation matrix | | Ajelli 2016 | 71.4 |
| Guinea | Unspecified - 7 Sep 2014 | 1.24 | | Unspecified | 1.04 - 1.42 | 95% CI | Other | | Taylor 2016 | 71.4 |
| Guinea, Liberia, Nigeria, Sierra Leone | 30 Dec 2013 - 2014 | | 1.2 - 2.02 | Mean | | | Branching process | Region | Aylward 2014 | 100.0 |
| Guinea, Liberia, Nigeria, Sierra Leone | 22 Mar 2014 - 22 Aug 2014 | 1.78 | | Unspecified | | | Compartmental model | | Fisman 2014 (b) | 28.6 |
| Guinea, Liberia, Sierra Leone | 28 Dec 2013 - 3 Oct 2014 | 1.44 | 0.75 - 1.92 | Other | | | Compartmental model | | Barbarossa 2015 | 42.9 |
| Guinea, Liberia, Sierra Leone | 2013 - 11 Nov 2014 | 1.21 | 1.09 - 1.22 | Unspecified | | | Branching process | Region | Evans 2015 | 42.9 |
| Guinea, Liberia, Sierra Leone | 2013 - 2014 | 1.30 | 0.36 - 3.37 | Mean | 0.64 | Standard Deviation | Growth rate | Region | Krauer 2016 | 100.0 |
| Guinea, Liberia, Sierra Leone | 1 Jan 2014 - 28 Sep 2014 | 1.96 | | Unspecified | | | Other | | Pruyt 2015 | 71.4 |
| Guinea, Liberia, Sierra Leone | Jan 2014 - Sep 2015 | | 0.98 - 2.39 | Unspecified | | | Next generation matrix | Other, Region | Lin 2020 | 42.9 |
| Guinea, Liberia, Sierra Leone | 22 Mar 2014 - 20 Aug 2014 | | 1.51 - 2.53 | Unspecified | | | Compartmental model | Region | Althaus 2014 | 42.9 |
| Guinea, Liberia, Sierra Leone | 25 Mar 2014 - 3 May 2015 | 1.35 | | Median | 1.35 - 1.35 | 95% CI | Growth rate | | Liu 2015 | 71.4 |

| Country | Survey date | Central estimate | Central range | Central type | Uncertainty | Uncertainty type | Method | Disaggregated by | Article | QA score (%) |
|---|---|---|---|---|---|---|---|---|---|---|
| Guinea, Liberia, Sierra Leone | Mar 2014 - Dec 2014 | | 1.24 - 1.56 | Unspecified | | | | Region | Diaz 2018 | 85.7 |
| Guinea, Liberia, Sierra Leone | 6 Jul 2014 - 9 Aug 2014 | 1.80 | 1.5 - 2 | Unspecified | | | Compartmental model | | Gomes 2014 | 85.7 |
| Guinea, Liberia, Sierra Leone | 2014 - 2015 | 1.14 | | Unspecified | | | Next generation matrix | | Long 2018 | 0.0 |
| Guinea, Liberia, Sierra Leone | 2014 - 2015 | 1.40 | | Unspecified | | | Next generation matrix | | Denes 2019 | 57.1 |
| Guinea, Liberia, Sierra Leone | 2014 | | 1.4 - 1.9 | Unspecified | | | Compartmental model | Region | Stockdale 2021 | 42.9 |
| Guinea, Liberia, Sierra Leone | Unspecified - 18 Oct 2014 | 1.79 | | Unspecified | 1.78 - 1.81 | 95% CI | Compartmental model | | Fisman 2014 | 28.6 |
| Guinea, Sierra Leone | May 2014 - Jun 2014 | 5.92 | | Mean | 2.97 - 11.12 | HPDI 95% | Genomics | | Saulnier 2017 | 71.4 |
| Liberia | 2013 - Dec 2014 | 2.00 | | Unspecified | | | Next generation matrix | | Njankou 2018 | 57.1 |
| Liberia | 1 Jan 2014 - Apr 2015 | 2.50 | | Unspecified | | | Compartmental model | | Uekermann 2019 | 28.6 |
| Liberia | 1 Mar 2014 - 18 Mar 2015 | 1.23 | | Mean | 1.23 - 1.23 | 95% CI | Compartmental model | | Wang 2015 | 42.9 |
| Liberia | 17 Mar 2014 - Aug 2014 | 2.11 | | Unspecified | 1.88 - 2.71 | Other | Next generation matrix | | Valdez 2015 | 57.1 |
| Liberia | 18 Mar 2014 - 9 Nov 2014 | 1.40 | 0.7 - 2 | Mean | 0.1 - 3.9 | 95% CI | Compartmental model | Time | Funk 2017 | 100.0 |
| Liberia | 18 Mar 2014 - 9 Nov 2014 | 4.09 | | Standard Deviation | 1.47 - 8.35 | 95% CrI | Compartmental model | | Funk 2017 | 100.0 |
| Liberia | 23 Mar 2014 - 5 Oct 2014 | | 0.9 - 4.6 | Median | | | Compartmental model | Time | Enanoria 2015 | 85.7 |
| Liberia | 25 Mar 2014 - 3 May 2015 | 3.02 | | Median | 2.61 - 3.49 | 95% CI | Growth rate | | Liu 2015 | 71.4 |
| Liberia | 27 Mar 2014 - 22 Aug 2014 | 1.72 | | Unspecified | | | Compartmental model | | Fisman 2014 (b) | 28.6 |
| Liberia | 27 Mar 2014 - 25 Oct 2014 | 1.82 | | Median | 1.77 - 1.83 | 95% CI | Next generation matrix | | Luo 2019 | 85.7 |
| Liberia | 1 May 2014 - 1 Oct 2014 | 1.76 | 1.76 - 1.9 | Unspecified | | | Next generation matrix | | Khan 2015 | 28.6 |
| Liberia | 2 Jun 2014 - 2015 | 1.95 | | Unspecified | | | Compartmental model | | Xie 2019 | 57.1 |
| Liberia | 5 Jun 2014 - 14 Sep 2014 | | 1.71 - 1.94 | Unspecified | | | Compartmental model | Other | Ponce 2019 | 42.9 |
| Liberia | 14 Jun 2014 - 15 Dec 2014 | 2.49 | | Unspecified | 2.38 - 2.6 | 95% CI | Next generation matrix | | Lewnard 2014 | 85.7 |
| Liberia | 17 Jun 2014 - 23 Sep 2014 | 1.54 | | Unspecified | | | Compartmental model | | Webb 2015 | 71.4 |
| Liberia | 17 Jun 2014 - 22 Mar 2015 | 1.80 | | Mean | 1.77 - 1.83 | 95% CI | Next generation matrix | | Shen 2015 | 71.4 |
| Liberia | 29 Jun 2014 - 7 Oct 2014 | 2.01 | | Unspecified | | | Next generation matrix | | Xia 2015 | 85.7 |
| Liberia | Jul 2014 - Dec 2014 | 1.70 | | Median | 1.1 - 2.6 | 95% CI | Empirical (contact tracing) | | Lindblade 2015 | 100.0 |
| Liberia | 2014 - 2015 | 1.67 | | Median | 1.29 - 2.24 | 95% CrI | Other | | Tang 2023 | 42.9 |
| Liberia | 2014 | 2.22 | | Unspecified | | | Compartmental model | | Rivers 2014 | 42.9 |
| Liberia | Unspecified - 1 Oct 2014 | | 1.54 - 1.61 | Unspecified | | | Next generation matrix | | Imran 2017 | 42.9 |
| Liberia | Unspecified - 7 Sep 2014 | 2.06 | | Unspecified | 1.93 - 2.27 | 95% CI | Other | | Taylor 2016 | 71.4 |
| Multi-country: Africa, Europe, USA (n = 12) | Jul 2014 - Mar 2015 | 0.50 | 0.05 - 2.9 | Unspecified | 0.2 - 1 | 90% CI | Branching process | Other | Toth 2015 | 71.4 |
| Multi-country: Africa, Europe, USA (n = 8) | 2014 - 2015 | 2.10 | 1.5 - 2.6 | Unspecified | | | Compartmental model | Region | Ivorra 2015 | 42.9 |
| Nigeria | 17 Jul 2014 - 20 Oct 2014 | 3.20 | | Mean | 0.8 - 16.4 | 95% CrI | Branching process | | Chan 2020 | 85.7 |
| Nigeria | 17 Jul 2014 - 20 Oct 2014 | 10.00 | | Mean | 3 - 18.5 | 95% CrI | Branching process | | Chan 2020 | 85.7 |
| Nigeria | 20 Jul 2014 - 1 Oct 2014 | | 0.4 - 12 | Unspecified | | | Empirical (contact tracing) | Disease generation | Fasina 2014 | 71.4 |
| Nigeria | 20 Jul 2014 - 20 Oct 2014 | 9.01 | | Unspecified | 5.22 - 15.55 | 95% CI | Compartmental model | | Althaus 2015 | 71.4 |
| Nigeria | Jul 2014 - Sep 2014 | 2.57 | | Unspecified | | | Next generation matrix | | Do 2016 | 14.3 |
| Sierra Leone | 2013 - Dec 2014 | 2.50 | | Unspecified | | | | | Njankou 2018 | 57.1 |
| Sierra Leone | 1 Jan 2014 - Feb 2015 | 1.50 | | Unspecified | | | Compartmental model | | Uekermann 2019 | 28.6 |
| Sierra Leone | 1 Mar 2014 - 18 Mar 2015 | 1.18 | | Mean | 1.18 - 1.18 | 95% CI | Compartmental model | | Wang 2015 | 42.9 |
| Sierra Leone | 25 Mar 2014 - 3 May 2015 | 1.90 | | Median | 1.86 - 1.95 | 95% CI | Growth rate | | Liu 2015 | 71.4 |
| Sierra Leone | 1 May 2014 - 1 Oct 2014 | 1.49 | 1.36 - 1.49 | Unspecified | | | Next generation matrix | | Khan 2015 | 28.6 |
| Sierra Leone | 12 May 2014 - 13 Nov 2015 | 1.30 | | Unspecified | 1.26 - 2.53 | Range | Other | | Chen 2021 | 71.4 |
| Sierra Leone | 19 May 2014 - 11 Jan 2015 | 1.70 | | Unspecified | | | Compartmental model | Region | Li 2017 (b) | 57.1 |

| Country | Survey date | Central estimate | Central range | Central type | Uncertainty | Uncertainty type | Method | Disaggregated by | Article | QA score (%) |
|---|---|---|---|---|---|---|---|---|---|---|
| Sierra Leone | 19 May 2014 - 11 Jan 2015 | 1.90 | | Unspecified | | | Other | | Li 2020 | 0.0 |
| Sierra Leone | 25 May 2014 - 18 Jun 2014 | 2.18 | 1.65 - 2.18 | Median | 1.24 - 3.55 | HPDI 95% | Genomics | | Stadler 2014 | 100.0 |
| Sierra Leone | 25 May 2014 - 20 Jun 2014 | | 2.1 - 3.81 | Median | | | Compartmental model | | Volz 2014 | 85.7 |
| Sierra Leone | 25 May 2014 - 18 Jan 2015 | | 0.6 - 5.9 | Median | | | Compartmental model | Time | Enanoria 2015 | 85.7 |
| Sierra Leone | 27 May 2014 - 1 Jun 2015 | | 0.72 - 1.77 | Unspecified | | | Compartmental model | Time | Webb 2016 | 57.1 |
| Sierra Leone | 27 May 2014 - 23 Sep 2014 | 1.26 | | Unspecified | | | Compartmental model | | Webb 2015 | 71.4 |
| Sierra Leone | 27 May 2014 - 31 Aug 2014 | 1.29 | 1.29 - 1.47 | Unspecified | 1.27 - 1.37 | 95% CI | Compartmental model | | Scarpino 2015 | 42.9 |
| Sierra Leone | 27 May 2014 - 31 Aug 2014 | 1.40 | | Unspecified | 1.1 - 1.8 | HPDI 95% | Genomics | | Scarpino 2015 | 42.9 |
| Sierra Leone | 27 May 2014 - 22 Mar 2015 | 1.61 | | Mean | 1.56 - 1.66 | 95% CI | Next generation matrix | | Shen 2015 | 71.4 |
| Sierra Leone | 27 May 2014 - 25 Oct 2014 | 1.67 | | Median | 1.59 - 1.76 | 95% CI | Next generation matrix | | Luo 2019 | 85.7 |
| Sierra Leone | 27 May 2014 - 22 Aug 2014 | 8.33 | | Unspecified | | | Compartmental model | | Fisman 2014 (b) | 28.6 |
| Sierra Leone | May 2014 - Jul 2014 | | 1.21 - 1.6 | Median | | | | Other | Duchene 2019 | 57.1 |
| Sierra Leone | May 2014 - May 2015 | | 1.7 - 4.8 | Unspecified | | | Compartmental model | Region | Getz 2019 | 71.4 |
| Sierra Leone | Jul 2014 - Aug 2014 | 1.63 | | Mean | 0 - 4 | Range | Empirical (contact tracing) | | Ajelli 2015 | 71.4 |
| Sierra Leone | Jul 2014 - Nov 2014 | 2.24 | | Unspecified | 1.52 - 4.51 | 95% CI | Growth rate | | Ajelli 2015 | 71.4 |
| Sierra Leone | Sep 2014 - 2 Feb 2015 | | 0.5 - 8.4 | Unspecified | | | Compartmental model | Region | Kucharski 2015 | 100.0 |
| Sierra Leone | 20 Oct 2014 - 30 Mar 2015 | 2.00 | | Unspecified | 1.8 - 2.2 | 95% CI | Other | | Lau 2017 | 85.7 |
| Sierra Leone | 20 Oct 2014 - 30 Mar 2015 | 2.39 | | Median | 2.05 - 2.84 | 95% CrI | Other | | Lau 2017 (b) | 100.0 |
| Sierra Leone | 28 Nov 2014 - Feb 2015 | 2.90 | 2.1 - 4 | Mean | 1.2 - 6.9 | 90% CrI | Compartmental model | Time | Funk 2019 | 100.0 |
| Sierra Leone | 1 Dec 2014 - 28 Feb 2015 | 0.29 | | Mean | 0.11 - 0.55 | | Empirical (contact tracing) | Level of exposure | Stehling-Ariza 2016 | 100.0 |
| Sierra Leone | 1 Dec 2014 - 28 Feb 2015 | 0.93 | | Mean | 0.15 - 2.3 | 95% CI | Empirical (contact tracing) | Level of exposure | Stehling-Ariza 2016 | 100.0 |
| Sierra Leone | 2014 - 2015 | 1.66 | | Median | 1.31 - 2.15 | 95% CrI | Other | | Tang 2023 | 42.9 |
| Sierra Leone | 2014 | 1.78 | | Unspecified | | | Compartmental model | | Rivers 2014 | 42.9 |
| Sierra Leone | Unspecified - 7 Sep 2014 | 1.71 | | Unspecified | 1.4 - 1.82 | 95% CI | Other | | Taylor 2016 | 71.4 |
| Sierra Leone | Unspecified | | 1.81 - 1.84 | Unspecified | | | Next generation matrix | | Imran 2017 | 42.9 |
| Sierra Leone | Unspecified | 2.13 | | Other | | | Next generation matrix | | Abbate 2016 | 85.7 |
| Unspecified | Unspecified - 6 Sep 2014 | | 1.51 - 2.26 | Unspecified | | | Unspecified | | Hunt 2014 | 14.3 |
| **DRC, 2014** | | | | | | | | | | |
| DRC | 26 Jul 2014 - 4 Oct 2014 | 5.15 | | Unspecified | 3.95 - 6.69 | 95% CI | Compartmental model | | Althaus 2015 (b) | 71.4 |
| DRC | 2014 | 0.84 | | Mean | 0.59 - 1.15 | 95% CrI | | | Burch 2017 | 42.9 |
| **DRC, 2018-2020** | | | | | | | | | | |
| DRC | 3 May 2018 - 12 Sep 2019 | | 1.1 - 1.21 | Mean | | | Compartmental model | Time | Vossler 2022 | 42.9 |
| DRC | 5 Aug 2018 - 2 Feb 2020 | 1.08 | | Unspecified | | | Compartmental model | | Potluri 2022 | 71.4 |
| DRC | 5 Aug 2018 - 20 Oct 2019 | 7.73 | | Unspecified | | | Next generation matrix | | Mouanguissa 2021 | 57.1 |
| DRC | Aug 2018 - Sep 2019 | | 0.88 - 1.22 | Median | 0.76 - 1.31 | 95% CI | Branching process | Time | Jombart 2020 | 42.9 |
| DRC | 4 Aug 2019 - 10 Jan 2019 | 1.83 | | Unspecified | | | Compartmental model | | Hart 2019 | 71.4 |
| DRC | 4 Aug 2019 - 10 Jan 2019 | 1.90 | | Unspecified | | | Compartmental model | | Hart 2019 | 71.4 |
| **Uganda, 2022-2023** | | | | | | | | | | |
| Uganda | Aug 2022 - Nov 2022 | | 1.99 - 2.7 | Unspecified | | | | Other | Marziano 2023 | 100.0 |
| **Multiple outbreaks** | | | | | | | | | | |
| DRC, Guinea, Liberia, Sierra Leone | 1995 - 2016 | | 1.53 - 266.72 | Unspecified | | | Compartmental model | Region, Time | Siewe 2020 (b) | 28.6 |

##### 4.3.3 Effective reproduction number

Table S14: Effective R estimates

| Country | Survey date | Central estimate | Central range | Central type | Uncertainty | Uncertainty type | Method | Disaggregated by | Article | QA score (%) |
|---|---|---|---|---|---|---|---|---|---|---|
| **DRC, 1995** | | | | | | | | | | |
| DRC | 1 Mar 1995 - 12 Jul 1995 | 0.73 | | Unspecified | | | Compartmental model | | Zaman 2009 | 100.0 |
| **West Africa 2013-2016** | | | | | | | | | | |
| Guinea | 22 Mar 2014 - 22 Aug 2014 | 1.04 | | Mean | 0.01 | Standard Deviation | Compartmental model | | Kiskowski 2014 | 100.0 |
| Guinea | Mar 2014 - Aug 2014 | 0.66 | | Unspecified | | | Branching process | Other, Region | Kucharski 2016 | 71.4 |
| Guinea | Mar 2014 - Aug 2014 | 7.00 | | Unspecified | | | Branching process | Other, Region | Kucharski 2016 | 71.4 |
| Guinea | Apr 2014 - 18 Mar 2015 | | 1.05 - 1.31 | Unspecified | | | Growth rate | Region, Time | Hsieh 2015 | 28.6 |
| Guinea | Aug 2014 - Feb 2015 | 2.00 | | Mean | | | Empirical (contact tracing) | | Valencia 2017 | 40.0 |
| Guinea | Aug 2014 - Feb 2015 | 3.00 | | Mean | | | Empirical (contact tracing) | | Valencia 2017 | 40.0 |
| Guinea, Liberia, Sierra Leone | 30 Dec 2013 - 18 Jul 2015 | | 0 - 3.5 | Unspecified | | | Compartmental model | Other, Region | Santermans 2016 | 100.0 |
| Guinea, Liberia, Sierra Leone | Dec 2013 - 25 Nov 2014 | | 0.93 - 1 | Other | | | Branching process | Region, Time | Agua-Agum 2015 | 100.0 |
| Guinea, Liberia, Sierra Leone | 2013 - 11 Nov 2014 | | 0.86 - 1.72 | Unspecified | | | Branching process | Region, Time | Evans 2015 | 42.9 |
| Guinea, Liberia, Sierra Leone | Jan 2014 - Feb 2015 | 2.30 | 1 - 7 | Mean | 1.8 - 2.7 | 95% CI | Unspecified | Time | Cleaton 2016 | 42.9 |
| Guinea, Liberia, Sierra Leone | 25 Mar 2014 - 22 Mar 2015 | | 0.62 - 1.38 | Mean | | | Growth rate | Region, Time | Wiratsudakul 2016 | 100.0 |
| Guinea, Liberia, Sierra Leone | 1 Jul 2014 - 8 Sep 2014 | | 1.2 - 2.3 | Unspecified | | | Compartmental model | Other, Region | Towers 2014 | 0.0 |
| Guinea, Liberia, Sierra Leone | 30 Dec 2014 - 31 Aug 2014 | | 1.38 - 1.81 | Mean | | | | Region | Aylward 2014 | 100.0 |
| Guinea, Liberia, Sierra Leone | 2014 - 2015 | 1.01 | | Mean | 0.27 - 2.58 | Range | Next generation matrix | | Denes 2019 | 57.1 |
| Guinea, Liberia, Sierra Leone | 18 Jun 2015 - 4 Aug 2015 | 2.58 | 0.64 - 4.74 | Unspecified | | | Empirical (contact tracing) | | Tiffany 2017 | 28.6 |
| Liberia | 22 Mar 2014 - 22 Aug 2014 | 1.17 | | Mean | 0.03 | Standard Deviation | Compartmental model | | Kiskowski 2014 | 100.0 |
| Liberia | Apr 2014 - 18 Mar 2015 | | 1.05 - 3.58 | Unspecified | | | Growth rate | Region, Time | Hsieh 2015 | 28.6 |
| Liberia | 27 May 2014 - 21 Dec 2014 | | 0.42 - 2.9 | Mean | | | Compartmental model | Time | Siettos 2015 | 14.3 |
| Liberia | 27 May 2014 - 22 Dec 2014 | | 0.38 - 6.1 | Mean | | | Compartmental model | Other, Time | Russo 2016 | 42.9 |
| Liberia | 7 Jul 2014 - 22 Sep 2014 | 1.73 | | Unspecified | 1.66 - 1.83 | 95% CI | Branching process | Symptoms, Other | Yamin 2016 | 100.0 |
| Liberia | Jul 2014 - Dec 2014 | 0.10 | | Median | 0.02 - 0.6 | 95% CI | Empirical (contact tracing) | | Lindblade 2015 | 100.0 |
| Liberia | Dec 2014 - Mar 2015 | 0.76 | | Unspecified | | | Branching process | Other, Region | Kucharski 2016 | 71.4 |
| Liberia | Dec 2014 - Mar 2015 | 5.00 | | Unspecified | | | Branching process | Other, Region | Kucharski 2016 | 71.4 |
| Liberia, Sierra Leone | 14 Apr 2014 - 11 Oct 2014 | | 1.9 - 2.5 | Unspecified | | | Other | Region | Majumder 2015 | 28.6 |
| Sierra Leone | 22 Mar 2014 - 22 Aug 2014 | 1.11 | | Mean | 0.02 | Standard Deviation | Compartmental model | | Kiskowski 2014 | 100.0 |
| Sierra Leone | 27 Apr 2014 - 3 Feb 2016 | | 0.16 - 3.31 | Mean | | | Branching process | Time | Bodine 2018 | 100.0 |
| Sierra Leone | Apr 2014 - 18 Mar 2015 | | 1.06 - 2.12 | Unspecified | | | Growth rate | Region, Time | Hsieh 2015 | 28.6 |
| Sierra Leone | 22 May 2014 - 3 Dec 2014 | | 1.1 - 2.13 | Unspecified | | | Compartmental model | Time | White 2015 | 57.1 |
| Sierra Leone | 25 May 2014 - 25 Jan 2015 | | 0.44 - 2.22 | Mean | | | Compartmental model | Region, Time | Yang 2015 | 85.7 |
| Sierra Leone | 27 May 2014 - 21 Dec 2014 | | 1.93 - 2.5 | Mean | | | Compartmental model | Time | Siettos 2015 | 14.3 |
| Sierra Leone | May 2014 - 22 Oct 2014 | 0.46 | | Unspecified | | | Unspecified | | Fang 2016 | 71.4 |

| Country | Survey date | Central estimate | Central range | Central type | Uncertainty | Uncertainty type | Method | Disaggregated by | Article | QA score (%) |
|---|---|---|---|---|---|---|---|---|---|---|
| Sierra Leone | Jul 2014 - Aug 2014 | 2.24 | | Mean | | | Branching process | | Ajelli 2015 | 71.4 |
| Sierra Leone | 11 Oct 2014 - Unspecified | 1.30 | | Unspecified | | | Empirical (contact tracing) | Other | DeSilva 2019 | 40.0 |
| Sierra Leone | 11 Oct 2014 - Unspecified | 4.00 | | Unspecified | | | Empirical (contact tracing) | Disease generation | DeSilva 2019 | 40.0 |
| Sierra Leone | 22 Oct 2014 - Sep 2015 | 0.09 | | Unspecified | | | Unspecified | | Fang 2016 | 71.4 |
| Sierra Leone | 21 Dec 2014 - 15 Aug 2015 | | 0.68 - 1.98 | Mean | | | Compartmental model | Time | Siettos 2016 | 42.9 |
| Sierra Leone | 2014 - Unspecified | 1.26 | | Unspecified | 1.04 - 1.54 | HPDI 95% | Genomics | | Alizon 2014 | 0.0 |
| Sierra Leone | 18 Jan 2015 | | 0.1 - 0.97 | Median | | | Compartmental model | Region | Camacho 2015 | 100.0 |
| Sierra Leone | Jul 2015 - Sep 2015 | 1.20 | 1.2 - 1.8 | Unspecified | | | Empirical (contact tracing) | | Glynn 2018 | 100.0 |
| Sierra Leone | Oct 2015 - Jan 2016 | | 0.83 - 9 | Unspecified | | | Empirical (contact tracing) | Time | Kelly 2018 | 85.7 |
| Sierra Leone | Sep 2016 - Jul 2017 | | 0 - 29 | Unspecified | | | Empirical (contact tracing) | Disease generation, Region | Kelly 2022 | 40.0 |
| **DRC, 2014** | | | | | | | | | | |
| DRC | 26 Jul 2014 - 7 Oct 2014 | 0.84 | | Unspecified | -0.38 - 2.06 | 95% CI | Empirical (contact tracing) | | Maganga 2014 | 28.6 |
| DRC | 26 Jul 2014 - 7 Oct 2014 | 1.29 | | Unspecified | -4.71 - 7.29 | 95% CI | Empirical (contact tracing) | | Maganga 2014 | 28.6 |
| **DRC, 2018** | | | | | | | | | | |
| DRC | 30 Apr 2018 - 24 May 2018 | 1.03 | 0.83 - 1.37 | Median | 0.83 - 1.37 | 95% CrI | Branching process | | Ahuka-Mundeke 2018 | 100.0 |
| **DRC, 2018-2020** | | | | | | | | | | |
| DRC | 30 Apr 2018 - 15 Jan 2019 | 0.90 | 0 - 1.8 | Median | 0.4 - 1.1 | 95% CI | Branching process | | Tariq 2019 | 100.0 |
| DRC | 8 May 2018 - 15 Apr 2019 | 1.11 | | Mean | | | Branching process | Other, Region, Time | Wannier 2019 | 85.7 |
| DRC | 5 Aug 2018 - 27 Jul 2019 | 2.49 | | Unspecified | | | Next generation matrix | | Juga 2020 | 28.6 |
| DRC | 2018 - 2020 | 0.33 | | Unspecified | | | Compartmental model | | Ouemba Tasse 2022 | 0.0 |
| DRC | Jan 2019 - Sep 2019 | | 0.4 - 1.1 | Median | | | Next generation matrix | Region, Time | Mizumoto 2019 | 100.0 |
| DRC | Apr 2019 - Aug 2019 | 1.06 | | Mean | 1.02 - 1.11 | 95% CI | Branching process | Other, Region, Time | Kelly 2020 | 85.7 |
| DRC | 12 Jun 2019 - 18 May 2020 | 1.14 | | Unspecified | | | Empirical (contact tracing) | | Keita 2023 | 28.6 |
| DRC | Jul 2019 | 2.19 | | Mean | | | Next generation matrix | Region, Time | Mizumoto 2019 | 100.0 |
| DRC | 29 Sep 2019 | 1.03 | | Median | 0.59 - 2.12 | 95% CrI | Next generation matrix | Region | Mizumoto 2019 | 100.0 |
| DRC | Sep 2019 | 1.20 | | Mean | | | Next generation matrix | Region, Time | Mizumoto 2019 | 100.0 |

24% of effective reproduction number estimates in Table S14 (unfiltered by QA score, n=55) were estimated using branching process models (n=13), 26% compartmental models (n=14), 11% next generation matrices (n=6), 22% contact tracing (n=12), 7% growth rate (n=4), 2% genomics (n=1) and 7% with an unspecified or "other" method (n=4).

##### 4.3.4 Secondary attack rate

Table S15: Secondary attack rates. Note: The three Fang 2016 estimates had no units, so an assumption was made to convert them into percentages.

| Country | Survey date | Central estimate (%) | Central range | Uncertainty (95% CI) | Population Sample | Sample size | Population Group | Disaggregated by | Article | QA score (%) |
|---|---|---|---|---|---|---|---|---|---|---|
| **DRC, 1976** | | | | | | | | | | |
| DRC | 1976 | | 0.1 - 0.8 | | Unspecified | | Unspecified | Unspecified | Emond 1977 | 0.0 |
| **DRC, 1977** | | | | | | | | | | |
| DRC | 1 Sep 1977 - 24 Oct 1977 | 5.6 | 3.4 - 27.3 | | Community based | | Other | Disease generation, Level of exposure, Other | International Commission 1978 | 40.0 |
| **South Sudan, 1976** | | | | | | | | | | |
| Sudan | Jun 1976 - Nov 1976 | 12 | 0.34 - 14 | | Household based | | Persons under investigation | Disease generation | WHO/Int. Study Team 1978 | 60.0 |
| Sudan | Jun 1976 - Nov 1976 | | 23 - 81 | | Household based | 17 | Persons under investigation | Level of exposure | WHO/Int. Study Team 1978 | 60.0 |
| **DRC 1981-1985*** | | | | | | | | | | |
| DRC | 1981 - 1985 | 15 | 7 - 19 | | Contact based | 188 | General population | Age | Jezek 1999 | 20.0 |
| **DRC, 1995** | | | | | | | | | | |
| DRC | 1 Jan 1995 - 3 Jun 1995 | 16 | | | Household based | 200 | Persons under investigation | | Dowell 1999 | 85.7 |
| **Uganda, 2000-2001** | | | | | | | | | | |
| Uganda | Aug 2000 - Jan 2001 | 2.5 | | | Contact based | ~1000 | Persons under investigation | Age, Sex | Okware 2002 | 40.0 |
| Uganda | 21 Oct 2000 - 22 Dec 2000 | 26 | | | Community based | 73 | Persons under investigation | | Borchert 2011 | 20.0 |
| **West Africa 2013-2016** | | | | | | | | | | |
| Guinea | 20 Sep 2014 - 31 Dec 2014 | | 2.1 - 7.9 | | Community based | 152 | Persons under investigation | Region | Dixon 2015 | 80.0 |
| Guinea, Liberia, Sierra Leone | 18 Jun 2015 - 4 Aug 2015 | 65 | 33 - 89 | | Contact based | 301 | Persons under investigation | Region | Tiffany 2017 | 28.6 |
| Sierra Leone | May 2014 - Sep 2015 | 5.9 | 5.6 - 6.2 | 5 - 7 | Household based | 1154 | Persons under investigation | Other | Fang 2016 | 71.4 |
| Sierra Leone | May 2014 - 22 Oct 2014 | 9.3 | | 7.7 - 11 | Household based | | Persons under investigation | | Fang 2016 | 71.4 |
| Sierra Leone | 22 Oct 2014 - Sep 2015 | 1.7 | | 1 - 3.1 | Household based | | Persons under investigation | | Fang 2016 | 71.4 |
| Sierra Leone | 15 Dec 2014 - 30 Apr 2015 | 9.9 | 3.3 - 26.8 | | Household based | 838 | Persons under investigation | Age, Level of exposure, Other, Sex, Symptoms, Time | Reichler 2018 | 80.0 |
| Sierra Leone | Jul 2015 - Sep 2015 | | 18 - 38 | | Household based | 937 | Household contacts of survivors | Other | Glynn 2018 | 100.0 |

##### 4.3.5 Growth rate

Table S16: Growth rates

| Country | Survey date | Central estimate | Unit | Central range | Central type | Uncertainty | Uncertainty type | Disaggregated by | Article | QA score (%) |
|---|---|---|---|---|---|---|---|---|---|---|
| **DRC, 1976** | | | | | | | | | | |
| DRC | 1976 - Unspecified | 1.43 | Per day | | Other/Unspecified | 0.69 - 2.62 | 95% CI | | Viboud 2016 | 75.0 |
| **West Africa 2013-2016** | | | | | | | | | | |
| Guinea | 22 Mar 2014 - 31 Aug 2014 | 0.01 | Per day | | Other/Unspecified | | | | Weitz 2015 | 75.0 |
| Guinea | 25 Mar 2014 - 3 May 2015 | 0.02 | Per day | | Mean | 0 | Standard Deviation | | Liu 2015 | 71.4 |
| Guinea, Liberia, Sierra Leone | 25 Mar 2014 - 3 May 2015 | 0.03 | Per day | | Mean | 0 | Standard Deviation | | Liu 2015 | 71.4 |
| Liberia | 17 Mar 2014 - Aug 2014 | 0.05 | Per day | | Other/Unspecified | 0 | Standard Error | | Valdez 2015 | 57.1 |
| Liberia | 25 Mar 2014 - 3 May 2015 | 0.09 | Per day | | Mean | 0.01 | Standard Deviation | | Liu 2015 | 71.4 |
| Liberia | 22 Jun 2014 - 31 Aug 2014 | 0.05 | Per day | | Other/Unspecified | | | | Weitz 2015 | 75.0 |
| Sierra Leone | 25 Mar 2014 - 3 May 2015 | 0.05 | Per day | | Mean | 0 | Standard Deviation | | Liu 2015 | 71.4 |
| Sierra Leone | 28 May 2014 - 31 Aug 2014 | 0.03 | Per day | | Other/Unspecified | | | | Weitz 2015 | 75.0 |
| **DRC, 2018-2020** | | | | | | | | | | |
| DRC | 6 Sep 2018 - 11 Mar 2019 | 0.12 | Per week | | Median | 0.11 - 0.14 | 95% CI | | Chowell 2019 | 85.7 |
| **Uganda, 2022-2023** | | | | | | | | | | |
| Uganda | Aug 2022 - Nov 2022 | 0.08 | Per day | | Other/Unspecified | 0.08 - 0.08 | 95% CrI | | Marziano 2023 | 100.0 |
| Uganda | Aug 2022 - Nov 2022 | 0.11 | Per day | | Other/Unspecified | 0.05 - 0.19 | 95% CrI | | Marziano 2023 | 100.0 |
| **Multiple outbreaks** | | | | | | | | | | |
| Uganda, Guinea, Liberia, Sierra Leone | 2000 - 2014 | | Per week | 0.08 - 2.38 | Other/Unspecified | | | Region, Time | Viboud 2016 | 75.0 |

##### 4.3.6 Doubling time

Table S17: Doubling times

| Country | Survey date | Central estimate (Days) | Central range | Uncertainty (95% CI) | Disaggregated by | Article | QA score (%) |
|---|---|---|---|---|---|---|---|
| **West Africa 2013-2016** | | | | | | | |
| Guinea | 22 Mar 2014 - 31 Aug 2014 | 61.0 | | | | Weitz 2015 | 75.0 |
| Guinea, Liberia, Nigeria, Sierra Leone | 30 Dec 2014 - 31 Aug 2014 | | 12.84 - 59.75 | | Region, Time | Aylward 2014 | 100.0 |
| Guinea, Liberia, Sierra Leone | Dec 2013 - 25 Nov 2014 | | -3322.71 - 8455.5 | | Region, Time | Agua-Agum 2015 | 100.0 |
| Guinea, Sierra Leone | 28 Sep 2014 - 11 Nov 2014 | 22.1 | | 18.9 - 25.59 | | Tong 2015 | 42.9 |
| Liberia | 22 Jun 2014 - 31 Aug 2014 | 14.0 | | | | Weitz 2015 | 75.0 |
| Sierra Leone | 28 May 2014 - 31 Aug 2014 | 21.0 | | | | Weitz 2015 | 75.0 |

##### 4.3.7 Overdispersion

Table S18: Overdispersion table

| Country | Survey date | Central estimate | Central range | Central type | Uncertainty | Uncertainty type | Method | Disaggregated by | Article | QA score (%) |
|---|---|---|---|---|---|---|---|---|---|---|
| **West Africa 2013-2016** | | | | | | | | | | |
| Guinea | Dec 2013 - Mar 2016 | 0.310 | | Mean | 0.25 - 0.37 | 95% CI | Empirical (contact tracing) | | Robert 2019 (b) | 100.0 |
| Guinea | Mar 2014 - Aug 2014 | 0.190 | | Other/Unspecified | | | Branching process | Other, Region | Kucharski 2016 | 71.4 |
| Guinea | Mar 2014 - Aug 2014 | 1.600 | | Other/Unspecified | | | Branching process | Other, Region | Kucharski 2016 | 71.4 |
| Guinea, Liberia, Sierra Leone | 30 Dec 2013 - 8 Jul 2015 | | 0.62 - 4.17 | Other/Unspecified | | | Branching process | Region | Santermans 2016 | 100.0 |
| Guinea, Liberia, Sierra Leone | Dec 2013 - May 2015 | 0.030 | | Other/Unspecified | | | Branching process | | Agua-Agum 2016 | 100.0 |
| Guinea, Liberia, Sierra Leone | Dec 2013 - May 2015 | 0.520 | | Other/Unspecified | | | Branching process | | Agua-Agum 2016 | 100.0 |
| Liberia | 4 Jul 2014 - 2 Sep 2014 | 2.200 | | Mean | 2 - 2.4 | IQR | Branching process | | Drake 2015 | 57.1 |
| Liberia | Dec 2014 - Mar 2015 | 0.820 | | Other/Unspecified | | | Branching process | Other, Region | Kucharski 2016 | 71.4 |
| Multi-country: Africa, Europe, USA (n = 12) | Jul 2014 - Mar 2015 | 0.090 | 0.09 - 0.5 | Other/Unspecified | 0.03 - 0.2 | 90% CI | Branching process | Other | Toth 2015 | 71.4 |
| Sierra Leone | Jul 2014 - Aug 2014 | 0.450 | | Other/Unspecified | 0.19 - 1.32 | 95% CI | Empirical (contact tracing) | | Ajelli 2015 | 71.4 |
| Sierra Leone | 4 Aug 2014 - 29 Mar 2015 | 0.065 | | Mean | 0.037 - 0.11 | HPDI 95% | Branching process | Time | Zhang 2022 | 57.1 |
| Sierra Leone | 20 Oct 2014 - 30 Mar 2015 | 0.370 | | Mean | | | Other | | Lau 2017 (b) | 100.0 |
| Sierra Leone | 20 Oct 2014 - 30 Mar 2015 | 0.470 | | Other/Unspecified | | | Other | | Lau 2017 | 85.7 |
| **DRC, 2018-2020** | | | | | | | | | | |
| DRC | 31 Jul 2018 - 26 Apr 2020 | 0.270 | | Other/Unspecified | 0.2 - 0.33 | 95% CrI | Empirical (contact tracing) | | Polonsky 2021 | 100.0 |
| **Multiple outbreaks** | | | | | | | | | | |
| Multi-country: Africa (n = 6) | 2000 - 2015 | 0.240 | 0.24 - 2.09 | Median | 0.06 | Standard Error | Empirical (contact tracing) | Other | Taube 2022 | 71.4 |

##### 4.3.8 Risk factors associated with transmission

Table S19: Risk factors for infection table. Total is the number of parameters extracted for each risk factor for infection.

| Risk Factor for Infection | Significant | | | Not significant | | | Unspecified significance | | | Total |
|---|---|---|---|---|---|---|---|---|---|---|
| | Adjusted | Not adjusted | Unspecified | Adjusted | Not adjusted | Unspecified | Adjusted | Not adjusted | Unspecified | |
| Close contact | 6 | 4 | 5 | 2 | 1 | 2 | 0 | 0 | 2 | 22 |
| Age | 5 | 2 | 2 | 0 | 4 | 3 | 1 | 0 | 0 | 17 |
| Sex | 2 | 1 | 0 | 5 | 4 | 3 | 1 | 0 | 0 | 16 |
| Funeral | 2 | 3 | 2 | 1 | 2 | 1 | 0 | 0 | 3 | 14 |
| Household contact | 3 | 2 | 0 | 0 | 0 | 1 | 0 | 0 | 1 | 7 |
| Occupation | 2 | 2 | 0 | 1 | 1 | 1 | 0 | 0 | 0 | 7 |
| Non-household contact | 1 | 1 | 0 | 0 | 0 | 0 | 0 | 0 | 1 | 3 |
| Hospitalisation | 0 | 2 | 0 | 0 | 0 | 0 | 0 | 0 | 1 | 3 |
| Contact with animal | 0 | 0 | 0 | 0 | 1 | 1 | 0 | 0 | 0 | 2 |
| Social gathering | 0 | 0 | 0 | 0 | 1 | 0 | 0 | 0 | 0 | 1 |
| Other | 8 | 5 | 5 | 6 | 3 | 6 | 1 | 1 | 2 | 37 |

Table S20: Risk factors for onward transmission by significance and adjustment.

| Risk Factor for Onward Transmission | Significant | Not significant |
|---|---|---|
| | Adjusted | |
| Age | 1 | 0 |
| First generation of transmission chain | 1 | 0 |
| Funeral | 1 | 0 |
| Hospitalisation | 1 | 0 |
| Sex | 1 | 0 |
| Socioeconomic status | 1 | 0 |
| Survival | 1 | 0 |
| Location (rural/urban) | 0 | 1 |

#### 4.4 Temporal dynamics of disease progression

##### 4.4.1 Time periods for the infection process

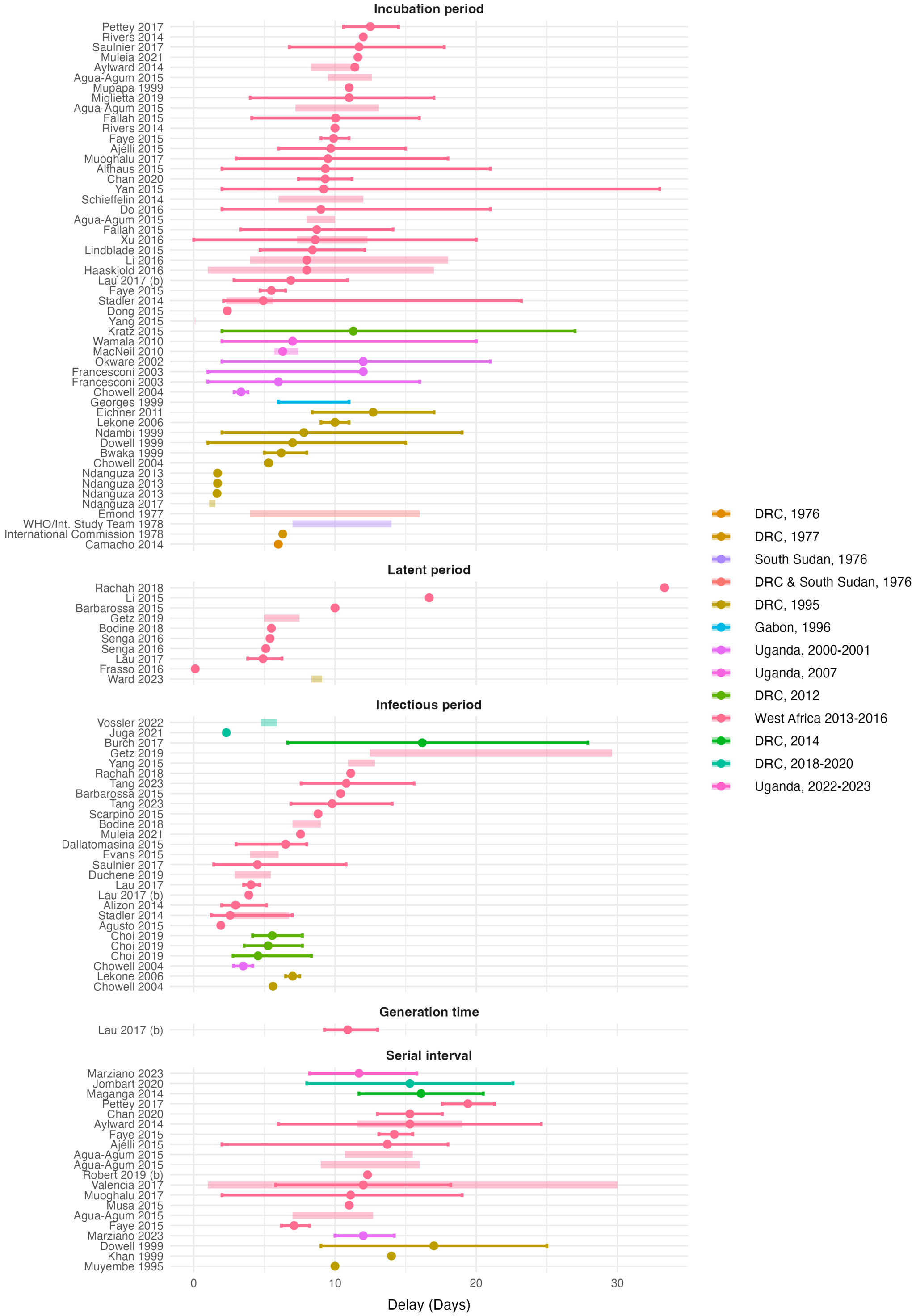

Figure S2: Time periods for the infection process, including the incubation period, latent period, infectious period, generation time and serial interval. This figure excludes one outlier (Martinez et al, 2022).

##### 4.4.2 Delays beyond symptom onset

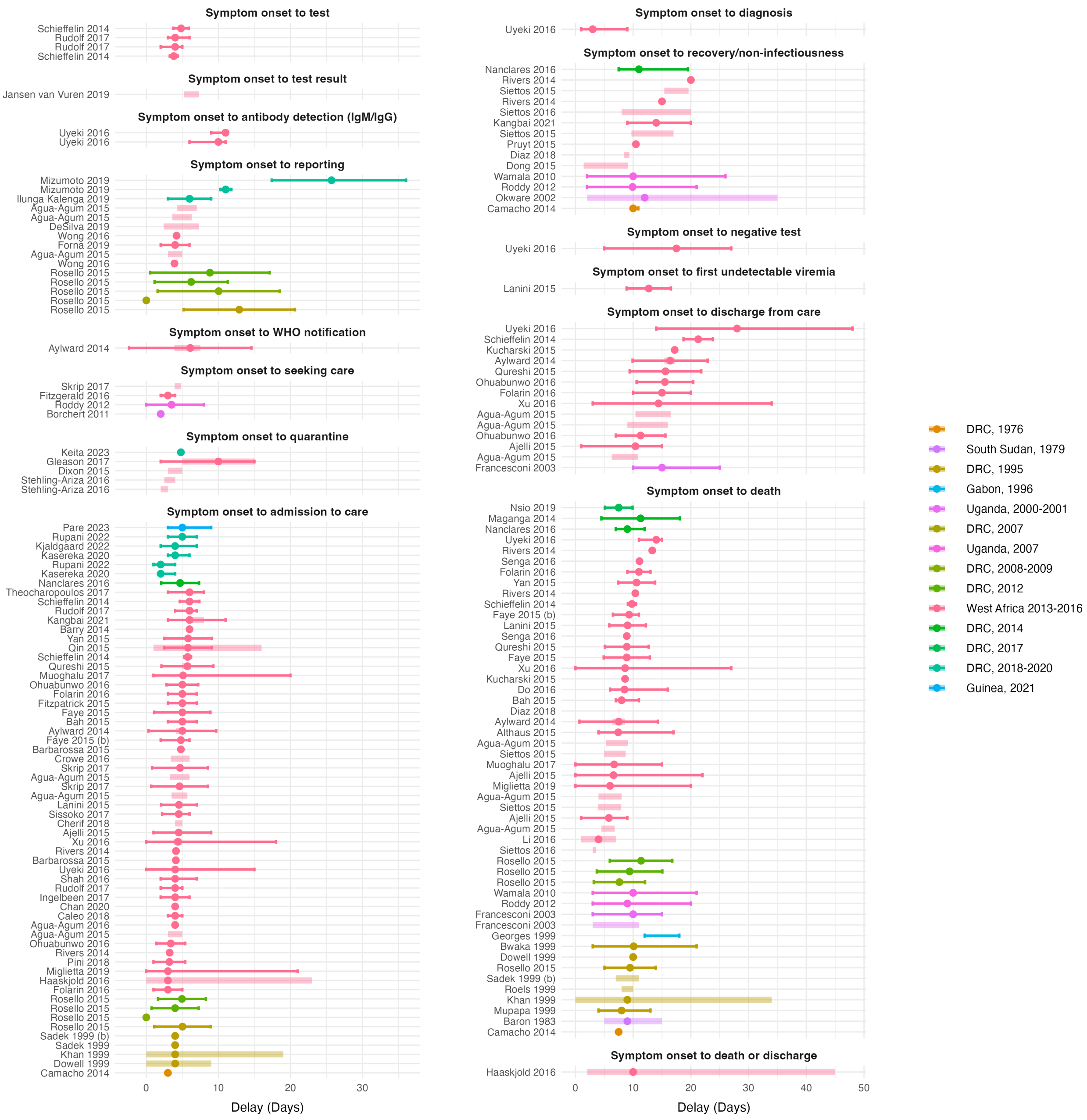

Figure S3: All delays from symptom onset to various endpoints, including test, test result, antibody detection, reporting, WHO notification, seeking care, quarantine, admission to care, diagnosis, recovery/non-infectiousness, negative test, undetectable viremia, discharge from care, death, and discharge or death (when no distinction was made between these outcomes).

##### 4.4.3 Delays beyond admission to care

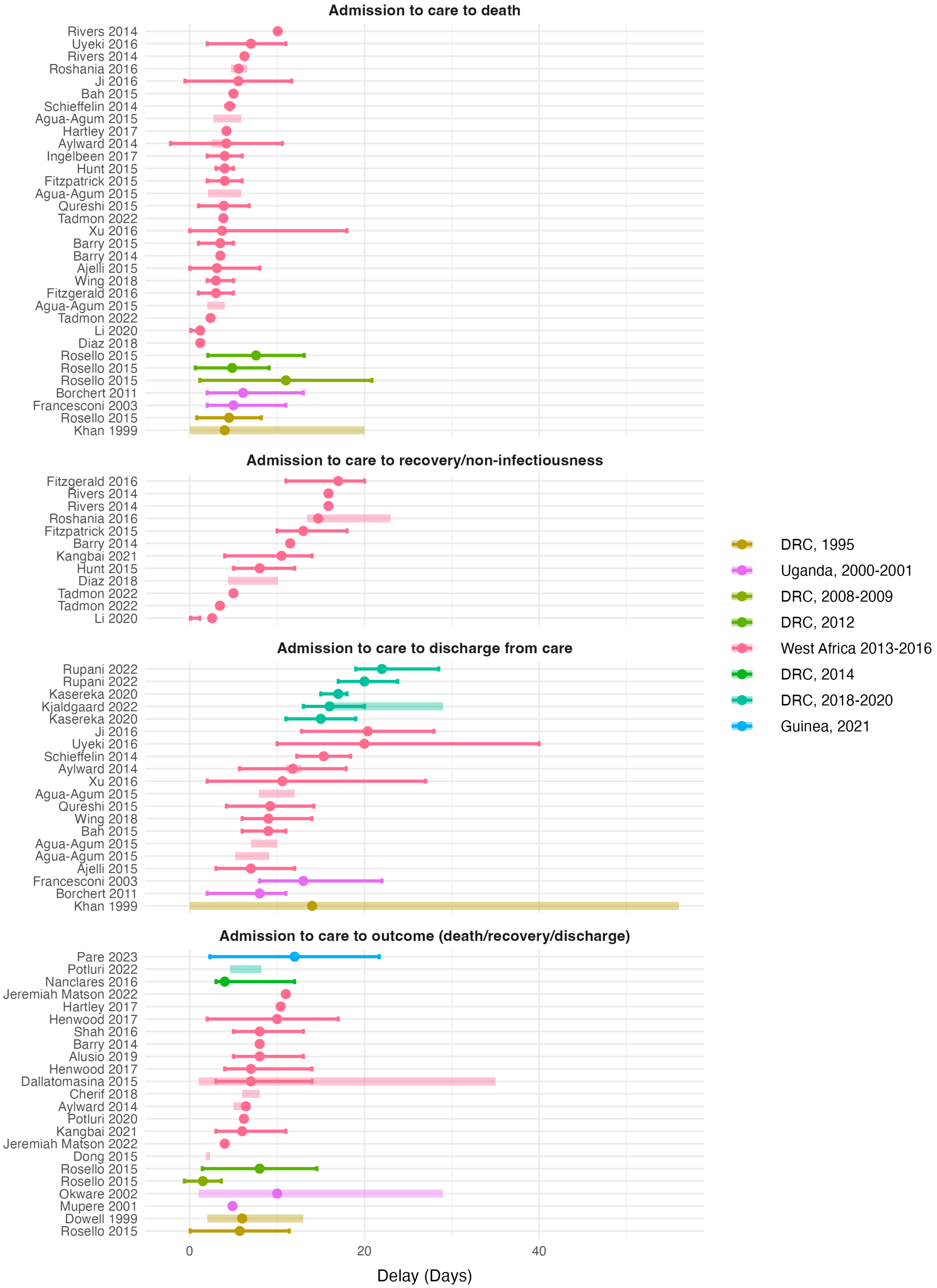

Figure S4: Delays beyond admission to care

###### 4.4.4 QA unfiltered meta-analysis by Ebola virus species

A

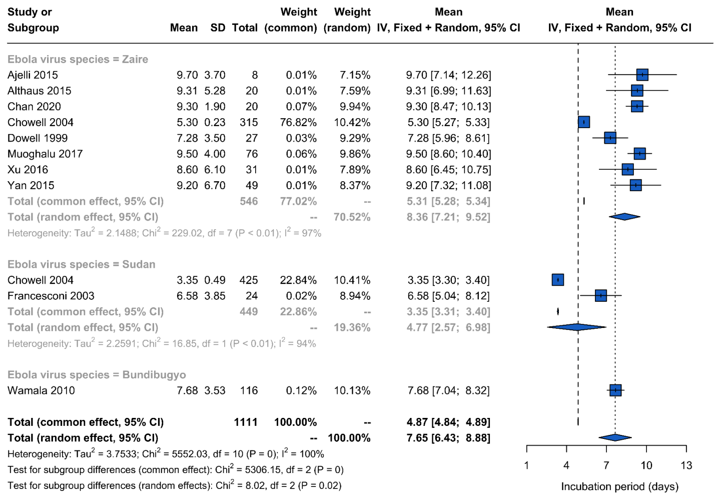

B

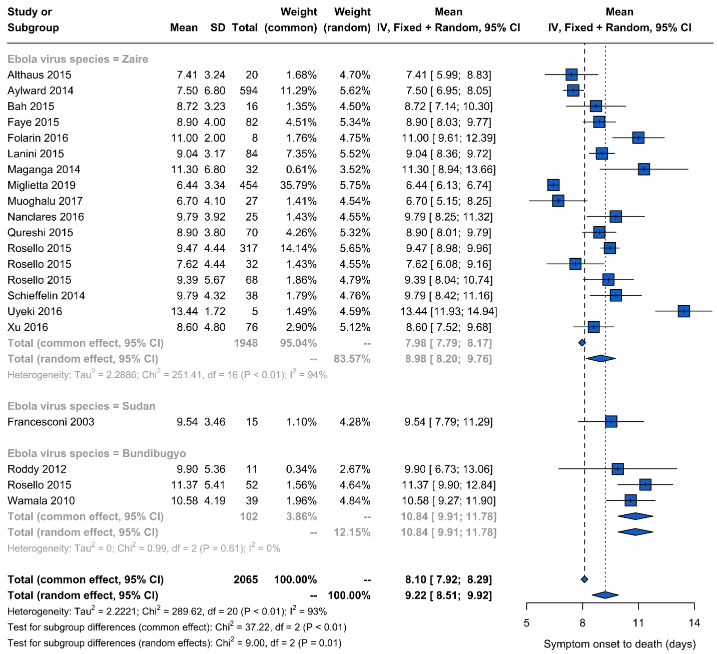

C

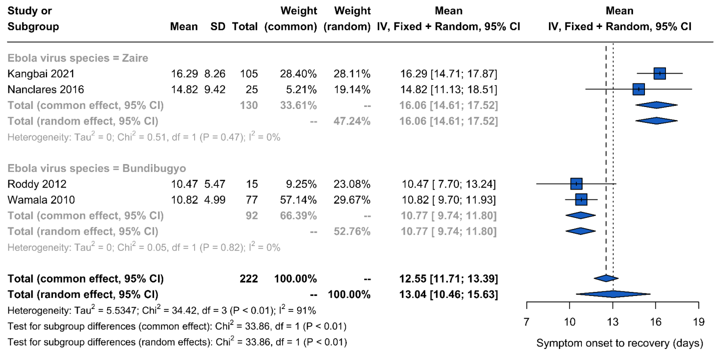

Figure S5: Meta-analysis for A) incubation period B) onset-to-death and C) onset-to-recovery regardless of QA score, with a species sub-group analysis

##### 4.4.5 QA unfiltered meta-analysis

**A**

| Study | Mean | SD | Total | Weight (common) | Weight (random) | Mean IV, Fixed + Random, 95% CI |
|---|---|---|---|---|---|---|
| Chowell 2004 | 5.61 | 0.19 | 315 | 89.88% | 33.60% | 5.61 [5.59; 5.63] |
| Chowell 2004 | 3.50 | 0.67 | 425 | 9.75% | 33.57% | 3.50 [3.44; 3.56] |
| Dallatomasina 2015 | 6.02 | 3.72 | 489 | 0.36% | 32.83% | 6.02 [5.69; 6.35] |
| **Total (common effect, 95% CI)** | | | **1229** | **100.00%** | **--** | **5.41 [5.39; 5.43]** |
| **Total (random effect, 95% CI)** | | | **--** | | **100.00%** | **5.04 [3.79; 6.29]** |

Heterogeneity: $Tau^2 = 1.2111$; $Chi^2 = 3815.78$, df = 2 (P = 0); $I^2 = 100\%$

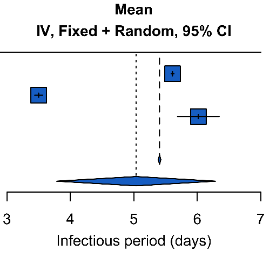

**B**

| Study | Mean | SD | Total | Weight (common) | Weight (random) | Mean IV, Fixed + Random, 95% CI |
|---|---|---|---|---|---|---|
| Ajelli 2015 | 13.70 | 4.50 | 12 | 0.95% | 9.26% | 13.70 [11.15; 16.25] |
| Aylward 2014 | 15.30 | 9.30 | 92 | 1.70% | 10.42% | 15.30 [13.40; 17.20] |
| Chan 2020 | 15.30 | 2.30 | 20 | 6.03% | 11.76% | 15.30 [14.29; 16.31] |
| Dowell 1999 | 17.14 | 2.93 | 200 | 37.30% | 12.27% | 17.14 [16.74; 17.55] |
| Jombart 2020 | 15.30 | 7.30 | 1426 | 42.70% | 12.29% | 15.30 [14.92; 15.68] |
| Maganga 2014 | 16.10 | 4.40 | 28 | 2.31% | 10.88% | 16.10 [14.47; 17.73] |
| Muoghalu 2017 | 11.10 | 5.20 | 47 | 2.77% | 11.10% | 11.10 [ 9.61; 12.59] |
| Pettey 2017 | 19.40 | 5.10 | 28 | 1.72% | 10.44% | 19.40 [17.51; 21.29] |
| Valencia 2017 | 12.00 | 6.20 | 109 | 4.53% | 11.57% | 12.00 [10.84; 13.16] |
| **Total (common effect, 95% CI)** | | | **1962** | **100.00%** | **--** | **15.79 [15.55; 16.04]** |
| **Total (random effect, 95% CI)** | | | **--** | | **100.00%** | **15.05 [13.50; 16.59]** |

Heterogeneity: $Tau^2 = 5.0110$; $Chi^2 = 146.03$, df = 8 (P < 0.01); $I^2 = 95\%$

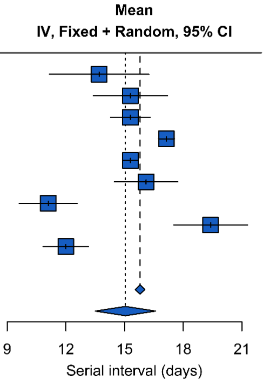

Figure S6: Meta-analysis for A) infectious period and B) serial interval regardless of QA score

#### 4.5 Severity

##### 4.5.1 Case Fatality Ratio (CFR)

Table S21: Case fatality ratio results

| Country | Survey date | Central estimate | Deaths | Cases | Central range | Uncertainty | Uncertainty type | Adjustment | Population Sample | Disaggregated by | Article | QA score (%) |
|---|---|---|---|---|---|---|---|---|---|---|---|---|
| **DRC, 1976** | | | | | | | | | | | | |
| DRC | Aug 1976 - Nov 1976 | 88 | | | | 80 - 94 | 95% CI | Unspecified | Population based | | Camacho 2014 | 71.4 |
| DRC | Aug 1976 - Oct 1976 | 96 | 252 | 262 | | 93 - 98 | 95% CI | Adjusted | Unspecified | Age, Time | Rosello 2015 | 100.0 |
| DRC | 1 Sep 1976 - 24 Oct 1976 | 88 | 280 | 318 | | | | Unspecified | Hospital based | | International Commission 1978 | 40.0 |
| **South Sudan, 1976** | | | | | | | | | | | | |
| Sudan | Jun 1976 - Nov 1976 | 53 | 151 | 284 | 35 - 67 | | | Naive | Hospital based | Age, Region, Sex, Time | WHO/Int. Study Team 1978 | 60.0 |
| **South Sudan, 1979** | | | | | | | | | | | | |
| South Sudan | 31 Jul 1979 - 6 Oct 1979 | 65 | 22 | 34 | 0 - 100 | | | Unspecified | Hospital based | Time | Baron 1983 | 14.3 |
| **DRC 1981-1985*** | | | | | | | | | | | | |
| DRC | 1981 - 1985 | 43 | 9 | 21 | | | | Naive | Hospital based | | Jezek 1999 | 20.0 |
| **Gabon, 1994** | | | | | | | | | | | | |
| Gabon | Dec 1994 - Feb 1995 | 59 | 29 | 49 | | | | Unspecified | Hospital based | | Georges 1999 | 80.0 |
| **DRC, 1995** | | | | | | | | | | | | |
| DRC | 1 Jan 1995 - 3 Jun 1995 | 95 | 52 | 55 | | | | Unspecified | Household based | | Dowell 1999 | 85.7 |
| DRC | 6 Jan 1995 - 16 Jul 1995 | 81 | 250 | 310 | | | | Unspecified | Community based | | Khan 1999 | 42.9 |
| DRC | 13 Jan 1995 - 24 Aug 1995 | 77 | | | | | | Unspecified | Population based | | Butler 1996 | 40.0 |
| DRC | Jan 1995 - Jun 1995 | 78 | 248 | 317 | | 73 - 83 | 95% CI | Adjusted | Unspecified | Age, Time | Rosello 2015 | 100.0 |
| DRC | 1 Mar 1995 - 16 Jul 1995 | | | | 82 - 83 | | | Unspecified | Population based | Other | Ndanguza 2017 | 57.1 |
| DRC | 9 Apr 1995 - 23 May 1995 | 74 | 101 | 136 | | | | Naive | Population based | | Muyembe 1995 | 50.0 |
| DRC | Apr 1995 - Jun 1995 | 96 | 14 | 15 | | | | Adjusted | Hospital based | | Mupapa 1999 | 20.0 |
| DRC | Apr 1995 - Unspecified | 78 | 18 | 23 | | | | Unspecified | Hospital based | | Ndambi 1999 | 100.0 |
| DRC | May 1995 | 81 | 250 | 310 | 69 - 96 | | | Naive | Hospital based | Age, Occupation, Other, Sex, Time | Sadek 1999 (b) | 60.0 |
| DRC | 1995 | | | | 54 - 83 | | | Naive | Unspecified | Other | Roels 1999 | 40.0 |
| **Gabon, 1996** | | | | | | | | | | | | |
| Gabon | Feb 1996 - Unspecified | | 21 | 37 | | | | Naive | Population based | | Amblard 1997 | 20.0 |
| Gabon | Feb 1996 - Unspecified | 68 | 21 | 31 | | | | Unspecified | Population based | | Georges 1999 | 80.0 |
| Gabon | Jul 1996 - Dec 1996 | | 40 | 52 | | | | Naive | Population based | | Amblard 1997 | 20.0 |
| Gabon | Oct 1996 - Mar 1997 | 75 | 45 | 60 | | | | Unspecified | Population based | | Georges 1999 | 80.0 |
| **Uganda, 2000-2001** | | | | | | | | | | | | |
| Uganda | Aug 2000 - Jan 2001 | 53 | 224 | 425 | | | | Unspecified | | | Okware 2002 | 40.0 |
| Uganda | 8 Oct 2000 - 27 Feb 2001 | 53 | 224 | 425 | 52 - 80 | | | Naive | Population based | Region | Lamunu 2004 | 40.0 |
| Uganda | 21 Oct 2000 - 22 Dec 2000 | 69 | 18 | 26 | | | | Naive | Community based | Other | Borchert 2011 | 20.0 |
| Uganda | Oct 2000 - Feb 2001 | 40 | 8 | 20 | | | | Adjusted | Hospital based | Other | Mupere 2001 | 40.0 |
| Uganda | Oct 2000 - Feb 2001 | 28 | 18 | 64 | | | | Adjusted | Hospital based | Other | Mupere 2001 | 40.0 |
| Uganda | Oct 2000 - Feb 2001 | 8 | 7 | 84 | | | | Adjusted | Hospital based | Other | Mupere 2001 | 40.0 |
| Uganda | 2000 - 2001 | | | | 29 - 77 | | | Unspecified | Unspecified | Age | McElroy 2014 | 100.0 |
| Uganda | 2000 | | | | 0 - 100 | | | Unspecified | | | Okware 2015 | 20.0 |
| **Republic of the Congo, 2005** | | | | | | | | | | | | |
| Republic of the Congo | 18 Apr 2005 - 8 Jul 2005 | 83 | 10 | 12 | | | | Unspecified | Community based | Region | Nkoghe 2011 (b) | 42.9 |
| **DRC, 2007** | | | | | | | | | | | | |
| DRC | Apr 2007 - Oct 2007 | 74 | 187 | | | 68 - 79 | 95% CI | Adjusted | Unspecified | Age, Time | Rosello 2015 | 100.0 |
| **Uganda, 2007** | | | | | | | | | | | | |
| Uganda | Aug 2007 - Dec 2007 | 32 | | | | | | Unspecified | | | MacNeil 2011 | 42.9 |
| Uganda | 29 Nov 2007 - 20 Feb 2008 | 42 | 11 | 26 | | | | Naive | Hospital based | | Roddy 2012 | 57.1 |
| Uganda | 29 Nov 2007 - 20 Feb 2008 | 25 | 37 | 149 | | | | Naive | Population based | | Roddy 2012 | 57.1 |
| Uganda | 29 Nov 2007 - 20 Feb 2008 | 34 | 39 | 116 | 29 - 100 | | | Unspecified | Community based | Region, Sex, Other | Wamala 2010 | 80.0 |
| Uganda | 2007 - Unspecified | 40 | 17 | 43 | | | | Adjusted | Community based | | MacNeil 2010 | 85.7 |
| **DRC, 2008-2009** | | | | | | | | | | | | |
| DRC | Nov 2008 - Jan 2009 | 44 | 14 | 32 | | 26 - 62 | 95% CI | Adjusted | Unspecified | Age, Time | Rosello 2015 | 100.0 |

| Country | Survey date | Central estimate | Deaths | Cases | Central range | Uncertainty | Uncertainty type | Adjustment | Population Sample | Disaggregated by | Article | QA score (%) |
|---|---|---|---|---|---|---|---|---|---|---|---|---|
| **DRC, 2012** | | | | | | | | | | | | |
| DRC | Jun 2012 - Nov 2012 | 54 | 28 | 52 | | 39 - 68 | 95% CI | Adjusted | Unspecified | Age, Time | Rosello 2015 | 100.0 |
| DRC | 2 Aug 2012 - 26 Nov 2012 | 54 | 28 | 52 | 0 - 100 | | | Adjusted | Other | Age, Other, Sex | Kratz 2015 | 71.4 |
| DRC | Aug 2012 - Oct 2012 | 74 | 49 | 66 | | 62 - 84 | 95% CI | Adjusted | Unspecified | Age, Time | Rosello 2015 | 100.0 |
| **West Africa 2013-2016** | | | | | | | | | | | | |
| Guinea | Dec 2013 - 25 Nov 2014 | 66 | | | | | | Unspecified | Community based | | Helleringer 2015 | 100.0 |
| Guinea | 1 Jan 2014 - 29 Mar 2015 | 46 | 429 | 932 | 40 - 60 | 43 - 49 | 95% CI | Adjusted | Hospital based | Region | Rico 2016 | 80.0 |
| Guinea | 1 Jan 2014 - 29 Mar 2015 | 60 | 817 | 1,355 | 53 - 76 | | | Naive | Community based | Region | Rico 2016 | 80.0 |
| Guinea | 1 Jan 2014 - 29 Mar 2015 | 40 | 168 | 420 | 23 - 44 | 35 - 45 | 95% CI | Adjusted | Hospital based | Region | Rico 2016 | 80.0 |
| Guinea | 1 Jan 2014 - 29 Mar 2015 | 53 | 295 | 553 | 44 - 60 | | | Naive | Community based | Region | Rico 2016 | 80.0 |
| Guinea | Jan 2014 - Dec 2015 | 63 | 437 | 695 | | | | Adjusted | Hospital based | | Cherif 2017 | 100.0 |
| Guinea | Jan 2014 - Dec 2015 | 68 | 1,574 | 2,313 | 66 - 81 | | | Adjusted | Hospital based | Age | Cherif 2018 | 100.0 |
| Guinea | 10 Feb 2014 - 11 Aug 2014 | 54 | | | | 49 - 63 | 95% CI | Unspecified | Community based | | Faye 2015 | 85.7 |
| Guinea | 1 Mar 2014 - 28 Feb 2015 | | | | 35 - 49 | | | Unspecified | Hospital based | Time | Faye 2015 (b) | 71.4 |
| Guinea | 17 Mar 2014 - 29 Mar 2015 | 60 | 719 | 1,205 | | | | Naive | Hospital based | Age, Other | Kerber 2016 | 40.0 |
| Guinea | 22 Mar 2014 - 25 Jan 2015 | 66 | | | | | | Unspecified | Unspecified | | Li 2015 | 42.9 |
| Guinea | 25 Mar 2014 - 26 Apr 2014 | 43 | 16 | 37 | | | | Adjusted | Hospital based | | Bah 2015 | 28.6 |
| Guinea | 25 Mar 2014 - 20 Aug 2014 | 44 | 39 | 90 | | 33 - 54 | 95% CI | Naive | Hospital based | | Barry 2014 | 14.3 |
| Guinea | Mar 2014 - Unspecified | 71 | | | | | | Unspecified | Hospital based | | Baize 2014 | 42.9 |
| Guinea | Mar 2014 - Unspecified | 86 | | | | | | Unspecified | Hospital based | | Baize 2014 | 42.9 |
| Guinea | Mar 2014 - Aug 2014 | 44 | 39 | 89 | | | | Naive | Hospital based | | Barry 2015 | 57.1 |
| Guinea | Mar 2014 - May 2015 | 48 | 70 | 146 | 23 - 74 | | | Unspecified | Hospital based | Other, Region, Sex, Symptoms | Sow 2020 | 80.0 |
| Guinea | 20 Sep 2014 - 31 Dec 2014 | 81 | 123 | 152 | 79 - 84 | | | Naive | Community based | Region | Dixon 2015 | 80.0 |
| Guinea | 29 Nov 2014 - 31 Jan 2015 | 59 | | | | | | Unspecified | Hospital based | | Vernet 2017 | 71.4 |
| Guinea | 1 Dec 2014 - 1 Mar 2015 | | | | 19 - 81 | | | Unspecified | Population based | Other | Dong 2015 | 14.3 |
| Guinea | 2 Dec 2014 - 23 Feb 2015 | 61 | 46 | 76 | | | | Naive | Hospital based | | Loubet 2016 | 0.0 |
| Guinea | 22 Jun 2017 - 9 Jul 2017 | 56 | 10 | 18 | | 31 - 78 | 95% CI | Unspecified | Household based | | Timothy 2019 | 100.0 |
| Guinea, Liberia, Nigeria, Sierra Leone | 30 Dec 2013 - 14 Sep 2014 | 71 | | | 32 - 84 | 69 - 73 | 95% CI | Adjusted | | Age, Occupation, Region, Sex, Symptoms, Time, Other | Aylward 2014 | 100.0 |
| Guinea, Liberia, Sierra Leone | 28 Dec 2013 - 3 Oct 2014 | 73 | | | | | | Unspecified | Community based | | Barbarossa 2015 | 42.9 |
| Guinea, Liberia, Sierra Leone | 28 Dec 2013 - 3 Oct 2014 | 61 | | | | | | Unspecified | Hospital based | | Barbarossa 2015 | 42.9 |
| Guinea, Liberia, Sierra Leone | 30 Dec 2013 - 28 Sep 2015 | 63 | | | 40 - 86 | 62 - 64 | 95% CI | Adjusted | | Age, Other, Region, Time | Garske 2016 | 100.0 |
| Guinea, Liberia, Sierra Leone | 30 Dec 2013 - 8 Jul 2015 | | | | 19 - 66 | | | Unspecified | Population based | Region | Santermans 2016 | 100.0 |
| Guinea, Liberia, Sierra Leone | Dec 2013 - 25 Nov 2014 | | | | 24 - 58 | | | Naive | Population based | Region, Time | Agua-Agum 2015 | 100.0 |
| Guinea, Liberia, Sierra Leone | Dec 2013 - 25 Nov 2014 | | | | 52 - 100 | | | Adjusted | Population based | Age, Occupation, Other, Region, Sex, Time | Agua-Agum 2015 | 100.0 |
| Guinea, Liberia, Sierra Leone | 2013 - 2016 | 83 | | 33,338 | | 46 - 86 | 95% CI | Adjusted | Population based | Age, Other, Region, Symptoms | Forna 2019 | 100.0 |
| Guinea, Liberia, Sierra Leone | 2013 - 2016 | 72 | | 33,338 | | 56 - 80 | 95% CI | Naive | Population based | Age, Other, Region, Symptoms | Forna 2019 | 100.0 |
| Guinea, Liberia, Sierra Leone | 2013 - 2016 | 75 | | 18,644 | | 74 - 77 | 95% CI | Naive | Population based | Age, Other, Region, Symptoms | Forna 2019 | 100.0 |
| Guinea, Liberia, Sierra Leone | Jan 2014 - Feb 2015 | 74 | | | | 68 - 80 | 95% CI | Unspecified | | Age, Level of exposure, Region, Time | Cleaton 2016 | 42.9 |
| Guinea, Liberia, Sierra Leone | 22 Mar 2014 - 20 Aug 2014 | | | | 48 - 74 | | | Unspecified | Unspecified | Region | Althaus 2014 | 42.9 |
| Guinea, Liberia, Sierra Leone | 24 May 2014 - 30 Sep 2014 | | | | 39 - 67 | | | Unspecified | Population based | Region | D'Silva 2017 | 71.4 |
| Liberia | Dec 2013 - 25 Nov 2014 | 43 | | | | | | Unspecified | Community based | | Helleringer 2015 | 100.0 |
| Liberia | 28 Feb 2014 - 1 Dec 2014 | | | | 42 - 47 | | | Unspecified | | | Fallah 2015 | 42.9 |
| Liberia | 20 Mar 2014 - 20 Sep 2014 | 53 | 330 | 619 | | | | Naive | | | Ki 2015 | 0.0 |
| Liberia | 20 Mar 2014 - 20 Sep 2014 | 82 | | | | | | Unspecified | | | Ki 2015 | 0.0 |
| Liberia | 21 Mar 2014 - 31 Dec 2014 | 54 | 229 | 428 | | 49 - 58 | 95% CI | Naive | | Other | Weppelmann 2016 | 100.0 |
| Liberia | 4 Apr 2014 - 29 Mar 2015 | 69 | 2,701 | 3,897 | | 68 - 71 | 95% CI | Naive | Population based | Age, Region, Sex, Time | Furuse 2017 | 100.0 |
| Liberia | 14 May 2014 - 7 Mar 2015 | 59 | 10 | 17 | | | | Unspecified | Community based | | Kuehne 2016 | 28.6 |
| Liberia | 27 May 2014 - 22 Dec 2014 | | | | 34 - 83 | | | Unspecified | Unspecified | Other, Time | Russo 2016 | 42.9 |
| Liberia | 27 May 2014 - 21 Dec 2014 | | | | 12 - 70 | | | Adjusted | Population based | Time | Siettos 2015 | 14.3 |
| Liberia | 7 Jul 2014 - 22 Sep 2014 | 55 | 76 | 139 | 9 - 93 | 60 - 64 | 95% CI | Naive | | Age, Other, Sex, Symptoms | Yamin 2016 | 100.0 |

| Country | Survey date | Central estimate | Deaths | Cases | Central range | Uncertainty | Uncertainty type | Adjustment | Population Sample | Disaggregated by | Article | QA score (%) |
|---|---|---|---|---|---|---|---|---|---|---|---|---|
| Liberia | Jul 2014 - Dec 2014 | 68 | | | 50 - 92 | 60 - 74 | 95% CI | Unspecified | Hospital based | Age, Other, Time | Lindblade 2015 | 100.0 |
| Liberia | 28 Aug 2014 - 18 Dec 2014 | | | | 33 - 80 | | | Unspecified | Hospital based | Other | Jeremiah Matson 2022 | 100.0 |
| Liberia | 15 Sep 2014 - 4 Jan 2015 | 51 | 82 | 160 | | | | Naive | Hospital based | | Levine 2015 | 100.0 |
| Liberia | 2014 | 50 | | | | | | Unspecified | Unspecified | | Rivers 2014 | 42.9 |
| Liberia, Sierra Leone | 15 Sep 2014 - 31 Dec 2015 | 58 | 244 | 424 | | | | Unspecified | Hospital based | | Alusio 2019 | 100.0 |
| Liberia, Sierra Leone | 15 Sep 2014 - 15 Sep 2015 | 46 | 6 | 13 | | | | Adjusted | Hospital based | Other | Henwood 2017 | 71.4 |
| Liberia, Sierra Leone | 15 Sep 2014 - 15 Sep 2015 | 54 | 87 | 162 | | | | Unspecified | Hospital based | Other | Henwood 2017 | 71.4 |
| Liberia, Sierra Leone | 15 Sep 2014 - 15 Sep 2015 | 58 | 268 | 465 | 38 - 93 | | | Adjusted | Hospital based | Age, Region, Sex | Roshania 2016 | 100.0 |
| Liberia, Sierra Leone | Sep 2014 - Sep 2015 | 58 | 244 | 424 | | | | Unspecified | Hospital based | Other | Garbern 2019 | 50.0 |
| Liberia, Sierra Leone | Sep 2014 - Sep 2015 | 58 | | | | | | Unspecified | Hospital based | Age, Other, Region, Sex | Peters 2019 | 85.7 |
| Multi-country: Africa, Europe, USA (n = 10) | Dec 2013 - Mar 2016 | 40 | 11,325 | 28,652 | | | | Unspecified | | Occupation, Region | Shultz 2016 | 42.9 |
| Multi-country: Africa, Europe, USA (n = 6) | Jan 2014 - 12 Oct 2014 | 55 | 425 | 236 | | | | Unspecified | Unspecified | | Hossain 2016 | 0.0 |
| Multi-country: Europe & USA (n = 9) | Aug 2014 - Dec 2015 | 18 | 5 | 27 | | | | Naive | Hospital based | | Uyeki 2016 | 80.0 |
| Nigeria | 20 Jul 2014 - 20 Oct 2014 | 39 | | | | 14 - 71 | 95% CrI | Unspecified | Unspecified | | Althaus 2015 | 71.4 |
| Nigeria | 20 Jul 2014 - 1 Oct 2014 | 40 | 8 | 20 | | 22 - 61 | 95% CI | Unspecified | Population based | | Fasina 2014 | 71.4 |
| Nigeria | Jul 2014 - Unspecified | 40 | 7 | 20 | 22 - 46 | | | Adjusted | Contact based | Occupation | Musa 2015 | 60.0 |
| Nigeria | 2014 | | | | 33 - 46 | | | Naive | Population based | Sex | Fawole 2016 | 40.0 |
| Sierra Leone | Dec 2013 - 25 Nov 2014 | 31 | | | | | | Unspecified | Community based | | Helleringer 2015 | 100.0 |
| Sierra Leone | 1 May 2014 - 31 Jan 2015 | 69 | | | | | | Unspecified | Hospital based | Occupation | Senga 2016 | 85.7 |
| Sierra Leone | 1 May 2014 - 31 Jan 2015 | 74 | | | | | | Unspecified | Hospital based | Occupation | Senga 2016 | 85.7 |
| Sierra Leone | 12 May 2014 - 13 Nov 2015 | | | | 19 - 46 | | | Unspecified | | Other, Time | Chen 2021 | 71.4 |
| Sierra Leone | 19 May 2014 - 11 Jan 2015 | 51 | | | | 12 - 107 | 95% CI | Unspecified | Hospital based | | Li 2020 | 0.0 |
| Sierra Leone | 23 May 2014 - 31 Jan 2015 | 74 | 2,536 | 3,422 | | 73 - 76 | 95% CrI | Naive | Population based | | Wong 2016 | 75.0 |
| Sierra Leone | 25 May 2014 - 18 Jun 2014 | 74 | 64 | 87 | 33 - 94 | | | Adjusted | Hospital based | Age, Other, Region, Sex | Schieffelin 2014 | 100.0 |
| Sierra Leone | 27 May 2014 - 21 Dec 2014 | | | | 24 - 67 | | | Adjusted | Population based | Time | Siettos 2015 | 14.3 |
| Sierra Leone | 28 May 2014 - 31 Dec 2015 | 46 | | | | | | Unspecified | | Other, Time | Potluri 2020 | 100.0 |
| Sierra Leone | May 2014 - Apr 2015 | 94 | 29 | 31 | | 79 - 99 | 95% CI | Unspecified | Household based | | Caleo 2018 | 100.0 |
| Sierra Leone | 1 Jun 2014 - 7 Nov 2015 | 37 | 1,816 | 4,954 | 25 - 73 | | | Naive | Population based | Age | Lamunu 2017 | 40.0 |
| Sierra Leone | 23 Jun 2014 - 5 Oct 2014 | 53 | 259 | 488 | | 49 - 58 | 95% CI | Unspecified | Hospital based | Occupation | Dallatomasina 2015 | 85.7 |
| Sierra Leone | 26 Jun 2014 - 12 Oct 2014 | 51 | 270 | 525 | 24 - 85 | | | Adjusted | Hospital based | Occupation, Other, Region, Time | Fitzpatrick 2015 | 80.0 |
| Sierra Leone | 26 Jun 2014 - 31 Dec 2014 | 57 | 52 | 91 | 46 - 82 | | | Unspecified | Hospital based | Age, Other, Region, Sex | Shah 2016 | 100.0 |
| Sierra Leone | Jun 2014 - Apr 2015 | | | | 0 - 38 | | | Unspecified | Hospital based | Age, Other, Sex | Kangbai 2019 | 85.7 |
| Sierra Leone | Jun 2014 - Apr 2015 | 26 | 279 | 1,077 | 1 - 100 | | | Adjusted | Hospital based | Age, Occupation, Other, Sex, Symptoms | Kangbai 2020 | 100.0 |
| Sierra Leone | Jun 2014 - Apr 2015 | 26 | 248 | 938 | 3 - 100 | | | Adjusted | Hospital based | Age, Occupation, Other, Sex, Symptoms | Kangbai 2020 (b) | 80.0 |
| Sierra Leone | 1 Jul 2014 - 30 Jun 2015 | 24 | 93 | 454 | 0 - 40 | | | Unspecified | Population based | Region | Miglietta 2019 | 28.6 |
| Sierra Leone | Jul 2014 - Nov 2014 | 86 | 42 | 49 | | 73 - 94 | 95% CI | Naive | Population based | | Ajelli 2015 | 71.4 |
| Sierra Leone | 14 Aug 2014 - 31 Mar 2015 | 57 | 160 | 282 | 0 - 100 | 51 - 63 | 95% CI | Adjusted | Hospital based | Age, Other, Region, Sex, Symptoms, Time | Fitzgerald 2016 | 80.0 |
| Sierra Leone | 12 Sep 2014 - 23 Feb 2015 | 44 | 110 | 249 | | | | Naive | Hospital based | Age, Other, Region, Sex, Symptoms | Theocharopoulos 2017 | 60.0 |
| Sierra Leone | 13 Sep 2014 - 26 Nov 2014 | 48 | 99 | 205 | 10 - 100 | | | Adjusted | Hospital based | Age, Occupation, Other, Region, Sex | Kangbai 2021 | 80.0 |
| Sierra Leone | Sep 2014 - Jan 2015 | 66 | 142 | 216 | | | | Adjusted | Community based | Other | Crowe 2016 | 85.7 |
| Sierra Leone | 1 Oct 2014 - 21 Mar 2015 | 49 | 139 | 285 | | | | Naive | Hospital based | | Ji 2016 | 100.0 |
| Sierra Leone | 1 Oct 2014 - 14 Nov 2014 | 69 | | | | | | Unspecified | Hospital based | Age | Qin 2015 | 42.9 |
| Sierra Leone | 1 Oct 2014 - 9 Nov 2014 | 60 | 51 | 85 | | | | Unspecified | Hospital based | | Yan 2015 | 100.0 |
| Sierra Leone | 11 Oct 2014 - 9 Dec 2014 | 51 | 38 | 74 | | | | Unspecified | Population based | | DeSilva 2019 | 40.0 |
| Sierra Leone | Oct 2014 - Mar 2015 | 73 | 98 | 134 | | | | Adjusted | Hospital based | Sex | Li 2016 (b) | 100.0 |
| Sierra Leone | Oct 2014 - Apr 2015 | 66 | 94 | 142 | 55 - 74 | 58 - 74 | 95% CI | Unspecified | Population based | Age, Sex | Muoghalu 2017 | 100.0 |
| Sierra Leone | 15 Nov 2014 - 18 Jan 2015 | 55 | 76 | 139 | 9 - 93 | | | Naive | Hospital based | Age, Other, Sex, Symptoms | Xu 2016 | 100.0 |
| Sierra Leone | 1 Dec 2014 - 28 Feb 2015 | 64 | 32 | 50 | | | | Unspecified | Community based | Level of exposure | Stehling-Ariza 2016 | 100.0 |
| Sierra Leone | 1 Dec 2014 - 28 Feb 2015 | 63 | 27 | 43 | | | | Unspecified | Community based | Level of exposure | Stehling-Ariza 2016 | 100.0 |
| Sierra Leone | 8 Dec 2014 - 9 Jan 2015 | 37 | 55 | 150 | 24 - 40 | | | Unspecified | Hospital based | Age, Other, Sex | Hunt 2015 | 71.4 |

| Country | Survey date | Central estimate | Deaths | Cases | Central range | Uncertainty | Uncertainty type | Adjustment | Population Sample | Disaggregated by | Article | QA score (%) |
|---|---|---|---|---|---|---|---|---|---|---|---|---|
| Sierra Leone | 12 Dec 2014 - 14 Mar 2015 | 46 | | | | | | Unspecified | Hospital based | Age | Rudolf 2017 | 60.0 |
| Sierra Leone | 12 Dec 2014 - 14 Mar 2015 | 67 | | | | | | Unspecified | Hospital based | Age | Rudolf 2017 | 60.0 |
| Sierra Leone | 13 Dec 2014 - 20 Apr 2015 | 50 | 50 | 101 | | | | Naive | Hospital based | | Lanini 2015 | 100.0 |
| Sierra Leone | 19 Dec 2014 - 17 Feb 2015 | 58 | 18 | 31 | 35 - 86 | | | Unspecified | Hospital based | Sex | Haaskjold 2016 | 85.7 |
| Sierra Leone | 21 Dec 2014 - 15 Aug 2015 | | | | 10 - 39 | | | Unspecified | | Time | Siettos 2016 | 42.9 |
| Sierra Leone | Dec 2014 - Jan 2015 | 78 | 28 | 36 | | 61 - 90 | 95% CI | Naive | Unspecified | | Richardson 2016 | 80.0 |
| Sierra Leone | 2014 | 75 | | | | | | Unspecified | Unspecified | | Rivers 2014 | 42.9 |
| Sierra Leone | 13 Jan 2015 - 5 Apr 2015 | 57 | 8 | 14 | | | | Adjusted | Community based | | Li 2016 | 100.0 |
| Sierra Leone | 16 Jan 2015 - 10 Mar 2015 | 47 | 29 | 62 | | | | Adjusted | Other | | Jiang 2017 | 80.0 |
| Sierra Leone | Jun 2015 - Jan 2016 | 58 | 227 | 395 | | | | Naive | Household based | | Bower 2016 | 14.3 |
| **DRC, 2014** | | | | | | | | | | | | |
| DRC | 26 Jul 2014 - 7 Oct 2014 | 74 | | | | | | Unspecified | Community based | | Maganga 2014 | 28.6 |
| DRC | 13 Aug 2014 - 8 Sep 2014 | 62 | 23 | 37 | 62 - 68 | | | Adjusted | Unspecified | Other | Li 2019 | 71.4 |
| DRC | 28 Aug 2014 - 8 Nov 2014 | 48 | 12 | 25 | | | | Unspecified | Hospital based | | Nanclares 2016 | 80.0 |
| **DRC, 2017** | | | | | | | | | | | | |
| DRC | 27 Mar 2017 - 1 Jul 2017 | 50 | 4 | 8 | | | | Naive | | | Nsio 2019 | 100.0 |
| **DRC, 2018** | | | | | | | | | | | | |
| DRC | 4 Apr 2018 - 27 May 2018 | 52 | 25 | 48 | | | | Naive | Community based | | Kelly 2019 | 28.6 |
| DRC | 5 Apr 2018 - 30 May 2018 | 50 | 25 | 50 | 36 - 64 | 36 - 64 | 95% CI | Naive | Unspecified | | Ahuka-Mundeke 2018 | 100.0 |
| DRC | 5 Apr 2018 - 30 May 2018 | 56 | 25 | 50 | 39 - 72 | 39 - 72 | 95% CI | Adjusted | Unspecified | | Ahuka-Mundeke 2018 | 100.0 |
| **DRC, 2018-2020** | | | | | | | | | | | | |
| DRC | Jul 2018 - May 2020 | 62 | 61 | 579 | 3 - 94 | | | Unspecified | Hospital based | Age, Occupation | Baller 2022 | 0.0 |
| DRC | 5 Aug 2018 - 2 Feb 2020 | | | | 50 - 80 | | | Unspecified | Hospital based | Time | Potluri 2022 | 71.4 |
| DRC | 5 Aug 2018 - 2 Feb 2020 | | | | 50 - 80 | | | Unspecified | Community based | Time | Potluri 2022 | 71.4 |
| DRC | 7 Dec 2018 - 29 Jan 2020 | 63 | 157 | 248 | | | | Adjusted | Hospital based | | Rupani 2022 | 80.0 |
| DRC | 7 Dec 2018 - 29 Jan 2020 | 25 | 34 | 137 | | | | Adjusted | Hospital based | | Rupani 2022 | 80.0 |
| DRC | 30 Mar 2019 - 3 Aug 2019 | 55 | 117 | 213 | | | | Adjusted | Hospital based | | Kasereka 2020 | 100.0 |
| DRC | 30 Mar 2019 - 3 Aug 2019 | 23 | 10 | 44 | | | | Adjusted | Hospital based | | Kasereka 2020 | 100.0 |
| DRC | 24 Apr 2019 - 14 Oct 2019 | 45 | 139 | 307 | 41 - 71 | | | Adjusted | Hospital based | Age | Kjaldgaard 2022 | 100.0 |
| DRC | 12 Jun 2019 - 18 May 2020 | 12 | 4 | 32 | | | | Unspecified | Other | | Keita 2023 | 28.6 |
| DRC | 12 Jun 2019 - 18 May 2020 | 48 | 42 | 87 | | | | Unspecified | Community based | | Keita 2023 | 28.6 |
| Uganda | 18 Sep 2020 - Dec 2000 | 71 | 12 | 17 | | | | Unspecified | Contact based | | Francesconi 2003 | 57.1 |
| Uganda | 18 Sep 2020 - Dec 2000 | 100 | 10 | 10 | | | | Unspecified | Contact based | | Francesconi 2003 | 57.1 |
| **Multiple outbreaks** | | | | | | | | | | | | |
| DRC | Aug 1976 - Nov 2012 | 79 | 526 | | 44 - 96 | 76 - 82 | 95% CI | Adjusted | Unspecified | Other | Rosello 2015 | 100.0 |
| Multi-country: Africa (n = 7) | 1976 - 2014 | 66 | 1,595 | 2,411 | | | | Unspecified | | Region, Time | Shultz 2016 | 42.9 |

###### 4.5.2 Risk factors associated with severity

Table S22: A) Risk factors associated with death, B) Protective factors associated with recovery. Total is the number of parameters extracted for each risk/protective factor.

**A**

| Risk Factor for Death | Significant | | | Not significant | | | Unspecified significance | | | Total |
|---|---|---|---|---|---|---|---|---|---|---|
| | Adjusted | Not adjusted | Unspecified | Adjusted | Not adjusted | Unspecified | Adjusted | Not adjusted | Unspecified | |
| Age | 18 | 12 | 11 | 9 | 12 | 4 | 1 | 1 | 0 | 68 |
| Sex | 3 | 5 | 2 | 14 | 14 | 7 | 1 | 1 | 0 | 47 |
| Occupation | 1 | 1 | 1 | 6 | 5 | 2 | 0 | 0 | 1 | 17 |
| Hospitalisation | 2 | 2 | 2 | 1 | 0 | 0 | 0 | 0 | 0 | 7 |
| Close contact | 1 | 1 | 0 | 1 | 2 | 0 | 0 | 0 | 0 | 5 |
| Comorbidity | 0 | 0 | 0 | 0 | 4 | 1 | 0 | 0 | 0 | 5 |
| Funeral | 1 | 0 | 0 | 0 | 2 | 0 | 0 | 0 | 0 | 3 |
| Household contact | 0 | 0 | 0 | 2 | 1 | 0 | 0 | 0 | 0 | 3 |
| Non-household contact | 0 | 0 | 0 | 2 | 0 | 0 | 0 | 0 | 0 | 2 |
| Other | 26 | 22 | 11 | 21 | 22 | 9 | 2 | 1 | 0 | 114 |

**B**

| Protective Factor for Recovery | Significant | | | Not significant | | | Total |
|---|---|---|---|---|---|---|---|
| | Adjusted | Not adjusted | Unspecified | Adjusted | Not adjusted | Unspecified | |
| Age | 1 | 1 | 0 | 0 | 1 | 1 | 4 |
| Sex | 0 | 0 | 0 | 1 | 1 | 1 | 3 |
| Hospitalisation | 0 | 0 | 1 | 0 | 0 | 0 | 1 |
| Other | 1 | 2 | 0 | 1 | 1 | 0 | 5 |

#### 4.6 Mutation rates

Table S23: Mutation rates

| Country | Survey date | Central estimate | Central range | Central type | Uncertainty | Uncertainty type | Genome site | Population Sample | Sample size | Disaggregated by | Species | Article | QA score (%) |
|---|---|---|---|---|---|---|---|---|---|---|---|---|---|
| **Evolutionary rate (Substitutions/site/year ($10^{-4}$))** | | | | | | | | | | | | | |
| DRC | May 2007 - Feb 2009 | | 1.5 - 7.5 | Other/Unspecified | | | Whole genome, Glycoprotein, Nucleoprotein | Other | | | Zaire | Grard 2011 | 57.1 |
| DRC, Guinea, Liberia, Sierra Leone | 2013 - 2016 | 4.7 | 1 - 10 | Mean | 3.4 - 5.7 | 95% CrI | Whole genome, Partial sequences | | 85 | Other | Zaire | Li 2019 | 71.4 |
| Guinea, Liberia, Sierra Leone | 2013 - 2015 | 7.3 | | Other/Unspecified | | | Glycoprotein | | 600 | | Zaire | Dudas 2019 | 71.4 |
| Guinea, Liberia, Sierra Leone | 2013 - 2015 | 8.2 | | Other/Unspecified | | | Whole genome | | 600 | | Zaire | Dudas 2019 | 71.4 |
| Guinea, Mali, Sierra Leone | Dec 2013 - Jan 2015 | 14.2 | | Mean | 12.2 - 16.2 | 95% CrI | Whole genome | | 179 | | Zaire | Carroll 2015 | 57.1 |
| Guinea, Sierra Leone | 2014 - 2015 | 12.5 | | Mean | | | Partial genome | | 318 | | Zaire | Park 2015 | 100.0 |
| Sierra Leone | May 2014 - Sep 2015 | 9.6 | 5 - 9.63 | Mean | 8.6 - 10.6 | 95% CrI | Whole genome | | | Other | Zaire | Whitmer 2018 | 100.0 |
| Guinea | 12 Feb 2021 - 4 Mar 2021 | 11 | | Other/Unspecified | | | Whole genome, Partial genome | | 12 | | Zaire | Keita 2021 | 71.4 |
| DRC | Aug 1976 - Aug 2018 | | 2.4 - 8.6 | Mean | | | Whole genome | Population based | 15 | Other, Time | Zaire | Mbala-Kingebeni 2019 | 100.0 |
| Multi-country: Africa (n = 5) | 1976 - 2014 | 10.75 | | Median | 9.32 - 12.2 | HPDI 95% | Glycoprotein | | 65 | Other | Zaire | Azarian 2015 | 100.0 |
| Multi-country: Africa, Asia, Europe, USA (n = 9) | 1976 - 2008 | 10.93 | 7.66 - 13.94 | Other/Unspecified | 0.52 - 24.61 | HPDI 95% | Glycoprotein | | | | All species | Li 2014 | 100.0 |
| Multi-country: Africa, Asia, Europe, USA (n = 9) | 1976 - 2008 | 23.14 | | Other/Unspecified | 16.49 - 31.66 | HPDI 95% | Polymerase (L) | | | | All species | Li 2014 | 100.0 |
| **Mutation rate (Substitutions/site/year ($10^{-4}$))** | | | | | | | | | | | | | |
| Liberia | Sep 2014 - Feb 2015 | 9.17 | | Other/Unspecified | 0.523 | Other | Whole genome | Hospital based | 25 | Other | Zaire | Kugelman 2015 | 28.6 |
| Liberia | Sep 2014 - Feb 2015 | | 9.44 - 15.67 | Other/Unspecified | | | Whole genome | Hospital based | 25 | Other | Zaire | Kugelman 2015 | 28.6 |
| **Substitution rate (Substitutions/site/year ($10^{-4}$))** | | | | | | | | | | | | | |
| DRC | 3 Nov 2007 - 10 Sep 2012 | 8.4 | | Mean | 1.1 - 21 | 95% CrI | Whole genome | | 12 | | Bundibugyo | Hulseberg 2021 | 100.0 |
| Guinea | Feb 2014 - Unspecified | 10.7 | | Mean | | | Glycoprotein | | | | Zaire | Dudas 2014 | 60.0 |
| Guinea | Unspecified | 2 | | Mean | 1 - 3 | HPDI 95% | Partial genome | | 49 | | All species | Membrebe 2019 | 57.1 |
| Guinea, Mali, Sierra Leone | Mar 2014 - Dec 2014 | | 8.7 - 9.1 | Mean | 6.8 - 11 | 95% CrI | Whole genome | Hospital based | 195 | | Zaire | Simon-Loriere 2015 | 71.4 |
| Guinea, Mali, Sierra Leone | 2014 | 13 | | Mean | | | Whole genome | | 106 | | Zaire | Hoenen 2015 | 83.3 |
| Guinea, Sierra Leone | 28 Sep 2014 - 11 Nov 2014 | 12.3 | | Other/Unspecified | 10.4 - 14.1 | HPDI 95% | Whole genome | Community based | 175 | | Zaire | Tong 2015 | 42.9 |
| DRC | 8 May 2018 - 24 Jul 2018 | 36 | | Mean | 12.2 - 59.9 | HPDI 95% | Whole genome | | 15 | | Zaire | Mbala-Kingebeni 2019 (b) | 71.4 |
| Multi-country: Africa (n = 5) | 15 Aug 1976 - 4 Jun 2018 | 6.9 | | Mean | 5.5 - 8.3 | HPDI 95% | Whole genome | | 50 | | Zaire | Mbala-Kingebeni 2019 (b) | 71.4 |
| Multi-country: Africa, Asia, Europe, USA (n = 6) | 1976 - 1995 | 0.36 | | Mean | 0.109 | Standard Error | Glycoprotein | | | | Unspecified | Suzuki 1997 | 42.9 |
| DRC | Unspecified | 1.4 | | Mean | 0.473 - 2.45 | 95% CrI | Whole genome | | | | Zaire | Vrancken 2020 | 85.7 |

#### 4.7 Transmission Models

Table S24: Summary of EVD model types.

| Model type | Total |
|---|---|
| Compartmental | 210 |
| Other or combination | 49 |
| Branching process | 19 |
| Agent/Individual based | 17 |

Table S25: Summary of EVD model assumptions. Total is the number of models with this assumption extracted (assumptions were not extracted for all models).

| Assumptions | Total |
|---|---|
| **Compartmental** | |
| Homogeneous mixing | 83 |
| Heterogeneity in transmission rates between groups | 43 |
| Heterogeneity in transmission rates over time | 42 |
| Latent period the same as incubation period | 27 |
| **Other or combination** | |
| Heterogeneity in transmission rates between groups | 15 |
| Heterogeneity in transmission rates over time | 11 |
| Homogeneous mixing | 9 |
| Latent period the same as incubation period | 5 |
| Age dependent susceptibility | 1 |
| **Branching process** | |
| Heterogeneity in transmission rates over time | 4 |
| Heterogeneity in transmission rates between groups | 3 |
| Homogeneous mixing | 1 |
| **Agent/Individual based** | |
| Heterogeneity in transmission rates between groups | 9 |
| Heterogeneity in transmission rates over time | 4 |
| Latent period the same as incubation period | 3 |
| Age dependent susceptibility | 2 |

Table S26: Models overview. * = Zaire, ** = Bundibugyo, *** = Bundibugyo, Sudan, Taï Forest and Zaire

| Stochastic or Deterministic | Theoretical model | Interventions | Code available | Assumptions | Article | QA score (%) |
|---|---|---|---|---|---|---|
| **Agent/Individual based** | | | | | | |
| Stochastic | Yes | Behaviour changes, Contact tracing, Hospitals, Safe burials, Treatment Centres | | Age dependent susceptibility, Heterogeneity in transmission rates between groups | Ajelli 2018 | 100.00 |
| Stochastic | Yes | Vaccination | | Heterogeneity in transmission rates between groups, Unspecified | Bisanzio 2023 | 100.00 |
| Stochastic | | | | Heterogeneity in transmission rates between groups and over time | Cope 2014 | 100.00 |
| Stochastic | | Hospitals, Other | | Unspecified | Fadikar 2018 | 100.00 |
| Stochastic | | | Yes | Heterogeneity in transmission rates between groups, Latent period the same as incubation period | Kahn 2020 | 100.00 |
| Stochastic | | Other, Safe burials, Treatment Centres | | Heterogeneity in transmission rates between groups | Merler 2015 | 100.00 |
| Stochastic | | Vaccination | | Heterogeneity in transmission rates between groups | Merler 2016 | 100.00 |
| Stochastic | | Contact tracing, Hospitals, Safe burials | | Heterogeneity in transmission rates over time | Rizzo 2016 | 100.00 |
| Stochastic | | Hospitals | | Unspecified | Gomes 2014 | 85.71 |
| Stochastic | | | | Unspecified | Gomez-Barroso 2017 | 80.00 |
| Stochastic | | Contact tracing, Hospitals, Safe burials, Treatment Centres, Vaccination | | Age dependent susceptibility, Heterogeneity in transmission rates between groups | Ajelli 2016 | 71.43 |
| Stochastic | Yes | | | Heterogeneity in transmission rates between groups | Kustudic 2021 | 50.00 |
| Stochastic | | | | Unspecified | Fallah 2015 | 42.86 |
| Stochastic | | Other, Safe burials | | Heterogeneity in transmission rates over time, Latent period the same as incubation period | Siettos 2016 | 42.86 |
| Stochastic | | | | Heterogeneity in transmission rates between groups and over time, Latent period the same as incubation period | Uekermann 2019 | 28.57 |
| Stochastic | Yes | Behaviour changes, Hospitals, Other, Quarantine, Safe burials | | Unspecified | Venkatramanan 2018 | 25.00 |
| | | | | Unspecified | Khaleque 2017 | 0.00 |
| **Branching process** | | | | | | |
| Stochastic | | | | Unspecified | Djaafara 2021 | 100.00 |
| Stochastic | | Quarantine, Vaccination | Yes | Heterogeneity in transmission rates between groups and over time | Green 2022 | 100.00 |
| Stochastic | | | | Unspecified | Holbrook 2022 | 100.00 |
| Stochastic | | | Yes | Heterogeneity in transmission rates over time | Kelly 2019 (b) | 100.00 |
| Stochastic | | | | Unspecified | Nouvellet 2018 | 100.00 |
| Stochastic | | Other, Quarantine | Yes | Heterogeneity in transmission rates between groups | Yamin 2016 | 100.00 |
| Stochastic | | | | Unspecified | Nishiura 2014 | 75.00 |
| Stochastic | | Quarantine, Safe burials | | Unspecified | Fang 2016 | 71.43 |
| Stochastic | | Vaccination | | Unspecified | Kucharski 2016 | 71.43 |
| Stochastic | | | | Unspecified | Toth 2015 | 71.43 |
| Stochastic | | Hospitals, Safe burials | | Heterogeneity in transmission rates between groups | Drake 2015 | 57.14 |
| Stochastic | | | | Heterogeneity in transmission rates over time | Zhang 2022 | 57.14 |
| Stochastic | | | Yes | Unspecified | Lee 2022 | 50.00 |
| Stochastic | | | | Unspecified | Choi 2019** | 42.86 |
| Stochastic | | | | Unspecified | Jombart 2020 | 42.86 |
| Stochastic | | Vaccination | | Heterogeneity in transmission rates over time | Worden 2019 | 42.86 |
| Stochastic | | Vaccination | | Homogeneous mixing | Kelly 2019 | 28.57 |
| Stochastic | | | | Unspecified | Champredon 2018 | 25.00 |
| Stochastic | Yes | | Yes | Unspecified | Park 2019 | 25.00 |
| **Compartmental** | | | | | | |
| Deterministic | Yes | | | Homogeneous mixing, Latent period the same as incubation period, Unspecified | Almocera 2019 | 100.00 |
| Deterministic | Yes | | | Heterogeneity in transmission rates between groups | Altaf 2019 | 100.00 |
| Deterministic | Yes | Other, Vaccination | | Homogeneous mixing | Anguelov 2020 | 100.00 |
| Stochastic | | | | Homogeneous mixing | Asher 2018 | 100.00 |
| Deterministic | Yes | | | Homogeneous mixing | Baldassi 2016* | 100.00 |

| Stochastic or Deterministic | Theoretical model | Interventions | Code available | Assumptions | Article | QA score (%) |
|---|---|---|---|---|---|---|
| Deterministic | Yes | | | Homogeneous mixing, Latent period the same as incubation period | Berge 2016 | 100.00 |
| Deterministic | Yes | | | Homogeneous mixing, Latent period the same as incubation period | Berge 2018 | 100.00 |
| Deterministic | Yes | Contact tracing, Quarantine | | Homogeneous mixing | Berge 2018 (b) | 100.00 |
| Deterministic | Yes | | | Heterogeneity in transmission rates between groups, Homogeneous mixing | Berge 2018 (c) | 100.00 |
| Deterministic | Yes | | | Heterogeneity in transmission rates between groups, Homogeneous mixing | Berge 2018 (c) | 100.00 |
| Deterministic | Yes | Behaviour changes, Other, Vaccination | | Homogeneous mixing | Berge 2018 (d) | 100.00 |
| Deterministic | Yes | Vaccination | | Heterogeneity in transmission rates between groups, Homogeneous mixing | Bhunu 2016 | 100.00 |
| Deterministic | Yes | Hospitals, Quarantine, Safe burials | | Heterogeneity in transmission rates between groups and over time | Blackwood 2016 | 100.00 |
| Deterministic | | Vaccination | | Heterogeneity in transmission rates between groups, Latent period the same as incubation period | Bodine 2018 | 100.00 |
| Stochastic | | Hospitals, Safe burials, Treatment Centres | | Heterogeneity in transmission rates over time | Camacho 2015 | 100.00 |
| Deterministic | | | | Homogeneous mixing | Chapwanya 2022 | 100.00 |
| | Yes | Treatment, Vaccination | | Heterogeneity in transmission rates between groups and over time | De la Sen 2017 (b) | 100.00 |
| Deterministic | Yes | Safe burials, Treatment, Vaccination | | Unspecified | De la Sen 2019 | 100.00 |
| Stochastic | | | | Homogeneous mixing | Frasso 2016 | 100.00 |
| Stochastic | | Hospitals | | Heterogeneity in transmission rates over time | Funk 2017 | 100.00 |
| Stochastic | | | Yes | Heterogeneity in transmission rates over time | Funk 2018 | 100.00 |
| Stochastic | | | Yes | Heterogeneity in transmission rates over time | Funk 2019 | 100.00 |
| Stochastic | Yes | Other, Vaccination | Yes | Unspecified | Getz 2015 | 100.00 |
| Deterministic | Yes | Hospitals, Quarantine, Treatment | | Unspecified | Gutfraind 2015 | 100.00 |
| Stochastic | Yes | | | Homogeneous mixing | Kamara 2020 | 100.00 |
| Stochastic | | Behaviour changes, Hospitals, Quarantine | | Heterogeneity in transmission rates between groups | Ko 2019 | 100.00 |
| Stochastic | | | Yes | Unspecified | Kramer 2016 | 100.00 |
| Stochastic | | Treatment Centres | | Heterogeneity in transmission rates over time | Kucharski 2015 | 100.00 |
| Deterministic | | | | Unspecified | Leander 2016 | 100.00 |
| Deterministic | | Vaccination | | Heterogeneity in transmission rates between groups and over time, Homogeneous mixing, Latent period the same as incubation period | Lee 2019 | 100.00 |
| Stochastic | | Hospitals, Safe burials | | Homogeneous mixing | Legrand 2007 | 100.00 |
| Stochastic | Yes | | Yes | Homogeneous mixing | Lekone 2006 | 100.00 |
| Deterministic | | Other, Quarantine, Treatment, Vaccination | | Homogeneous mixing | Mbah 2022 | 100.00 |
| Deterministic | Yes | Behaviour changes, Safe burials | | Unspecified | Mhlanga 2019 | 100.00 |
| | Yes | | | Unspecified | Nazir 2020 | 100.00 |
| Stochastic | Yes | | | Homogeneous mixing | Nieddu 2017 | 100.00 |
| Deterministic | Yes | Other | | Unspecified | Njankou 2022 | 100.00 |
| Deterministic | Yes | | | Heterogeneity in transmission rates between groups | Njankou 2023 | 100.00 |
| Stochastic | | Hospitals, Vaccination | | Heterogeneity in transmission rates between groups and over time, Homogeneous mixing | Potluri 2020 | 100.00 |
| Stochastic | | | | Heterogeneity in transmission rates between groups and over time, Homogeneous mixing, Latent period the same as incubation period | Riad 2019 | 100.00 |
| Deterministic | | Vaccination | | Heterogeneity in transmission rates between groups and over time | Robert 2019 | 100.00 |
| Deterministic | | | Yes | Heterogeneity in transmission rates over time, Homogeneous mixing | Santermans 2016 | 100.00 |
| Deterministic | Yes | | | Heterogeneity in transmission rates between groups | Seidu 2020 | 100.00 |
| Stochastic | | | | Unspecified | Shakiba 2021 | 100.00 |
| Deterministic | Yes | Other | | Unspecified | Siewe 2020 | 100.00 |
| Deterministic | | | Yes | Unspecified | Stadler 2014 | 100.00 |
| Deterministic | Yes | | | Latent period the same as incubation period | Verma 2019 | 100.00 |
| | Yes | Hospitals, Safe burials | | Unspecified | Wang 2017 | 100.00 |
| Deterministic | | | | Unspecified | Wiratsudakul 2016 | 100.00 |

| Stochastic or Deterministic | Theoretical model | Interventions | Code available | Assumptions | Article | QA score (%) |
|---|---|---|---|---|---|---|
| | | Treatment | | Unspecified | Zaman 2009 | 100.00 |
| Deterministic | | | Yes | Homogeneous mixing | Abbate 2016 | 85.71 |
| Deterministic | Yes | Behaviour changes | Yes | Homogeneous mixing | Abbate 2016 | 85.71 |
| Deterministic | | Hospitals, Safe burials | | Heterogeneity in transmission rates between groups, Homogeneous mixing | Diaz 2018 | 85.71 |
| Stochastic | | | | Heterogeneity in transmission rates between groups | Enanoria 2015 | 85.71 |
| Deterministic | | Other, Treatment Centres | | Homogeneous mixing | Lewnard 2014 | 85.71 |
| Stochastic | | | | Unspecified | Luo 2019 | 85.71 |
| Stochastic | | | Yes | Heterogeneity in transmission rates between groups and over time | Muleia 2021 | 85.71 |
| Deterministic | | Safe burials | | Latent period the same as incubation period | Sofonea 2018 | 85.71 |
| Stochastic | | Quarantine, Safe burials, Treatment | | Heterogeneity in transmission rates over time | Volz 2014 | 85.71 |
| Deterministic | | Quarantine, Safe burials, Treatment | | Heterogeneity in transmission rates over time | Volz 2014 | 85.71 |
| Deterministic | | Quarantine, Safe burials, Treatment | | Unspecified | Volz 2014 | 85.71 |
| | | Hospitals, Safe burials | | Unspecified | Xia 2015 | 85.71 |
| Deterministic | | | | Heterogeneity in transmission rates over time, Homogeneous mixing | Yang 2015 | 85.71 |
| Stochastic | | | | Unspecified | Gomez-Barroso 2017 | 80.00 |
| Deterministic | Yes | Quarantine | | Unspecified | Abbasi 2020 | 75.00 |
| Deterministic | Yes | Other | | Homogeneous mixing | Agusto 2016* | 75.00 |
| Deterministic | Yes | Hospitals, Quarantine, Vaccination | | Heterogeneity in transmission rates between groups, Homogeneous mixing | Ahmad 2016* | 75.00 |
| Deterministic & Stochastic | | | | Unspecified | Atkins 2016 | 75.00 |
| Deterministic | Yes | Vaccination | | Homogeneous mixing | Brettin 2018 | 75.00 |
| | | | | Unspecified | Burghardt 2016 | 75.00 |
| | | Other | | Heterogeneity in transmission rates between groups and over time, Latent period the same as incubation period | Buyuktahtakin 2018 | 75.00 |
| | Yes | Hospitals | | Heterogeneity in transmission rates between groups | Dautel 2021 | 75.00 |
| Deterministic | | Behaviour changes | | Heterogeneity in transmission rates over time | Djiomba Njankou 2022 | 75.00 |
| Deterministic | Yes | | | Unspecified | Dokuyucu 2020 | 75.00 |
| Deterministic | Yes | | | Homogeneous mixing | El Rhoubari 2019 | 75.00 |
| Deterministic | Yes | | | Unspecified | Farman 2022 (b) | 75.00 |
| Stochastic | | | | Heterogeneity in transmission rates over time | Getz 2018 | 75.00 |
| Deterministic | | | | Heterogeneity in transmission rates over time | Getz 2018 | 75.00 |
| Deterministic | | | | Heterogeneity in transmission rates over time | Getz 2018 | 75.00 |
| Stochastic | | | | Heterogeneity in transmission rates over time | Getz 2018 | 75.00 |
| Deterministic | Yes | Behaviour changes, Other, Safe burials | | Unspecified | Guo 2016 | 75.00 |
| Deterministic & Stochastic | Yes | | | Unspecified | Kamara 2021 | 75.00 |
| Deterministic | Yes | Quarantine | | Heterogeneity in transmission rates between groups | Ngwa 2016 | 75.00 |
| Deterministic | Yes | Other, Quarantine, Treatment, Vaccination | | Heterogeneity in transmission rates between groups | Ojoma 2021 | 75.00 |
| Stochastic | Yes | Contact tracing | | Heterogeneity in transmission rates between groups | Shahtori 2018 | 75.00 |
| Deterministic | Yes | | | Latent period the same as incubation period | Tahir 2019 | 75.00 |
| Stochastic | | Behaviour changes | | Unspecified | Vinson 2016 | 75.00 |
| | | Safe burial, Unspecified | | Homogeneous mixing | Weitz 2015 | 75.00 |
| Deterministic | | Contact tracing, Other, Quarantine | | Heterogeneity in transmission rates over time, Homogeneous mixing, Latent period the same as incubation period, Unspecified | Althaus 2015 | 71.43 |
| Deterministic | | Contact tracing, Other, Quarantine, Safe burials, Treatment | Yes | Heterogeneity in transmission rates over time, Homogeneous mixing, Latent period the same as incubation period, Unspecified | Althaus 2015 (b) | 71.43 |
| Deterministic & Stochastic | | Behaviour changes, Hospitals, Safe burials | | Heterogeneity in transmission rates between groups and over time, Homogeneous mixing | Camacho 2014 | 71.43 |
| Deterministic | | Other, Quarantine, Treatment Centres | Yes | Heterogeneity in transmission rates between groups | D'Silva 2017* | 71.43 |
| Stochastic | | Other, Quarantine, Treatment Centres | Yes | Heterogeneity in transmission rates between groups | D'Silva 2017* | 71.43 |

| Stochastic or Deterministic | Theoretical model | Interventions | Code available | Assumptions | Article | QA score (%) |
|---|---|---|---|---|---|---|
| Stochastic | | | | Heterogeneity in transmission rates between groups | Fasina 2014* | 71.43 |
| Deterministic | | | | Heterogeneity in transmission rates over time | Getz 2019 | 71.43 |
| Deterministic | | Other, Quarantine | Yes | Heterogeneity in transmission rates over time, Homogeneous mixing, Latent period the same as incubation period | Hart 2019 | 71.43 |
| Stochastic | | Vaccination | | Unspecified | Potluri 2022 | 71.43 |
| | | Other, Quarantine, Safe burials, Vaccination | | Heterogeneity in transmission rates between groups and over time | Shen 2015 | 71.43 |
| Stochastic | | Contact tracing | | Unspecified | Webb 2015* | 71.43 |
| Deterministic | | Behaviour changes, Hospitals, Other, Safe burials | | Heterogeneity in transmission rates between groups, Homogeneous mixing | Agusto 2015* | 57.14 |
| Deterministic | | | | Unspecified | Li 2017 (b) | 57.14 |
| Deterministic & Stochastic | | | | Homogeneous mixing | Mouanguissa 2021 | 57.14 |
| Stochastic | | | | Homogeneous mixing | Ndanguza 2017 | 57.14 |
| Deterministic | | Treatment Centres | | Homogeneous mixing | Njankou 2018 | 57.14 |
| Stochastic | | Hospitals, Behaviour changes, Safe burials | | Heterogeneity in transmission rates between groups, Latent period the same as incubation period | Valdez 2015 | 57.14 |
| | | Hospitals | | Unspecified | Webb 2016 | 57.14 |
| Deterministic | | Quarantine, Treatment | | Homogeneous mixing, Latent period the same as incubation period, Heterogeneity in transmission rates over time | White 2015 | 57.14 |
| Deterministic | | Contact tracing, Hospitals, Other, Vaccination | | Heterogeneity in transmission rates over time, Latent period the same as incubation period | Xie 2019 | 57.14 |
| Deterministic | | Other, Safe burials | | Unspecified | Agbomola 2022 | 50.00 |
| Deterministic | Yes | Vaccination | | Homogeneous mixing, Latent period the same as incubation period | Ahmad 2020 | 50.00 |
| Deterministic | Yes | Quarantine | | Homogeneous mixing | Ahmad 2021 | 50.00 |
| Stochastic | Yes | Contact tracing, Other, Quarantine, Safe burials | | Unspecified | Bouba 2023 | 50.00 |
| Stochastic | | | | Unspecified | Brown 2016 | 50.00 |
| Deterministic | Yes | | Yes | Homogeneous mixing | Cheema 2021 | 50.00 |
| Deterministic | | Hospitals, Quarantine | | Homogeneous mixing, Latent period the same as incubation period | Chinyoka 2021 | 50.00 |
| Deterministic | Yes | Treatment, Vaccination | | Homogeneous mixing | Chukwu 2020 | 50.00 |
| Deterministic | | Safe burials, Treatment Centres, Vaccination | | Heterogeneity in transmission rates between groups and over time | Diakite 2016* | 50.00 |
| Deterministic | | Quarantine | | Heterogeneity in transmission rates between groups, Homogeneous mixing, Latent period the same as incubation period | Diakite 2016 | 50.00 |
| Deterministic | Yes | Hospitals, Safe burials | | Unspecified | Gaffey 2018 | 50.00 |
| Deterministic | | Hospitals, Treatment, Vaccination | | Latent period the same as incubation period | Jiang 2017 (b) | 50.00 |
| Deterministic | | Hospitals, Other, Safe burials | | Homogeneous mixing | Juga 2023 | 50.00 |
| Deterministic | Yes | | | Unspecified | Lopez 2016 | 50.00 |
| Deterministic | Yes | Quarantine | | Homogeneous mixing | Nisar 2023 | 50.00 |
| Deterministic | Yes | Vaccination | | Homogeneous mixing, Latent period the same as incubation period | Obeng-Kusi 2021 | 50.00 |
| Deterministic | | | | Unspecified | Ord 2018 | 50.00 |
| Deterministic | Yes | Behaviour changes, Other, Vaccination | | Homogeneous mixing | Rachah 2016 | 50.00 |
| Deterministic | Yes | Treatment, Vaccination | | Homogeneous mixing | Rachah 2016 (b) | 50.00 |
| Deterministic | Yes | Vaccination | | Homogeneous mixing | Rachah 2017 | 50.00 |
| | Yes | | | Unspecified | Rafiq 2020 | 50.00 |
| Deterministic | Yes | Behaviour changes, Hospitals, Safe burials | | Heterogeneity in transmission rates over time, Homogeneous mixing | Roy 2017 | 50.00 |
| Deterministic | Yes | Behaviour changes, Hospitals, Other, Vaccination | | Homogeneous mixing, Latent period the same as incubation period | Seck 2022 | 50.00 |
| Deterministic | | | | Unspecified | Singh 2023 | 50.00 |
| Deterministic | | | | Heterogeneity in transmission rates over time | Smirnova 2019 | 50.00 |
| Deterministic | Yes | | | Homogeneous mixing | Srivastava 2020 | 50.00 |
| Deterministic | Yes | | | Unspecified | Srivastava 2020 (b) | 50.00 |
| Deterministic | Yes | | | Unspecified | Srivastava 2021 | 50.00 |
| Deterministic | Yes | Vaccination | | Homogeneous mixing | Tahir 2018 | 50.00 |
| Deterministic | Yes | | | Homogeneous mixing, Latent period the same as incubation period | Tandon 2018 | 50.00 |

| Stochastic or Deterministic | Theoretical model | Interventions | Code available | Assumptions | Article | QA score (%) |
|---|---|---|---|---|---|---|
| Deterministic | Yes | | | Unspecified | Telionis 2020 | 50.00 |
| Deterministic | Yes | | Yes | Homogeneous mixing | Tsanou 2017 | 50.00 |
| Deterministic & Stochastic | | Quarantine, Vaccination | | Homogeneous mixing | Tulu 2017 (b) | 50.00 |
| Deterministic | Yes | | | Homogeneous mixing, Latent period the same as incubation period | Tulu 2017 (c) | 50.00 |
| Stochastic | Yes | | | Unspecified | Wang 2019 | 50.00 |
| Stochastic | Yes | Treatment Centres | | Heterogeneity in transmission rates between groups | Yin 2021 | 50.00 |
| Deterministic | Yes | Behaviour changes, Other, Treatment | | Heterogeneity in transmission rates between groups | Zakary 2017 | 50.00 |
| Deterministic | | Other | | Heterogeneity in transmission rates over time, Homogeneous mixing, Latent period the same as incubation period, Unspecified | Althaus 2014 | 42.86 |
| Deterministic | | Other, Quarantine, Safe burials, Treatment Centres | | Heterogeneity in transmission rates over time, Homogeneous mixing, Latent period the same as incubation period | Barbarossa 2015 | 42.86 |
| Stochastic | | Vaccination | | Unspecified | Camacho 2015 (b) | 42.86 |
| | | | | Unspecified | Evans 2015 | 42.86 |
| Deterministic | | Quarantine, Safe burials | | Homogeneous mixing | Imran 2017 | 42.86 |
| Deterministic | | Other | Yes | Homogeneous mixing | Ivorra 2015 | 42.86 |
| Deterministic | | Treatment | | Homogeneous mixing | Li 2015 | 42.86 |
| Deterministic | | | | Unspecified | Ndanguza 2013* | 42.86 |
| Stochastic | | Hospitals, Safe burials | | Heterogeneity in transmission rates between groups, Homogeneous mixing | Ponce 2019 | 42.86 |
| Deterministic | | Vaccination | | Homogeneous mixing | Rachah 2018 | 42.86 |
| Deterministic & Stochastic | | Contact tracing, Other, Treatment | | Homogeneous mixing, Latent period the same as incubation period | Rivers 2014 | 42.86 |
| Deterministic | | | Yes | Heterogeneity in transmission rates over time | Stockdale 2021 | 42.86 |
| Stochastic | | | Yes | Heterogeneity in transmission rates over time | Tang 2023 | 42.86 |
| Stochastic | | | Yes | Heterogeneity in transmission rates over time | Vossler 2022 | 42.86 |
| Deterministic | | | | Unspecified | Wang 2015 | 42.86 |
| Stochastic | | | Yes | Unspecified | Ward 2023 | 42.86 |
| Stochastic | | Other | | Heterogeneity in transmission rates over time | Chowell 2004 | 28.57 |
| Deterministic | | Hospitals | | Unspecified | Juga 2020 | 28.57 |
| Deterministic | | Behaviour changes, Hospitals, Other | | Homogeneous mixing | Juga 2021 | 28.57 |
| Deterministic | | Quarantine | | Heterogeneity in transmission rates between groups | Khan 2015 | 28.57 |
| Deterministic | | Other | | Heterogeneity in transmission rates between groups and over time, Homogeneous mixing | Siewe 2020 (b) | 28.57 |
| Deterministic | | Behaviour changes, Quarantine | | Unspecified | Tadmon 2022 | 28.57 |
| Deterministic | | | | Unspecified | Area 2015* | 25.00 |
| Deterministic | Yes | | | Unspecified | Atangana 2014 | 25.00 |
| Deterministic | Yes | | Yes | Unspecified | Bachinsky 2013 | 25.00 |
| Deterministic | Yes | | | Homogeneous mixing | Bhardwaj 2021 | 25.00 |
| Stochastic | | | | Unspecified | Champredon 2018 | 25.00 |
| Deterministic | Yes | Contact tracing, Hospitals, Other, Quarantine | | Unspecified | Chen 2014*** | 25.00 |
| | Yes | | | Unspecified | Dike 2017 | 25.00 |
| Deterministic | Yes | | | Homogeneous mixing | Emile Franc Doungmo 2016* | 25.00 |
| | Yes | | | Unspecified | Gong 2019 | 25.00 |
| Deterministic | Yes | Other | | Homogeneous mixing | Grigorieva 2015 | 25.00 |
| Deterministic | Yes | Other, Quarantine | | Homogeneous mixing | Grigorieva 2017 | 25.00 |
| Deterministic | Yes | Hospitals, Other, Safe burials | | Homogeneous mixing | Grigorieva 2018 | 25.00 |
| Deterministic | Yes | | | Unspecified | Hasan 2023 | 25.00 |
| Stochastic | | | | Unspecified | Kaur 2021 | 25.00 |
| Deterministic | | Behaviour changes | | Heterogeneity in transmission rates between groups | Levy 2017 | 25.00 |

| Stochastic or Deterministic | Theoretical model | Interventions | Code available | Assumptions | Article | QA score (%) |
|---|---|---|---|---|---|---|
| Deterministic | | | | Unspecified | Mubayi 2021 | 25.00 |
| | Yes | Hospitals, Behaviour changes | | Heterogeneity in transmission rates between groups | Shah 2019 | 25.00 |
| Deterministic | Yes | Contact tracing, Hospitals, Quarantine, Safe burials, Vaccination | | Unspecified | Tadmon 2022 (b) | 25.00 |
| Deterministic & Stochastic | | Quarantine, Vaccination | | Homogeneous mixing | Tulu 2017 | 25.00 |
| Deterministic | Yes | | | Homogeneous mixing | Zafar 2021 | 25.00 |
| Deterministic | | Quarantine, Safe burials | | Unspecified | Do 2016 | 14.29 |
| Deterministic | | Other | | Heterogeneity in transmission rates between groups | Dong 2015 | 14.29 |
| | Yes | Safe burials, Vaccination | | Heterogeneity in transmission rates over time | Area 2018 | 0.00 |
| Stochastic | Yes | | | Unspecified | Atangana 2021 | 0.00 |
| Deterministic & Stochastic | | Quarantine, Vaccination | | Unspecified | Din 2022 | 0.00 |
| | Yes | Quarantine | | Unspecified | Farman 2022 | 0.00 |
| | Yes | | | Unspecified | Koca 2018 | 0.00 |
| | | Quarantine, Treatment, Vaccination | | Unspecified | Li 2015 (b) | 0.00 |
| Deterministic | | | | Homogeneous mixing | Li 2020 | 0.00 |
| | Yes | Quarantine, Vaccination | | Unspecified | Liu 2019 | 0.00 |
| Deterministic | | Behaviour changes | | Heterogeneity in transmission rates between groups and over time | Long 2018 | 0.00 |
| Deterministic | Yes | Contact tracing, Quarantine | | Homogeneous mixing | Madubueze 2018 | 0.00 |
| Deterministic | Yes | | | Unspecified | Momani 2022 | 0.00 |
| Deterministic | Yes | Other, Treatment, Vaccination | | Unspecified | Okyere 2020 | 0.00 |
| Deterministic | | Hospitals, Quarantine, Safe burials | | Unspecified | Ouemba Tasse 2022 | 0.00 |
| Deterministic | | | | Unspecified | Pan 2021 | 0.00 |
| Deterministic | Yes | Vaccination | | Homogeneous mixing | Rachah 2015 | 0.00 |
| Stochastic | Yes | | | Heterogeneity in transmission rates between groups | Rashid 2022 | 0.00 |
| Deterministic | Yes | | | Unspecified | Raza 2020 | 0.00 |
| | Yes | Treatment | | Unspecified | Shaikh 2023 | 0.00 |
| Deterministic | Yes | | | Homogeneous mixing | Singh 2020 | 0.00 |
| Deterministic | | | | Heterogeneity in transmission rates between groups | Sivaraman 2022 | 0.00 |
| Deterministic | Yes | | | Homogeneous mixing | Zhang 2020 | 0.00 |
| **Other or combination** | | | | | | |
| Deterministic | Yes | Quarantine | | Unspecified | Adams 2016 | 100.00 |
| Stochastic | | Vaccination | | Heterogeneity in transmission rates between groups | Chowell 2019 (b) | 100.00 |
| | | | Yes | Unspecified | De Maio 2018 | 100.00 |
| Deterministic | | | | Unspecified | De la Sen 2017 | 100.00 |
| Deterministic | Yes | | | Homogeneous mixing | Feng 2016 | 100.00 |
| Stochastic | | | | Heterogeneity in transmission rates over time | Ganyani 2018 | 100.00 |
| | | | | Unspecified | Gustafson 2017 | 100.00 |
| Stochastic | Yes | | | Unspecified | Jacobsen 2018 | 100.00 |
| Stochastic | | | Yes | Heterogeneity in transmission rates between groups | Kiskowski 2014 | 100.00 |
| Stochastic | | | | Heterogeneity in transmission rates between groups | Kiskowski 2016 | 100.00 |
| Stochastic | | | Yes | Heterogeneity in transmission rates between groups and over time | Kraemer 2019 | 100.00 |
| Stochastic | | | | Heterogeneity in transmission rates between groups and over time, Latent period the same as incubation period | Lau 2017 (b) | 100.00 |
| Stochastic | | Behaviour changes | | Heterogeneity in transmission rates over time, Homogeneous mixing | Lee 2019 | 100.00 |
| Stochastic | Yes | | | Unspecified | Pell 2016 | 100.00 |
| Deterministic | | | Yes | Heterogeneity in transmission rates between groups | Santermans 2016 | 100.00 |
| Stochastic | | | | Heterogeneity in transmission rates over time | Shaman 2014 | 100.00 |

| Stochastic or Deterministic | Theoretical model | Interventions | Code available | Assumptions | Article | QA score (%) |
|---|---|---|---|---|---|---|
| Deterministic | | | | Unspecified | Smirnova 2017 | 100.00 |
| | | | | Unspecified | Tuitea 2018 | 100.00 |
| | Yes | | | Unspecified | Zabinski 2018 | 100.00 |
| Stochastic | | Contact tracing, Quarantine | | Heterogeneity in transmission rates over time | Chan 2020 | 85.71 |
| | | | Yes | Homogeneous mixing | Chowell 2019 | 85.71 |
| Stochastic | | | Yes | Heterogeneity in transmission rates between groups and over time, Latent period the same as incubation period | Lau 2017 | 85.71 |
| Stochastic | | | | Homogeneous mixing | White 2007* | 85.71 |
| | Yes | | | Unspecified | Mangiarotti 2016* | 75.00 |
| Deterministic | Yes | | | Homogeneous mixing | Upadhyay 2016* | 75.00 |
| Deterministic | | Other, Vaccination | | Unspecified | Chen 2021 | 71.43 |
| Stochastic | | Behaviour changes | Yes | Unspecified | Pruyt 2015 | 71.43 |
| | | | | Unspecified | Saulnier 2017 | 71.43 |
| | | | | Unspecified | Taylor 2016 | 71.43 |
| | | | | Unspecified | Browne 2015 | 57.14 |
| Stochastic | | | | Heterogeneity in transmission rates between groups and over time | Martinez 2022 | 57.14 |
| Stochastic | | | | Heterogeneity in transmission rates over time | Park 2022 | 57.14 |
| Stochastic | | | Yes | Homogeneous mixing | Vaughan 2019 | 57.14 |
| Deterministic | Yes | | | Heterogeneity in transmission rates between groups | Ahmed 2023 | 50.00 |
| Stochastic | | | | Unspecified | Backer 2016 | 50.00 |
| Stochastic | | | | Heterogeneity in transmission rates between groups | Chen 2016 | 50.00 |
| Stochastic | Yes | Vaccination | Yes | Age dependent susceptibility, Heterogeneity in transmission rates between groups | Rachah 2016 (b) | 50.00 |
| Deterministic | Yes | | | Unspecified | Yamazaki 2018 | 50.00 |
| Stochastic | | | | Unspecified | Burch 2017 | 42.86 |
| Deterministic | | | | Unspecified | Jombart 2020 | 42.86 |
| Stochastic | | | | Latent period the same as incubation period | Russo 2016 | 42.86 |
| Deterministic | | | | Heterogeneity in transmission rates between groups, Homogeneous mixing | Scarpino 2015 | 42.86 |
| Deterministic | | Vaccination | | Homogeneous mixing, Latent period the same as incubation period, Unspecified | Fisman 2014 | 28.57 |
| Deterministic | | | | Homogeneous mixing, Latent period the same as incubation period, Unspecified | Fisman 2014 (b) | 28.57 |
| | | | | Unspecified | Hsieh 2015 | 28.57 |
| Stochastic | Yes | Behaviour changes | | Heterogeneity in transmission rates between groups and over time | Halvorsen 2022 | 25.00 |
| Deterministic | Yes | Contact tracing, Other, Quarantine, Safe burials | | Heterogeneity in transmission rates between groups and over time | Ivorra 2020 | 25.00 |
| Stochastic | | Safe burials | | Heterogeneity in transmission rates between groups | Siettos 2015* | 14.29 |
| Stochastic | Yes | | | Unspecified | Burkhead 2015 | 0.00 |

### 5 Pathogen Epidemiology Review Group (PERG) membership

Table S27: Pathogen Epidemiology Review Group (PERG) membership

| First Name | Surname | Affiliation |
|---|---|---|
| Aaron | Morris | University of Oxford |
| Alpha | Forna | University of Georgia |
| Amy | Dighe | Johns Hopkins |
| Anne | Cori | Imperial College London |
| Arran | Hamlet | Imperial College London |
| Ben | Lambert | Manchester University |
| Charlie | Whittaker | Imperial College London |
| Christian | Morgenstern | Imperial College London |
| Cyril | Geismar | Imperial College London |
| Dariya | Nikitin | Imperial College London |
| David | Jorgensen | Imperial College London |
| Ed | Knock | Imperial College London |
| Gina | Cuomo-Dannenburg | Imperial College London |
| Hayley | Thompson | PATH |
| Isobel | Routledge | UCSF |
| Janetta | Skarp | Imperial College London |
| Joseph | Hicks | Imperial College London |
| Juliette | Unwin | University of Bristol |
| Keith | Fraser | Imperial College London |
| Kelly | Charniga | Imperial College London |
| Kelly | McCain | Imperial College London |
| Lily | Geidelberg | Imperial College London |
| Lorenzo | Cattarino | UKHSA |
| Mara | Kont | Imperial College London |
| Marc | Baguelin | Imperial College London |
| Natsuko | Imai | Imperial College London (Wellcome Trust) |
| Nima | Moghaddas | Imperial College London |
| Patrick | Doohan | Imperial College London |
| Rebecca | Nash | Imperial College London |
| Richard | Shepherd | Imperial College London |
| Ruth | McCabe | University of Oxford |
| Sabine | van Elsland | Imperial College London |
| Sangeeta | Bhatia | Imperial College London |
| Sreejith | Radhakrishnan | University of Glasgow |
| Thomas | Rawson | Imperial College London |
| Tristan | Naidoo | Imperial College London |
| Zulma | Cucunuba Perez | Pontificia Universidad Javeriana |
| Jack | Wardle | Imperial College London |
